# Mapping the Functional Landscape of *KCNQ1* to Define Ion Channel Mechanisms and Arrhythmia Risk

**DOI:** 10.64898/2025.12.15.25341924

**Authors:** M. Lorena Harvey, Ellen Osborn, Megan C. Lancaster, Jeremy E. Smith, Qianyi Shen, Temidayo Abe, Ruoyi Cai, Ayesha Muhammad, Rose Kannankeril, Tao Yang, Maria E. Calandranis, Daniel J. Blackwell, Richard E. Dolder, Jessa L. Aldridge, Matthew R. Fleming, Marcia A. Blair, Eduardo Soria, Jack W. O’Sullivan, Giovanni Davogustto, Brett M. Kroncke, Calum A. MacRae, Bjorn C. Knollmann, Frederick P. Roth, Victoria N. Parikh, Euan A. Ashley, Jamie I. Vandenberg, Chai-Ann Ng, Dan M. Roden, Andrew M. Glazer

## Abstract

Loss-of-function variants in *KCNQ1* are the primary cause of congenital Long QT Syndrome (LQTS), characterized by QT prolongation and increased risk of fatal arrhythmias. The surge in genetic testing continues to uncover vast numbers of variants of uncertain significance in Mendelian disease genes, including *KCNQ1*. Using four multiplexed assays, we mapped the functional landscape of *KCNQ1*, identifying trafficking, gating, and dominant negative variants. We provide functional evidence for 13,403 variants, including 1,757 with strong pathogenic and 6,660 with moderate benign evidence. Structure-function analyses revealed variant enrichment in regions regulating trafficking, multimerization, ubiquitylation, and channel gating. Loss-of-function variants showed a risk ratio of 263 (95% CI 241-286) in an LQTS cohort and were significantly associated with QTc prolongation and LQTS diagnoses in biobank participants. By characterizing the functional impact of nearly all *KCNQ1* variants, we provide critical insights into arrhythmia mechanisms and open new avenues for precision medicine in LQTS.

**GRAPHICAL ABSTRACT:** 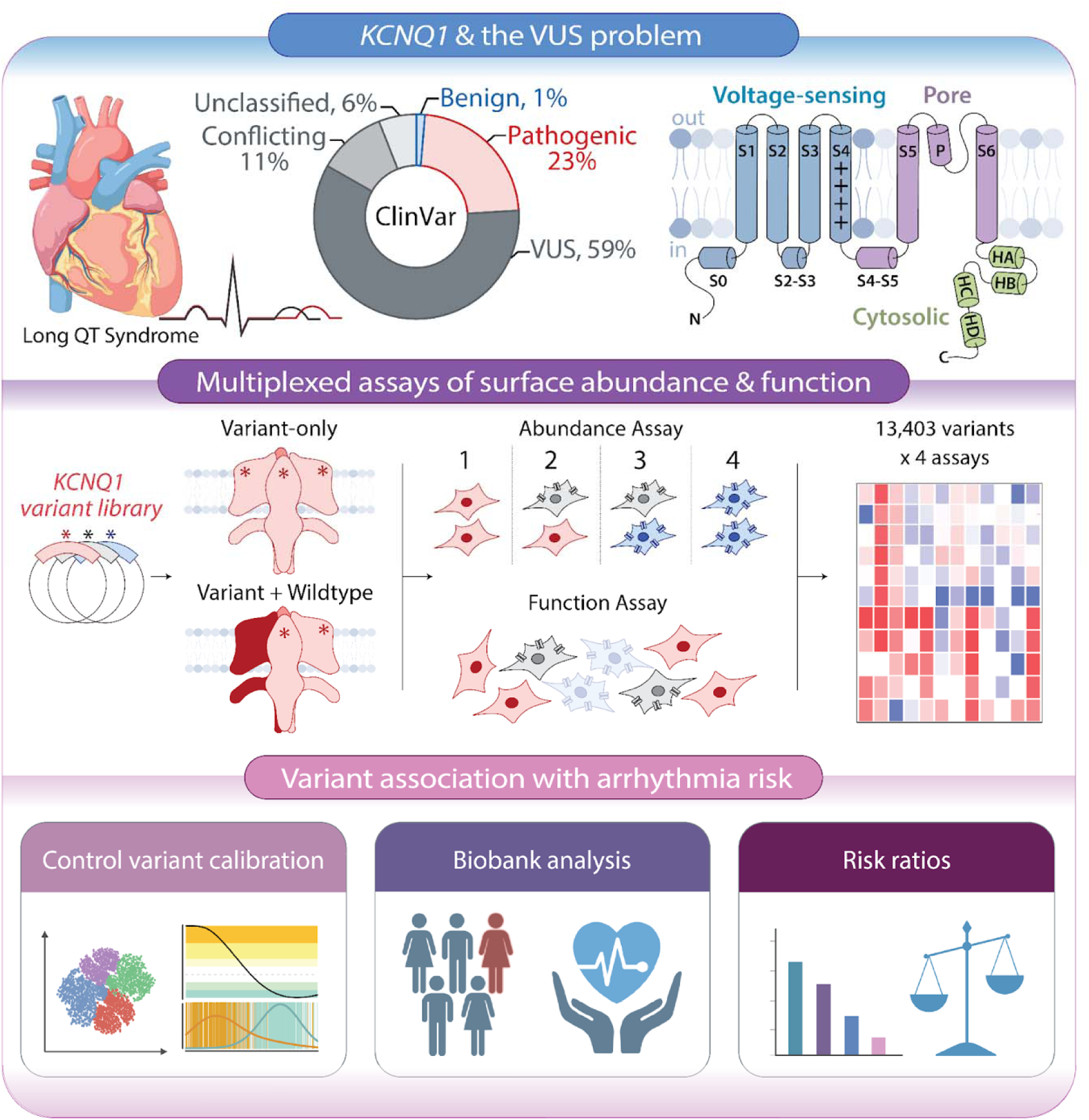

## Introduction

*KCNQ1* plays a critical role in cardiac electrophysiology and inherited arrhythmia risk. Loss-of-function variants in *KCNQ1* are the most common cause of the congenital long QT syndrome (LQTS).^1^ LQTS is characterized by QT prolongation on the electrocardiogram, which confers an increased risk of potentially fatal arrhythmias.^2,3^ Most *KCNQ1*-related LQTS cases arise in heterozygous carriers of loss-of-function variants. The much rarer Jervell and Lange-Nielsen Syndrome (JLNS) is usually caused by biallelic *KCNQ1* loss-of-function variants and is characterized by both severe QT prolongation and congenital sensorineural hearing loss. Over 250 rare *KCNQ1* variants have been identified in patients with Long QT Syndrome Type 1 (LQT1) or JLNS (Figure 1A).^4,5^ Additionally, a small number of *KCNQ1* gain-of-function variants have been associated with atrial fibrillation or short QT syndrome.^6,7^ Because LQTS is treatable with beta blocker therapy, stellate ganglion block, or implantable cardioverter defibrillators, *KCNQ1* has been designated by the American College of Medical Genetics and Genomics (ACMG) as a key “actionable” gene, for which incidentally discovered pathogenic variants should be reported.^8^ Identification of pathogenic *KCNQ1* variants can also enable cascade genetic screening in families.^9^ Thus, understanding which *KCNQ1* variants confer increased arrhythmia risk is essential for precision cardiovascular medicine.

**Figure 1:**
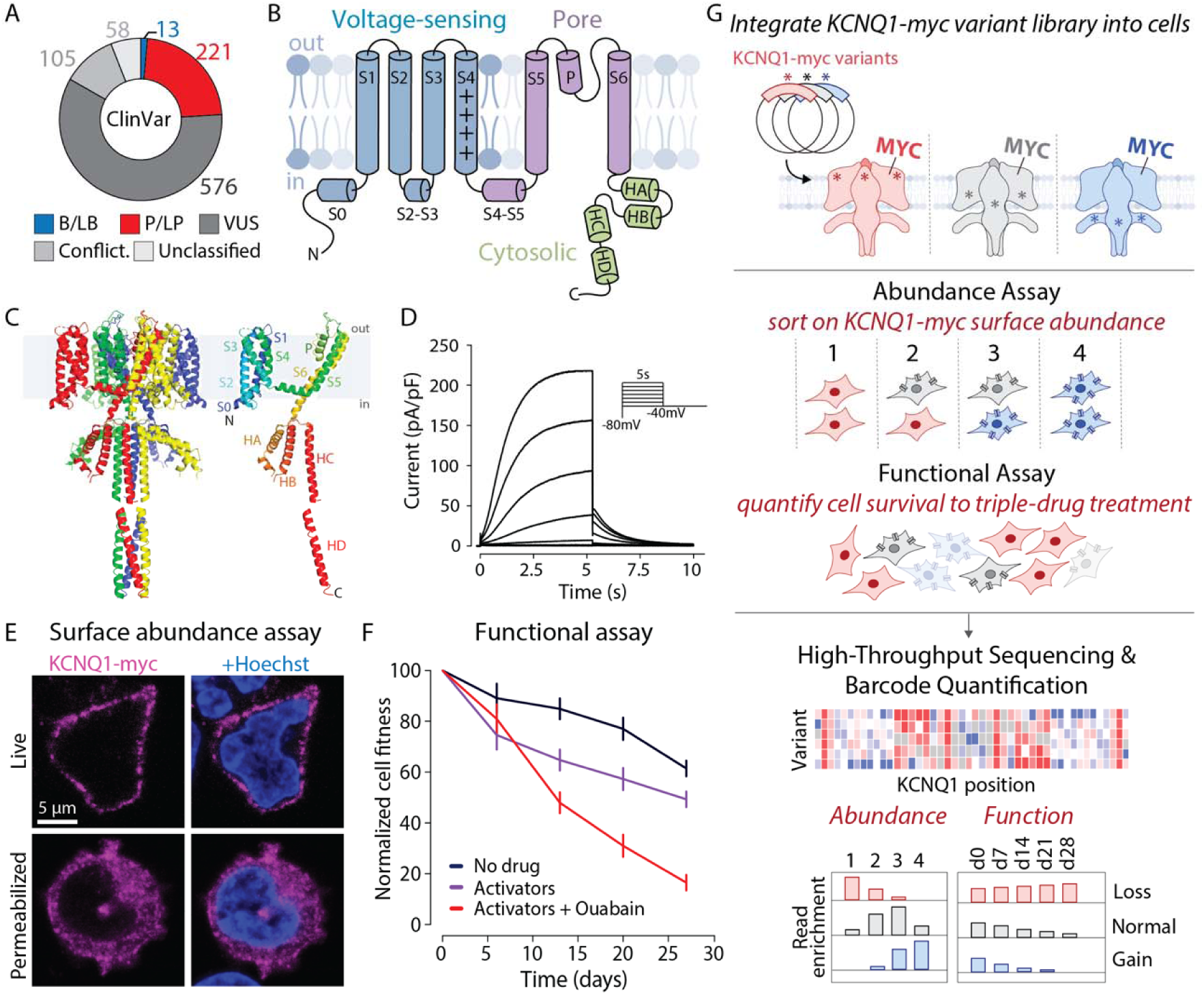
Overview of *KCNQ1* MAVE assays. A) Classification of *KCNQ1* missense variants from ClinVar (accessed on 2/11/25). B) Schematic of a K_V_7.1 monomer, highlighting the voltage-sensing domain (blue), the pore domain (pore), and the cytosolic region (green). C) Tetrameric (left) and monomeric (right) structures of K_V_7.1. The tetramer shows the spatial organization and interactions of four monomers (red, green, blue, yellow), while the monomer highlights the different helices. Most of the structure is from CryoEM structure PDB:6UZZ,^14^ but the HD helix is not present in this structure and is added from PDB:3BJ4, a crystal structure of the HD helix.^52^ D) Representative current trace of WT *I*_Ks_ (*KCNQ1*+*KCNE1*) in Chinese Hamster Ovary cells. The inset represents the voltage protocol used. E) Detection of myc-tagged *KCNQ1* for cell surface expression: anti-myc staining of live cells (top) and fixed/permeabilized cells (bottom). F) Measurement of the normalized cell fitness of HEK293 cells stably expressing WT *KCNQ1* compared to control cells under varying drug conditions. Black: no drug; Purple: two channel activators (ML277 and R-L3); Red: two channel activators (ML277 and RL-3) and ouabain. Data represent mean and standard error from two biological replicates. G) Overview of experimental workflow of MAVE assays of *KCNQ1* protein surface abundance and function.

*KCNQ1* encodes K_V_7.1, the pore-forming subunit of the voltage-gated potassium channel complex that generates the potassium current *I*_Ks_. This slow repolarizing current plays a key role in helping cardiomyocytes repolarize to negative membrane potentials at the end of the action potential. The complex also includes calmodulin and a beta subunit, encoded by *KCNE1*, which modulate channel activity.^10,11^ K_V_7.1 assembles as a homotetramer, with each monomer containing six transmembrane helices (S1-S6; Figure 1B-C). The S1-S4 helices and the adjacent S0 and S2-3 linker helices form the voltage-sensing domain (VSD). In the VSD, the positively charged S4 helix shifts outward upon cell depolarization, translating a shift of membrane potential into protein conformational changes and channel opening. The S5 and S6 helices, along with the pore helix and pore loop, form the pore domain responsible for potassium ion flux. The C-terminal cytosolic domain contains four additional helices, HA, HB, HC, and HD.^12,13^ HA and HB facilitate binding to calmodulin, and HC and HD form coiled-coil motifs critical for K_V_7.1 tetramerization.^14^ Other regions provide binding sites for the key channel activators KCNE1 and phosphatidylinositol 4,5-bisphosphate (PIP_2_).^14–16^ These sites coordinate precise gating and ion-selective functions to generate *I*_Ks_ (Figure 1D). *KCNQ1* variants can disrupt channel function primarily through two mechanisms: (1) by altering protein abundance at the cell surface, typically due to protein misfolding, or (2) by altering the channel’s ability to open or close appropriately, i.e., changing gating properties.^17^ In heterozygous individuals, loss-of-function variants may disrupt function via haploinsufficiency or through dominant negative effects (interfering with the function of the wildtype allele).^18^

Accurate clinical classification of *KCNQ1* variants is critical for advancing cardiovascular genomic medicine and guiding the diagnosis and treatment of inherited arrhythmias. The ACMG framework categorizes variants as pathogenic, likely pathogenic, likely benign, benign, or as a variant of uncertain significance (VUS).^19^ Most *KCNQ1* missense variants in the NIH database ClinVar are classified as VUS (59%) or have conflicting interpretations (11%; Figure 1A).

Furthermore, the *KCNQ1* variants reported to date in ClinVar represent only a small subset of all possible changes (8% of 12,844 possible missense variants), and ∼50% of rare variants detected in large newly sequenced cohorts are not yet present in ClinVar.^20^ *KCNQ1* variant function is conventionally assessed by manual patch clamp experiments examining individual variants.^21,22^ Recent work has also used automated patch clamping to examine dozens of variants.^23,24^ However, these experiments have not yet been calibrated to current ACMG guidelines using large numbers of benign and pathogenic controls. In addition, the rapid expansion of genetic testing has outpaced conventional functional assay capabilities, contributing to the VUS problem.^20^ Multiplexed Assays for Variant Effects (MAVEs) offer a promising solution by simultaneously assessing the function of thousands of variants using multiplexed assays coupled with deep sequencing.^25^ Well-validated MAVE functional studies, which incorporate large numbers of known positive and negative controls, can provide evidence in the ACMG scheme that a variant is pathogenic (PS3) or benign (BS3).^19,26^ This approach has not yet been applied to *KCNQ1*.

Here, we report MAVE functional measurements of 13,403 *KCNQ1* coding variants (94% of 14,196 possible coding variants) using fluorescence-activated cell sorting and cell survival-based assays in both variant-only and heterozygous (wildtype+variant) contexts. We identify over 2,500 variants with altered functional properties, including 339 variants likely affecting channel gating and 838 dominant negative variants. We demonstrate that the scores are highly correlated with clinical classifications and risk of LQTS. These datasets provide new insights into potassium channel biology, prospectively identify thousands of new deleterious variants, and highlight MAVEs as a key enabler of precision medicine.

## Results

### Development of KCNQ1 MAVE Assays

We first developed MAVE-compatible assays of K_V_7.1 surface abundance and function. We used antibody labeling of a myc epitope tag in the first extracellular loop of K_V_7.1,^27^ which we confirmed could measure channel surface abundance in non-permeabilized Human Embryonic Kidney (HEK293) cells (Figure 1E). We also developed a novel assay assessing channel function that relies on a long-term fitness defect of *KCNQ1*-expressing cells (Figure 1F, S1). When expressed for 28 days, WT *KCNQ1* caused a mild fitness defect (normalized fitness of 61% ± 3% s.e.m.), which was amplified (to 49% ± 2%) upon addition of two channel activators (ML277 and R-L3) together. Cell fitness was most reduced (to 17% ± 0.5%) when the two activators were combined with ouabain, an inhibitor of Na+/K+ ATPase, which maintains the potassium gradient across the plasma membrane. We term this assay (two activators and ouabain) a “triple-drug function assay.” To deploy these surface abundance and fitness assays in comprehensive MAVEs of *KCNQ1* coding variants (Figure 1G), we began by creating a comprehensive *KCNQ1* variant library. We mutagenized a plasmid with wildtype *KCNQ1* with inverse PCR mutagenesis with degenerate primers and tagged mutant plasmids with random 18mer barcodes. High-throughput sequencing of the plasmid library revealed that the library contained 266,868 barcodes corresponding to 13,706 *KCNQ1* variants (mean of 19.5 barcodes/variant; Figure S2). Using this library, we obtained at least one MAVE measurement for 13,403 variants. These included 12,223 missense, 544 synonymous, and 636 nonsense variants (Table S1, S2), collectively representing 94% of all possible *KCNQ1* coding variants. We rescaled the MAVE scores so that 1 corresponded to wild-type function and 0 corresponded to complete loss of function. To assess the quality of our MAVE datasets, we generated individual measurements of 24 variants and compared the scores to clinical classifications, arrhythmia cohorts, biobank cohorts, and computational predictors. Our complete project dataset is presented in File S1.

### MAVE of KCNQ1 Surface Abundance

Many variants in plasma membrane proteins cause defects in protein folding and/or cell surface trafficking, resulting in altered protein abundance at the cell surface.^28^ To evaluate how *KCNQ1* variants influence surface abundance, we introduced the pooled library of barcoded *KCNQ1* variants into HEK293 cells engineered with a “landing pad.”^29^ After integration of a KCNQ1 variant plasmid into the landing pad, each cell expresses a single variant at a consistent expression level.^29^ We then labeled live cells with a fluorophore-conjugated anti-myc antibody, performed flow sorting to sort the cells based on fluorescence intensity, and conducted deep sequencing of barcodes (Figure 1G). Surface abundance scores were generated for 93% of all possible coding variants (Figure 2A) and showed high correlations between replicates (Spearman ρ=0.89-0.92; Table S2). We defined early nonsense (residues 1–610) and late nonsense (611–676) variants and identified 2,321 loss-of-abundance, 1,333 partial loss-of-abundance, and 305 gain-of-abundance missense variants (Figure 2B). To assess the dataset, we compared MAVE abundance scores to single-variant measurements of cell surface abundance. The MAVE scores matched well (ρ=0.87) with 138 literature-reported single-variant measurements of surface abundance,^17,30,31^ as well as an additional 26 new single-variant measurements we generated using flow cytometry (Figure 2C-D, Table S3). A small number (21/541 or 3.8%) of synonymous variants had MAVE scores below 0.5. In follow-up single-variant measurements of 7 of these variants, all 7 had near-normal surface abundance, suggesting that these measurements are due to experimental noise in the MAVE experiment (Figure S3, Table S3). Comparing missense variant scores with clinical classifications revealed that 26/28 (93%) of benign/likely benign variants had normal surface abundance, while only 59/217 (27%) of pathogenic/likely pathogenic variants had normal surface abundance (Figure 2E, Table S4).

**Figure 2:**
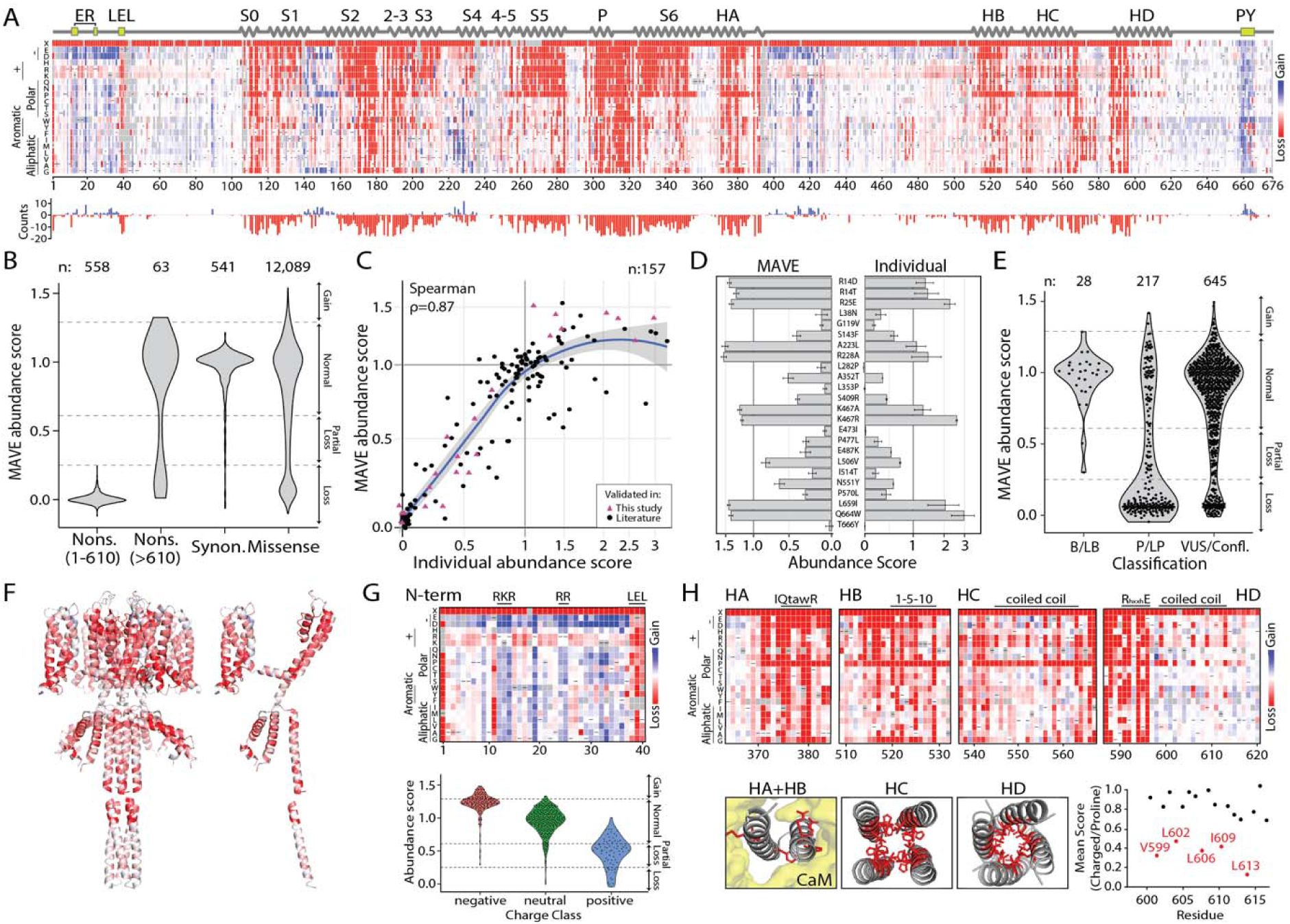
MAVE of K_V_7.1 surface abundance. A *KCNQ1* variant library was integrated into HEK293 landing pad cells, and surface abundance was quantified by staining, sorting, and deep sequencing. The dataset was generated from 5 biological replicates. A) Heatmap of residue-wise variant effects with WT residues indicated by black dashes, alongside a bar plot of variant counts by abundance effect (red: loss, blue: gain). B) Normalized abundance scores stratified by variant class. (C) Correlation of MAVE scores with 24 variants assessed in our study (purple triangles) and 133 variants curated from the literature (black circles) (n=157). Axes are presented on a pseudo-log scale. One outlier value (x=5.01, y=1.29) was removed from the plot. D) Abundance measurements of 24 individual variants compared with their corresponding MAVE scores. Individual measurements represent mean and standard error from 2 biological replicate samples per variant. E) MAVE surface abundance scores for ClinVar variants classified as B/LB, P/LP, or VUS/Conflicting. F) MAVE abundance scores mapped onto the K_V_7.1 protein structure. (red: low, white: WT-like, blue: high) G) Top: Heatmap of N-terminal variant effects with ER retention motifs (RKR, RR) and an LEL trafficking motif highlighted. Bottom: Mean abundance scores stratified by charge effect (negative, neutral, positive). See Figure S4 for full-length analysis. H) Heatmaps of variant effects in cytosolic helices HA–HD. HA+HB and HC structures (panel F, panel H) are from cryo-EM PDB:6UZZ^14^; HD is from the crystal structure PDB:3BJ4^52^. Bottom right: Mean abundance scores for charged or proline substitutions at residues 598–616 (HD coiled-coil domain).

Structure-function analysis revealed mutational sensitivity of key protein regions. *KCNQ1* does not have a signal peptide, but uses its first transmembrane segment as a signal anchor to insert into the membrane.^14^ We discovered mutational sensitivity of other key motifs in the N-terminus, including an LEL_38-40_ trafficking motif^32^ and previously undescribed ER retention motifs (RKR_12-14_ and RR_24-25_; Figure 2G). The N-terminal 40 residues and a large region between HA and HB had broad electric charge sensitivity (Figure 2G, S4). In both regions, most mutations with more negative charges had increased surface abundance while mutations with more positive charges had reduced abundance (p=1.5×10^-85^ and p=1.0×10^-204^, Kruskal-Wallis test). This effect was especially pronounced for mutations of a conserved poly-lysine motif (KKKKFK_419-424_) not previously implicated in channel function (Figure S4).^33^ Transmembrane helices S1-S6 were sensitive to charged and polar mutations (Figure 2A, F). The P helix and adjacent P loop, which line the central potassium-conducting pore, had high mutational intolerance (Figure 2F). Calmodulin-binding HA and HB helices showed a broad mutational sensitivity to negatively charged residues and helix-disrupting prolines (Figure 2G). In addition, they were sensitive to mutation of key residues critical for calmodulin binding: the IQ motif in HA (IQxxxR_375-380_) and the 1-5-10 motif in HB (MxxxVxxxxF_520-529_). Hydrophobic coiled-coil residues in HC and HD, which are important for channel multimerization, exhibited a periodic 3/7 residue mutational sensitivity to charged or proline mutations (Figure 2G)^14,27,34,35^. This corresponds to the heptad repeat pattern (a-g), where positions (a) and (d) play a critical role in coiled-coil function. In addition, mutations in charged residues in the HD helix that have been proposed to make inter-helix salt bridges,^36,37^ including the RhxxhE_591-596_ motif^36^ and R_594_, also caused loss of abundance (Figure S5). Nearly every variant in a C-terminal PY motif (LPxYxxx_659-665_) caused gain-of-abundance, and a few variants in this region caused loss-of-abundance. This “degron” motif is a binding site for E3 ubiquitin ligases, such as Nedd4-2, which ubiquitinate K_V_7.1 and target it for degradation.^38^

### MAVE of KCNQ1 Function

The surface abundance assay can only detect variants that disrupt cell surface abundance, but cannot detect variants that have normal abundance but disrupt channel gating, such as pathogenic variants A341V and R366W.^31,39^ To detect a broader set of function-disrupting variants, we deployed the triple-drug function assay (Figure 1G) on the library of *KCNQ1* variants (Figure 3). Gain-of-function (GOF) variants located in the PY motif decreased in frequency over time faster than synonymous variants, while loss-of-function (LOF) variants, including nonsense mutations from residues 1–610, increased in frequency compared to other variant types (Figure S6). Variant frequencies in initial versus final pools were used to calculate function scores, which were generated for 85% of possible variants (Figure 3A). Scores were consistent across six replicates (ρ = 0.46–0.54; Table S2). Loss-of-function variants were clustered in transmembrane and cytoplasmic helices, while gain-of-function variants localized to the C-terminal PY ubiquitination motif (Figure 3A-B). Early nonsense variants (1–610) were mostly loss-of-function. Late nonsense variants (611+) varied, including many nonsense variants which removed the PY motif and had gain-of-function scores (Figure 3C-D). Unexpectedly, nonsense variants from residues 239–307 had elevated function scores (Figure S6). Experiments across two plasmid systems with nonsense variants and a true *KCNQ1* truncation suggested that this phenomenon results from cryptic partial readthrough of the stop codon in some contexts (Figure S6).

**Figure 3.**
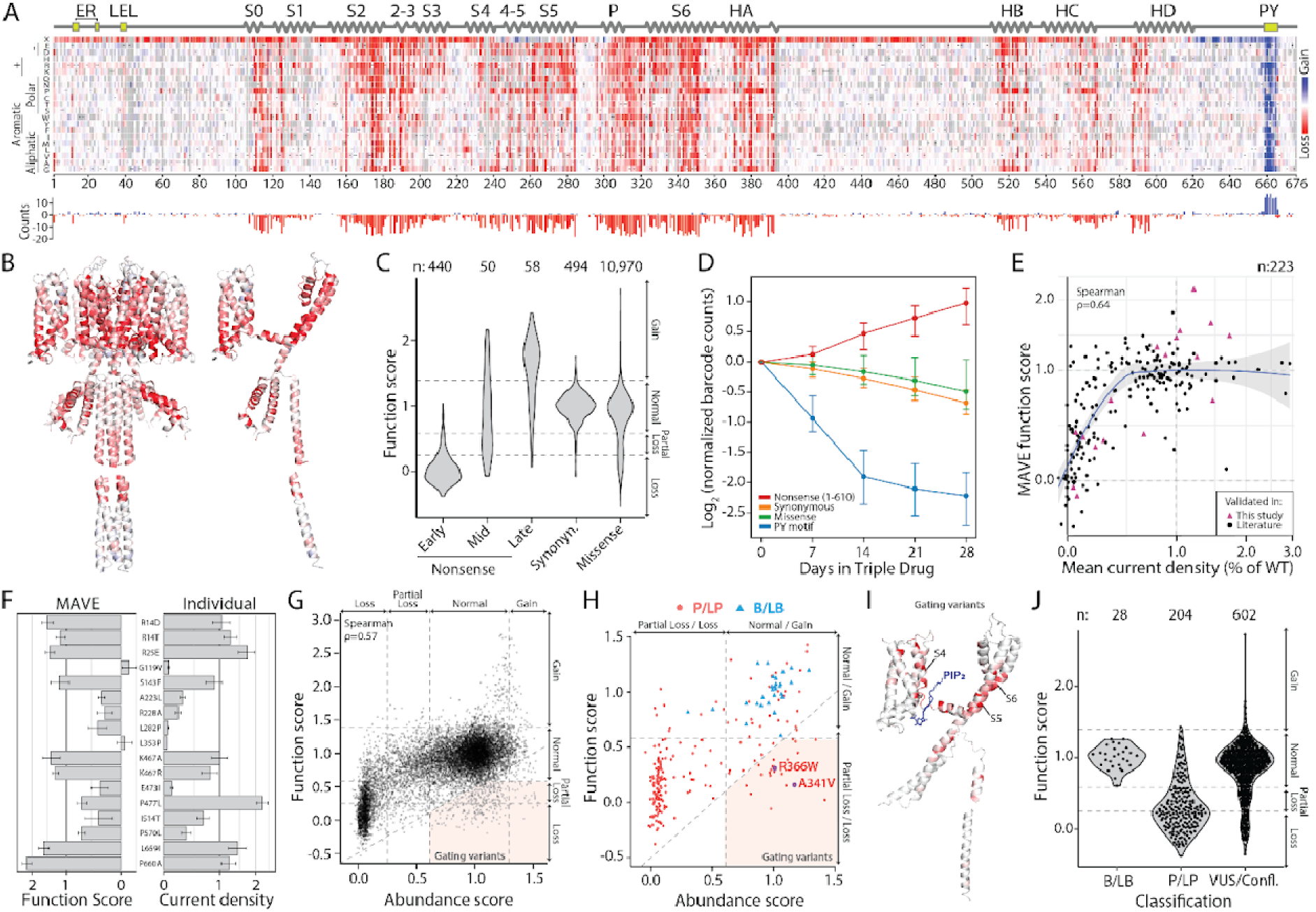
MAVE of K_V_7.1 channel function. Landing pad HEK293 cells expressing a barcoded *KCNQ1* variant library were treated with triple-drug media for 28 days. Samples were collected at days 0, 7, 14, 21, and 28. The dataset was generated from 6 biological replicates. A) Heatmap of function scores (red: low, white: normal, blue: high). WT residues marked by black dashes; bar graph shows increased/decreased surface abundance by position. B) Function scores mapped onto the K_V_7.1 structure (cryo-EM structure 6UZZ).^14^ C) Distribution of day 28 scores by variant class (nonsense: early/mid/late, synonymous, missense). D) Median enrichment/depletion scores over time by mutation type (IQR shown); positional nonsense-variant abundance. E–F) Comparison of MAVE function scores with literature-curated or experimentally evaluated variants. Individual patch clamp measurements performed in this study represent mean and standard error from 18-40 biological replicate cells per variant. E) MAVE scores for all missense variants, with assessed variants from this study shown as purple triangles and previously reported variants shown as black circles (Spearman ρ = 0.61; pseudo-log scale). F) MAVE scores vs. individual current-density assays. G–H) Abundance vs. function: G) all missense variants (ρ = 0.57), H) B/LB vs. P/LP, highlighting gating variants with normal trafficking but low function (e.g., A341V, R366W).^31,39^ I) Gating variants (red) mapped onto the K_V_7.1 structure, clustering near the PIP2 binding site (blue) and pore domain. J) Function scores stratified by clinical classification (B/LB, P/LP, VUS/Conflicting).

Overall, we identified 1,362 loss-of-function, 1,259 partial loss-of-function, and 339 gain-of-function missense variants in the triple-drug function assay (Figure 3F). To assess this dataset, we performed automated patch clamping on a subset of 20 variants and curated patch clamp measurements from an additional 47 published studies (Figure 3E-F, S7, S8). MAVE function scores correlated well with patch clamp data (ρ = 0.64 for current density, n=223 variants). MAVE surface abundance and MAVE function scores were generally correlated (ρ = 0.57); however, 339 variants had near-normal surface abundance but partial to complete loss-of-function (Figure 3G). These variants, which likely affect channel gating, included 22 known P/LP variants, including A341V and R366W (Figure 3H). These variants were enriched in regions critical for channel activation, including the voltage-sensing S4 helix, the S4-S5 linker (which binds the phospholipid PIP_2_, an essential channel activator^16^), and the pore domain (S5, pore helix, and S6; Figure 3I). In the pore domain, 118/129 (91%) of missense variants within 5Å of the central pore axis had partial or complete loss-of-function scores (Figure S9). Function scores matched clinical classifications even better than surface abundance scores: 28/28 (100%) benign/likely benign (B/LB) variants had normal function scores, while 168/204 (82%) pathogenic/likely pathogenic (P/LP) variants exhibited abnormal function scores (Figure 3J, Table S4).

### Heterozygous KCNQ1 MAVEs Reveal Dominant Negative Effects

Given that most LQT1 patients carry *KCNQ1* variants in a heterozygous form, that K_V_7.1 forms tetramers, and that some missense variants are known to exert dominant negative (DN) effects,^23,31^ we next comprehensively assessed *KCNQ1* variants for DN effects. To this end, we engineered HEK293 landing pad cells that stably expressed WT *KCNQ1* with an extracellular hemagglutinin (HA) epitope tag and introduced variant *KCNQ1*-myc variants into the landing pad. We first verified that this system could detect DN effects by showing that two known DN variants (L114P and H126L)^17^ reduced WT *KCNQ1*-HA surface abundance (Figure S10). We then performed multiplexed assays on the *KCNQ1*-myc library to assess surface abundance and function in this heterozygous context (Figure 4A). The surface abundance assay specifically measured abundance of the HA-tagged WT protein, while the function assay reflected the combined activity of the WT and variant channels. There were 2,551 variants with abundance scores below 0.25 in the variant-only assays, and 848 also dramatically reduced WT surface abundance (scores below 0.25) in the heterozygous context, indicating a DN effect (Figure 4B). Nearly all of these DN variants were located in helices in the voltage-sensing domain (n=375), pore domain (n=335), or cytoplasmic domain (n=84; Figure 4B). In contrast, 827 low-abundance variants—particularly early nonsense mutations—did not affect WT-HA abundance, indicating haploinsufficiency, and 876 had an intermediate effect on WT-HA abundance (Figure 4B). In the heterozygous function assay, many variants exhibited loss of function; however, due to higher experimental noise in this experiment, it was more challenging to distinguish between DN and haploinsufficient effects (Figure S11). Heterozygous abundance and function MAVE scores correlated with prior single-variant measurements of heterozygous surface abundance and function (ρ = 0.87 and ρ = 0.47, respectively; Figure S11).^17,23,40,41^

**Figure 4:**
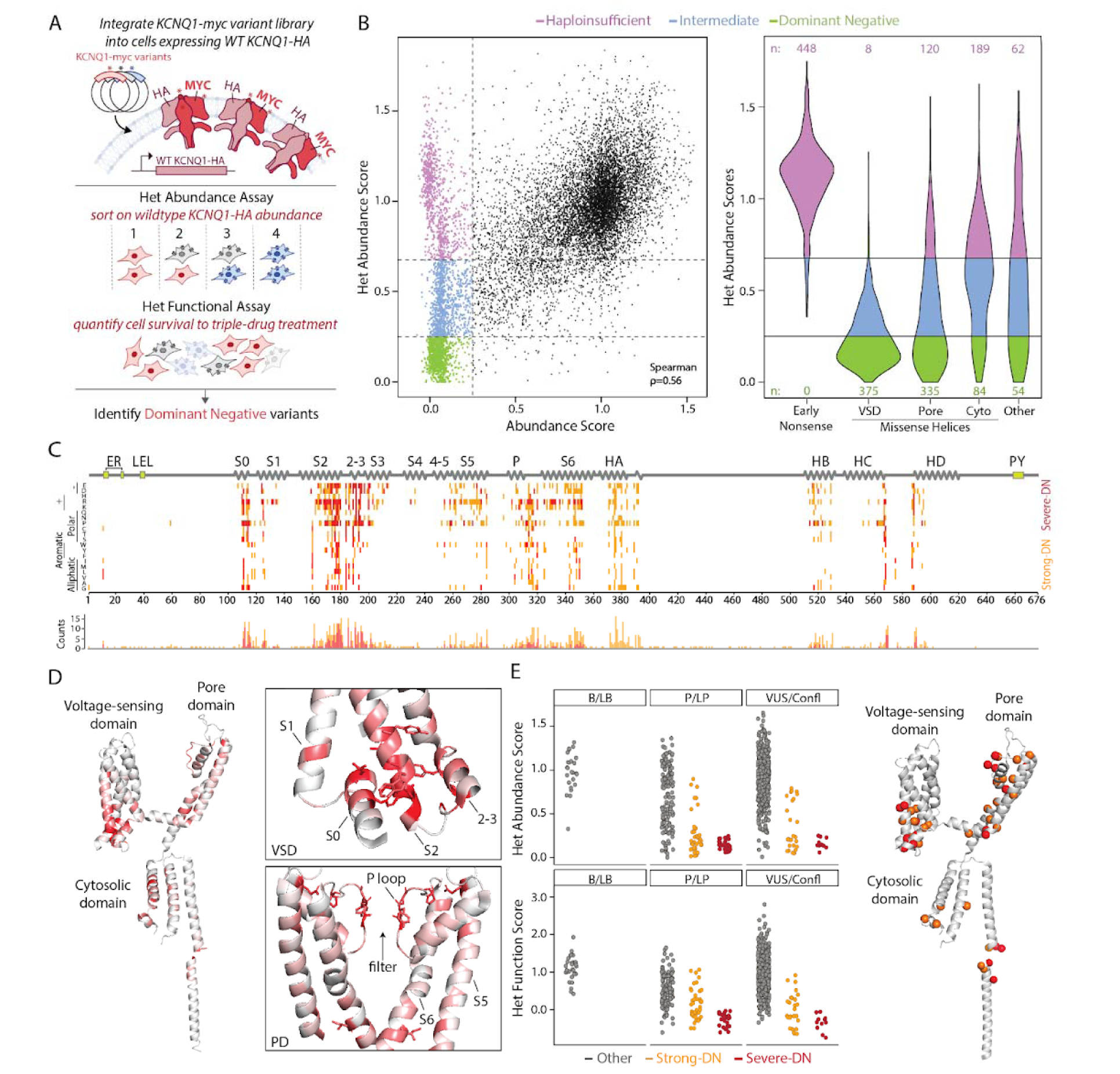
Heterozygous MAVEs identify dominant negative *KCNQ1* variants. A) Heterozygous MAVE experimental workflow. B) Left: Hom vs. Het Abundance Scores. Right: Violin plot of Het Abundance MAVE scores for variants with Variant-Only Abundance <0.25, highlighting dominant negative (green), haploinsufficient (purple), and intermediate (blue) variants by type. C) Heatmap of WT *KCNQ1*-HA surface abundance with *KCNQ1*-myc variants, showing strong (orange) and severe (red) dominant negative effects. D) DN variants mapped onto K_V_7.1 with zoomed-in views of the VSD and Pore domains; residues shown in stick format indicate sites with ≥ 10 DN variants. E) Jitter plot of Het Abundance vs. Het Function MAVE scores, showing strong-DN (orange), and severe-DN (red) variants in B/LB, P/LP, and VUS classes. P/LP variants with DN effects are mapped onto K_V_7.1; strong-DN (orange), severe-DN (red). Structures in panels D-E are derived from cryo-EM structure PDB:6UZZ.^14^ The Heterozygous abundance dataset was generated from 3 biological replicates and the heterozygous function dataset was generated from 15 biological replicates.

Integrating all four MAVE scores (variant-only/heterozygous × abundance/function), we identified 290 “severe DN” missense variants (loss-of-function in all four assays) and 548 “strong DN” variants (loss-of-function in 3 of 4 assays). Of these 838 variants, 94% localized to alpha helices—71% in transmembrane regions and 23% in cytoplasmic regions (Figure 4C). There was a striking hotspot of DN variants in a cytosolic-adjacent region of the voltage-sensing domain comprised of the S0, S2, and S2-3 helices (Figure 4D, Table 1). There were 11 residues in this region that each had at least 10 DN variants. All 11 residues make inter-helix contacts between VSD helices (Figure 4E, Table 1).^17^ The potassium selectivity filter GYGD_314-317_ in the pore loop also contained a large number of DN variants (mean of 11 DN variants/residue); there were also 10 selectivity filter known P/LP variants with DN effects in our assays (Figure 4E-F). Almost all B/LB variants had normal heterozygous abundance and function scores, but 69 known P/LP variants with DN effects were identified (Figure 4F, Table 1). We also identified 38 ClinVar VUS/conflicting variants with DN effects.

**Table 1:** *KCNQ1* dominant negative variants.

| Location | Coordinates | P/LP variants with DN | VUS/Conflicting with DN | Positions with $\geq 10$ DN variants |
| --- | --- | --- | --- | --- |
| S0 | 104-115 | Y**111**C, Y**111**S, L**114**P, E115G | V110E, E115K | 111, 114 |
| S1 | 120-143 | Y125D, Y125N | H126L | * |
| S2 | 150-181 | G168R, T169R, E170K, V173D, R**174**C, R**174**P, W**176**S, A**178**P, G**179**A, G**179**R, G**179**S | E170D, E170G, L175P, W**176**R | 160, 171, 174, 176, 177, 178, 179 |
| 2-3 | 186-194 | G186D, G186V, R190P | G186R, G**189**A, G**189**E, G**189**R, R192P, A194P | 189, 193 |
| S3 | 196-216 | L203P | I200N | * |
| S4 | 224-241 | * | * | * |
| 4-5 | 245-257 | * | * | * |
| S5 | 258-285 | H258P, H258R, R259P, E261K, L262P, L262Q, G269R, L273R, S277W | L262R, L266R, L282P | 284 |
| P helix | 298-311 | * | D301Y | * |
| P loop | 312-321 | G**314**D, G**314**R, G**314**S, Y315H, Y315N, G**316**A, G**316**R, G**316**W, D**317**A, D**317**G | P**320**R | 314, 316, 317, 320 |
| S6 | 322-361 | T322P, G325E, G325R, G325W, A341E, P343A, A344E | G350R, G350V, A352D, A352P, L353P | 346 |
| HA | 363-395 | A371E, A371P, A371T, R**380**S, R**380**W, W392R | I**375**F, Q**376**H, Q**376**R, A378P, W379G, R**380**G, R**380**K, S389Y, T391P | 375, 376, 380 |
| HB | 509-532 | M520R, Y522S | R518P | * |
| HC | 537-567 | S566Y, I**567**T | Q565P | 567 |
| HD | 587-620 | T587M, T587R, I**588**T, G589D, R594P, R594Q |  | 588 |
| Other | * | P117L, Y**184**D, Y**184**H, Y**184**S, G**568**A, G**568**R | R116P, G119V, R195P | 184, 568 |

Blue denotes described P/LP or VUS variants at positions with $\geq 10$ DN variants in our assay. DN=dominant negative (strong or severe).

### Combining Assays and Calibration with Benign and Pathogenic Variants

To better understand variant mechanisms, we next performed principal component analysis (PCA) using the four assay results (variant-only/heterozygous function and abundance scores).^42^ This analysis included 10,316 variants for which all four scores were measured (Figure 5A-C). The PCA analysis successfully distinguished variants by functional class, including early nonsense, synonymous, severe dominant negative, PY motif, and late nonsense variants (Figure 5A), as well as by classification (B/LB vs P/LP; Figure 5B). Unsupervised *k*-means clustering generated subsets that agreed with our functional classifications. We applied *k*-means clustering (n=6 clusters), and named the resulting clusters manually based on the scores of variants in each cluster (Figure S12). We called the six clusters dominant negative, haploinsufficient, partial loss-of-function, normal 1, normal 2, and gain-of-function (Figure S12D). Of the 26 B/LB variants included in the clustering analysis, 24 were classified into one of the two normal clusters. 109 out of 191 P/LP variants were assigned to the dominant negative cluster. In contrast, 26 P/LP variants were categorized into the haploinsufficient cluster, and 14 were grouped into the partial loss-of-function cluster (Figure 5A-C, Table S5).

**Figure 5:**
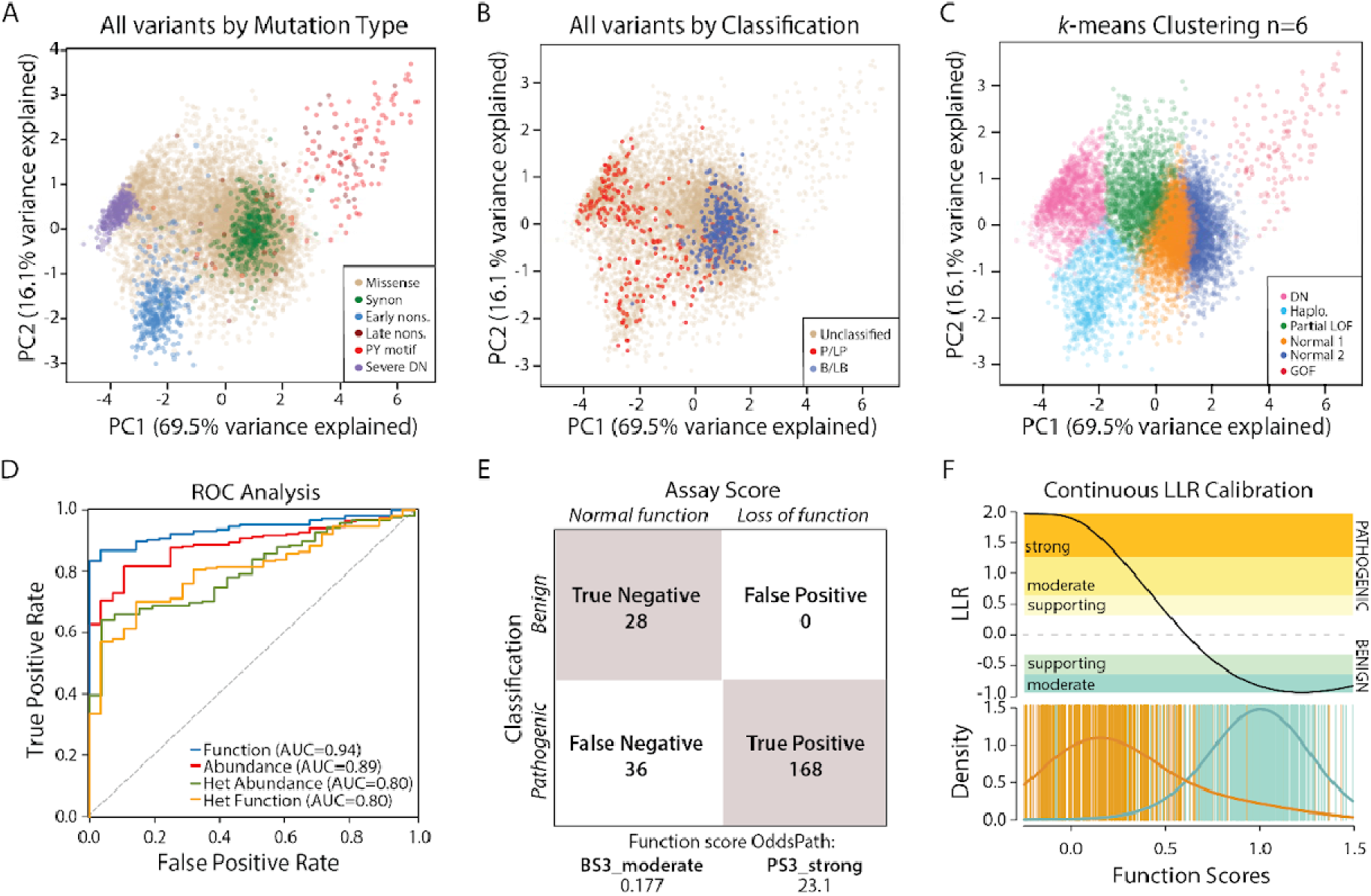
Combinations of assays and calibration with pathogenic and benign variants. A-C) Principal components analysis plotting first two principal components analysis, using the 4 scores for each variant as the input (variant-only and heterozygous function and abundance assays). A) Variants are color coded by category (synon=synonymous, nons.=nonsense, DN=dominant negative). B) Variants are color coded by classification. B/LB=benign/likely benign, P/LP=pathogenic/likely pathogenic. C) Variants were clustered by *k*-means clustering (n=6 clusters). Categories were named manually based on average behavior across the 4 assays. GOF=gain-of-function, Haplo=haploinsufficient, Dom. Neg=Dominant Negative. D) ROC curves for each assay’s ability to predict benign vs pathogenic variants. The assays are single variant and heterozygous (het) MAVE scores of abundance and function, with area under the curve (AUC) values listed in parentheses. E) OddsPath calculation for MAVE function assay. Classification grids show the true positive, true negative, false positive, and false negative rates based on OddsPath predictions and their alignment with known clinical classifications. F) Calibration of MAVE function scores using log-likelihood ratios (LLR). Top: relationship between LLR and MAVE function score. Bottom: density of pathogenic and benign variants across the scoring range.

The predictive power of individual assays in distinguishing P/LP from B/LB variants was evaluated using receiver operating characteristic (ROC) curves (Figure 5D), with area under the curve (AUC) values used to quantify their performance. Among the four assays, the variant-only function assay demonstrated the highest predictive ability with an AUC of 0.94, followed by the variant-only abundance assay with an AUC of 0.89. The heterozygous function and abundance assays showed lower predictive capabilities, each with an AUC of 0.80. We next assessed the performance of three methods that combine data from all four assays—Gaussian naive Bayes, logistic regression, and random forest—using ROC curves. These methods demonstrated similar predictive capabilities (AUC 0.92-0.96), with Gaussian naive Bayes performing best, achieving an AUC of 0.96 (Figure S13). To calibrate the scores for clinical application, we used known B/LB and P/LP variants, following the recommendations of the ClinGen Sequence Variant Interpretation working group (Figure 5E).^26^ This calibration involved calculating “OddsPath” values, which represent the log-likelihood ratio of variant pathogenicity or benignity based on abnormal or normal MAVE scores, using a binary cutoff approach. The function scores aligned most closely with clinical classifications, yielding an OddsPath value of 23.1 for pathogenicity and 0.177 for benignity (Figure 5E, Table S4).^26^ These OddsPath values correspond to PS3_strong and BS3_moderate classification strengths within the ACMG/AMP variant classification framework, further supporting the utility of these assays for clinical variant interpretation. In addition to this primary analysis, we also performed a contingency analysis calibrating each of the MAVE assays with 3 alternate benign and pathogenic truth sets (Figure S14, Table S4). We also calibrated the function scores with a continuous log-likelihood ratio (LLR) approach. This calibration resulted in a continuous range of evidence strengths, ranging from PS3_strong for loss-of-function variants to BS3_moderate for variants with normal function scores (Figure 5F). Applying the continuous LLR framework, variants received the following levels of evidence: 1,757 PS3-strong, 707 PS3-moderate, 333 PS3-supporting, 6,660 BS3-moderate, and 1,477 BS3-supporting. As an example of applying this framework, we analyzed six KCNQ1 VUS from a large published cohort of LQTS cases. Four of these variants demonstrated abnormal function in both the MAVE and automated patch clamp assays, receiving PS3-strong or PS3-moderate evidence (Table S6). In contrast, two variants displayed near-normal function or small functional alterations and were assigned either no evidence or BS3-moderate evidence.

Our assay uses a cDNA form of *KCNQ1* that cannot capture splicing effects. To investigate the potential impact of *KCNQ1* variants on splicing, we used the machine learning algorithm SpliceAI.^43^ Among all synonymous and missense variants resulting from single nucleotide changes, 44 variants exceeded the SpliceAI-recommended cutoff of 0.5, while 75 variants had SpliceAI scores in the suggestive range of 0.2 to 0.5. SpliceAI scores for all variants are presented in File S1, and variants with elevated SpliceAI scores are presented in Table S7.

### MAVE Scores are Associated with QTc Interval and Arrhythmia Risk

We next examined associations between *KCNQ1* MAVE scores and electrocardiographic and arrhythmia phenotypes in 711,448 participants across three large biobanks: All of Us, BioVU, and UK Biobank. The distribution of MAVE scores for all assayed variants spanned a wide range of functional effects, but variants with abnormal function were relatively depleted in biobank participants (Figure S15). Consistent with this, 35/36 variants with an allele frequency >3×10^-5^ in gnomAD had normal MAVE scores. Variants with an allele frequency <3×10^-5^ or absent from gnomAD had a mix of normal and abnormal MAVE scores (Figure 6A). Recognizing that many patients with P/LP variants and phenotypes may not carry the LQTS label, we looked at rates of LQTS International Classification of Disease (ICD) code in biobank participants. The fraction of participants carrying an LQTS ICD code increased with lower MAVE function scores, reaching 8.1% for loss-of-function variants (Figure 6B). Among 155,401 biobank participants with available electrocardiograms, we observed a spectrum of QTc values based on the *KCNQ1* function score (Figure 6B). Loss-of-function variant carriers exhibited a mean QTc prolongation of 25.9 ms (456.7 ms, 95% CI 450.0–463.3) compared to normal-function variant carriers (430.9 ms, 95% CI 429.7–431.9). In conventional “thorough QT” studies of drug effects, a 10 ms prolongation is enough to generate increased regulatory scrutiny.^44^

**Figure 6:**
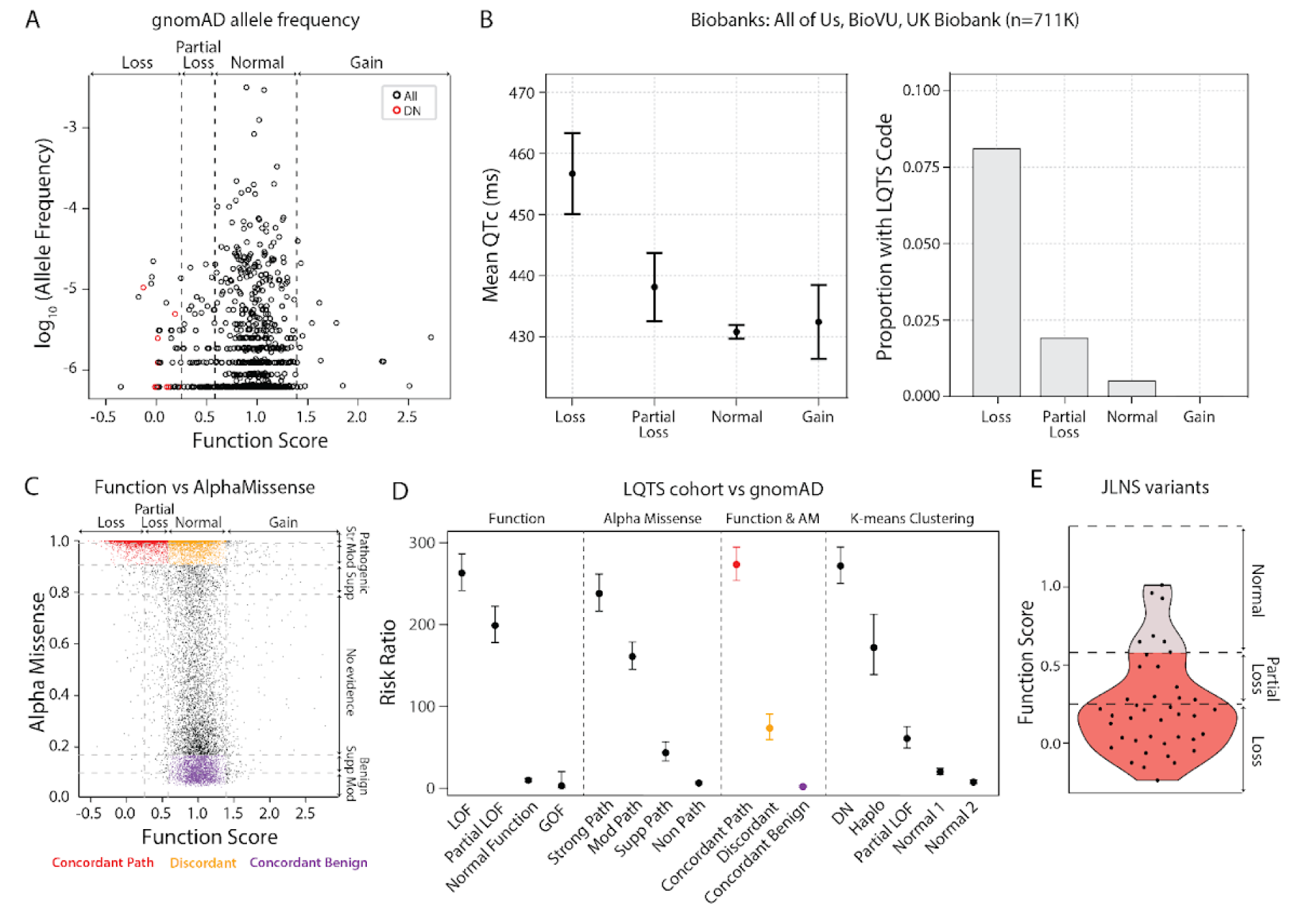
MAVE scores stratify *KCNQ1* variants and are associated with arrhythmia risk. A) Population allele frequency (gnomAD, logLJLJ) versus MAVE function score. DN variants are highlighted in red. B) Clinical associations of *KCNQ1* variant functional classes across large biobanks (BioVU, UK Biobank, All of Us; n=711K). Left: QTc intervals for carriers in each class, shown as mean ± 95% CI. Right: proportion of individuals with an LQTS diagnostic code by functional class (LOF, partial LOF, normal, GOF). C) Experimental functiol scores plotted against AlphaMissense pathogenicity probabilities shows concordance for many variants (ρ = - 0.49) but identifies a set of discordant variants with normal function scores and deleterious AlphaMissense predictions. D) Risk ratios (±95% CI) for variant classes in a LQTS cohort (n=1,847) versus gnomAD controls. Left to right: MAVE-only classes, AlphaMissense-only classes, combined classes, and *k*-means clustering classes. Numbers above points are unique variant counts. E) *KCNQ1* variants identified in at least one JLNS patient in the literature predominantly fall within the loss-of-function range.

MAVE scores were concordant with computational predictors (Figure S16), including AlphaMissense (Spearman ρ = -0.49; Figure 6C), but a subset of discordant variants retained normal function in MAVE despite being classified as pathogenic by AlphaMissense and other predictors. In a published cohort of 1,847 Long QT syndrome cases^4^, variant classes defined by MAVE scores alone demonstrated significantly elevated risk ratios (RR) compared to gnomAD controls, including an RR of 263 (95% CI 241-286) for loss-of-function variants (Figure 6D, Table S8). Computational predictor-defined classes also showed enrichment for Long-QT cases (Figure S17, Table S8), including an RR of 238 (216-262) for variants with strong pathogenic AlphaMissense scores (Figure 6D). Variants with concordant predictions of pathogenicity (MAVE loss or partial loss-of-function and AlphaMissense strong pathogenic) yielded an RR of 273 (254-294), while variants with concordant predictions of benignity (MAVE normal function and AlphaMissense benign) had a low RR of 1.8 (1.1-3.1). Discordant variants (MAVE normal function but AlphaMissense strong pathogenic) had a RR of 73.3 (59.4-90.5; 3.7-fold lower than concordant pathogenic variants), suggesting that while these variants collectively retain some arrhythmia risk, AlphaMissense overcalls *KCNQ1* variant pathogenicity or identifies variants with smaller-effect or lower-penetrance. When variants were grouped by *k*-means cluster, RRs showed a clear gradient (Figure 6D). Dominant-negative variants exhibited the strongest association with LQTS (RR = 272, 95% CI 250–295), followed by haploinsufficient (RR = 172, 139–213) and partial loss-of-function variants (RR = 60.7, 49.0–75.1), whereas the Normal 1 and Normal 2 clusters showed lower LQTS risk (RR = 20.0 and 7.3, 95% CI 16.6–24.1 and 5.3–10.2, respectively). As no variants in the gain-of-function cluster were identified among LQT cases, a risk ratio could not be calculated. Variants linked to JLNS (homozygous or compound heterozygous *KCNQ1* variants linked to QT prolongation and deafness) in the literature clustered within the loss-of-function range, with 83% (34/41) showing partial loss or loss-of-function in the MAVE assay (Figure 6E). Overall, these findings demonstrate that MAVE scores effectively stratify *KCNQ1* variants, predict arrhythmia risk, and outperform current computational tools in clinical contexts.

## Discussion

In this study, we comprehensively characterized 13,403 *KCNQ1* variants using four multiplexed assays to measure their effects on surface abundance and channel function in both variant-only and heterozygous conditions. The complementary data obtained from the four assays allowed us to dissect the mechanisms underlying the pathogenicity of *KCNQ1* variants, and structure-function analyses allowed us to decipher the structural rationale for key mutation-intolerant regions. Our comprehensive, protein-wide functional approach offers valuable new insights into the *KCNQ1* functional landscape and highlights the strong relationship between assay-based functional abnormalities and clinical arrhythmia risk.

Since it is activated at depolarized potentials, K_V_7.1 does not exhibit significant current at baseline and has a minimal fitness defect. We therefore developed a novel K_V_7.1 functional assay that used two activator drugs and a sensitizing drug, ouabain. This approach is analogous to the “triple-drug” treatment we previously used for an *SCN5A* selection assay,^45^ and could be a general approach for functional assays of ion channels.

Previous studies of large LQT1 cohorts have identified regional hotspots within the gene, particularly in the transmembrane domain (notably the pore region) and in conserved C-terminal regions corresponding to cytoplasmic helices.^4,46–49^ This study expands on those findings by providing enhanced resolution at the single-residue level. Our study validated previously identified motifs as having strong mutational effects, including those involved in forward trafficking (LEL),^32^ calmodulin binding (IQ and 1-5-10)^34^, and ubiquitylation (PY).^38^ Additionally, we discovered novel motifs and mutational patterns that would be challenging to detect through small-scale mutational studies. Variants that altered the newly described RR and RKR ER retention motifs caused a striking increase in cell-surface abundance. However, these variants exhibited only mild gain-of-function effects, as determined by the MAVE function assay and validated through patch clamp experiments. Variants altering the electric charge of the unstructured N-terminus and the 112-amino-acid linker between HA and HB influenced channel abundance. Strong electrostatic potentials across the channel have been previously noted.^33^ Although the detailed mechanism of the mutational charge sensitivity remains unclear, net charge in these regions may influence protein folding, membrane insertion during translation, ER retention, or protein-protein interactions.

Our maps also offer key insights into K_V_7.1 assembly and multimerization. Our MAVEs of *KCNQ1* (this study) and *KCNE1*^50^ show that disrupting K_V_7.1-calmodulin binding reduces surface abundance and function, highlighting calmodulin’s critical role in channel assembly.^51^ The *KCNQX* channels contain a C-terminal “assembly” domain with two tetrameric coiled-coils (HC and HD). Our maps reveal the periodic 3/7 hydrophobicity pattern required for coiled-coil formation, along with three charged residues (R_591_, R_594_, and E_596_) in the HD helix that form inter-helix salt bridges and show high mutational sensitivity. The 8 reported P/LP variants in these 3 residues all exhibit loss-of-abundance and function. Crystal structures of the HD helix show both trimeric^36^ and tetrameric^52^ coiled-coil states, with R_591_-E_596_ stabilizing the intermediate trimeric state and R_594_-E_596_ stabilizing the tetrameric state.^36^ Thus, this study advances understanding of coiled-coil sequence determinants, building on prior MAVE research of dimeric coiled coils.^53^

By comparing our surface abundance and function assays^54^, we were able to clarify the mechanisms underlying loss-of-function *KCNQ1* variants. The two MAVE assays showed strong correlation, with most loss-of-function variants also impairing cell surface abundance. This alignment highlights a common theme among transmembrane proteins, where loss-of-function variants are frequently associated with surface abundance or trafficking defects.^55^ However, we also identified 339 variants with near-normal surface abundance but loss-of-function, suggesting disruptions in channel gating mechanisms. Notably, this subset included 22 known pathogenic/likely pathogenic (P/LP) variants.

MAVE studies typically evaluate variants expressed individually, often using the HAP1 haploid cell line for saturation editing experiments.^56^ One notable exception is a *TP53* MAVE that assessed variants in both variant-only and heterozygous states, identifying those with DN effects.^57^ In our study, heterozygous MAVEs identified 838 *KCNQ1* missense variants with DN effects, including 68 of 218 known P/LP variants (31%). *k*-means clustering analysis revealed an even higher proportion of P/LP variants in the DN cluster (100 of 179 assignable variants, 56%). These DN variants were enriched in key regions, including the selectivity filter and a hotspot in the voltage-sensing domain (VSD) critical for VSD folding.^31^ This domain does not participate in the monomer-monomer interactions essential for multimerization, unlike the pore domain and intracellular helical regions. Consequently, variants that disrupt the VSD may still fold and assemble into mutant:wildtype multimers, potentially explaining their high burden of dominant-negative effects. DN variants may contribute to a higher penetrance form of LQT1; in our case-control analysis, DN variants were associated with higher risk ratios for LQTS compared to haploinsufficient or non-DN variants. DN variants are promising targets for allele-specific therapies.^58,59^ Overall, our findings offer a detailed understanding of the global landscape of dominant-negative effects, confirming prior research on specific *KCNQ1* variants in these regions,^12,17^ and uncovering novel biological insights.

Most *KCNQ1* nonsense variants exhibited loss-of-function effects, consistent with their clinical associations. For instance, R518X, R594X, and Q604X all showed loss-of-function scores, matching literature reports of LQTS or JLNS.^60,61,62^ In contrast, several late nonsense variants (residues 611+) that truncate the C-terminal PY motif displayed gain-of-function. Notably, Y662X, the most frequent *KCNQ1* nonsense variant in gnomAD, showed gain-of-function, consistent with its detection in several patients with early-onset atrial fibrillation, syncope, or sudden death.^40,63^ Located in the last exon and only 42 bp from the native stop codon, Y662X may escape nonsense-mediated decay. These functional data and patient reports may warrant reevaluation of Y662X’s classification, which currently ranges from VUS to benign in ClinVar. The *KCNQ1* gain-of-function truncations are analogous to gain-of-function PY motif deletions in the epithelial sodium channel (ENaC) that can cause Liddle syndrome.^64^ We also unexpectedly identified mid-nonsense variants (residues 239–307) with elevated function scores. Although these truncations remove the pore domain, expression of S253X produced K_V_7.1-like currents, possibly due to a nonsense readthrough mechanism. However, this effect was plasmid-dependent and absent in alternative constructs, suggesting limited physiological relevance. These mid-nonsense variants also likely undergo nonsense-mediated decay *in vivo*.

Determining assay performance is a critical step in ensuring the reliability and clinical utility of high-throughput functional assays. Our assay demonstrated robust predictive performance, achieving an area under the receiver operating characteristic curve of 0.94 for the variant-only function assay. While the other three MAVE datasets provide valuable insights into variant mechanisms, the variant-only function assay demonstrated the highest accuracy for clinical classification. We further calibrated the function scores using both binary (OddsPath) and continuous (Log Likelihood Ratio) approaches. The binary approach calculates OddsPath values based on score thresholds, simplifying classification decisions and aligning well with ACMG guidelines. Meanwhile, the continuous log-likelihood ratio (LLR) approach provides a nuanced spectrum of evidence strengths, accommodating the diversity of variant effects observed in functional assays. Both calibration approaches provided up to strong evidence toward pathogenicity (PS3_strong) and up to moderate evidence toward benignity (BS3_moderate). Applying the continuous LLR framework, we provide functional evidence towards pathogenicity or benignity for 10,934 variants, including 1,757 variants with PS3-strong and 6,660 variants with BS3-moderate evidence. We also performed a series of contingency analyses with alternate sets of truth variants, that mostly yielded similar results. Given the substantial number of *KCNQ1* variants supported by additional MAVE functional evidence, we expect that our scores will contribute significantly to addressing the issue of VUS in this gene.

The observed correlation between *KCNQ1* MAVE function scores and clinical phenotypes highlights the potential of MAVE as a tool for refining variant interpretation and assessing arrhythmia risk. We validated the clinical impact of variants with abnormal MAVE scores using four approaches. First, we demonstrated that variants with abnormal scores were underrepresented in the general population and biobank datasets. Second, loss-of-function variants showed strong associations with LQTS diagnoses in a case-control analysis. Third, abnormal function variants were associated with significant QTc prolongation and LQTS diagnoses across three large biobanks. Fourth, almost all *KCNQ1* variants detected in cases of JLNS had abnormal function in our assays. Computational predictors are increasingly being used to predict variant effects. Our analyses show high concordance between our MAVE scores and predictors including AlphaMissense. However, a case-control analysis of discordant scores suggests that the current computational predictors are overcalling pathogenicity in *KCNQ1*, or identifying variants that have lower penetrance. Overall, these analyses reinforce the clinical relevance of the function scores. Our MAVE scores provide a more nuanced stratification of variant risk, offering valuable insights into genotype-phenotype relationships, and improve the accuracy of clinical classifications compared to existing methods.

Our comprehensive maps offer a valuable dataset for future studies of specific *KCNQ1* features and variants, including channel features that could be modulated for therapeutic use. One example is a therapeutic avenue that seeks to prevent the ubiquitination-mediated degradation of *KCNQ1*.^65^ Another direction of future research could focus on integrating high-throughput function data with clinical practice to enhance patient diagnosis and risk stratification. This integration could pave the way for more personalized diagnosis and intervention for patients with *KCNQ1* variants and arrhythmia risk. Our datasets contribute to the growing body of MAVE studies of arrhythmia-related genes, including *CALM*, *KCNE1*, *KCNH2*, *KCNJ2*, and *SCN5A.*^45,50,66–68^ *KCNQ1* is one of five homologous *KCNQ* channels (*KCNQ1–KCNQ5*) and belongs to a broader family of voltage-gated cation channels, many of which are linked to clinical phenotypes. Two exciting future directions are to extend MAVE studies to these genes, and to use MAVE datasets to refine computational tools to predict variant function. Overall, our findings demonstrate that multiplexed assays can transform variant interpretation, sharpen disease-risk prediction, and accelerate the realization of genomic medicine.

### Limitations of the study

One limitation of our study is the inability to capture splice variants, which can be addressed using complementary high-throughput splicing assays^69^ or endogenous gene editing approaches.^70^ Additionally, because *KCNE1* co-expression occludes labeling of the *KCNQ1* myc tag and prevents binding of the *KCNQ1* activator compounds,^71^ we did not incorporate *KCNE1* into our MAVE abundance or function assays. This could potentially lead to discordant results in some variant pathogenicity classifications, although it has been shown that coexpression of *KCNQ1* with *KCNE1* generally results in only mild changes in *KCNQ1* cell surface abundance.^17^ The MAVE experiments were performed in HEK293 cells that may not express the full suite of *KCNQ1*-interacting or regulating proteins. We also did not model certain features of *KCNQ1* biology, such as adrenergic stimulation.

## Materials and Methods

### Overview of Multiplexed Assays of Variant Effect (MAVE) experimental approach

A myc epitope tag was inserted into the extracellular region of *KCNQ1* to facilitate the measurement of protein surface abundance. Saturation mutagenesis was performed to generate a comprehensive plasmid library of 13,706 single amino acid *KCNQ1* variants (obtaining 97% of 14,196 possible nonsense, synonymous, and missense variants). Random 18-bp barcodes were cloned into the plasmid library, resulting in many barcodes corresponding to each variant. Deep sequencing of the plasmid pool established correlations between the barcodes and their respective variants. HEK293 landing pad (LP) cells were transfected with the *KCNQ1-*myc variant library, which integrated into the landing pad (1 plasmid/cell). Surface abundance was evaluated by staining live cells with a fluorophore-conjugated anti-myc antibody, followed by fluorescence activated cell sorting (FACS) into 4 bins based on fluorescence intensity and deep sequencing for quantification. Channel function was assessed by treating the cells with triple-drug media over a 28-day period, with cell pellets collected every 7 days for deep sequencing and DNA quantification. In heterozygous studies, HEK293 LP cells expressing an HA-tagged copy of WT *KCNQ1* were transfected with the *KCNQ1* variant library, and cells were assessed for channel function or WT-KCNQ1-HA surface abundance as described above.

### KCNQ1 library generation

Each variant of the *KCNQ1* variant library consists of two key elements: a surface-exposed myc-tag epitope and an 18-base barcode. The myc-tag epitope facilitates the staining of cells expressing K_V_7.1 at the cell surface and the barcode enables the identification and quantification of each variant. The myc-tag epitope (5’-GAGCAAAAACTGATCTCAGAAGAGGATCTT-3’) was cloned into *KCNQ1* between residues 146 and 147, positioned within the extracellular loop that connects the S1 and S2 transmembrane domains. This tag was previously used to measure surface abundance of *KCNQ1* in non-multiplexed experiments.^17,27^ We next created a *KCNQ1* variant library using myc-tagged *KCNQ1.* We divided the *KCNQ1* cDNA sequence into three zones (residues 1-236, 236-394, and 394-676) using existing restriction sites that allowed modular manipulation and subcloning of each zone. A list of restriction enzyme sites separating each of the three zones is presented in Table S9. The *KCNQ1* variant library was generated using saturation plasmid mutagenesis, a method that employs inverse PCR on a compact template plasmid containing just a single zone of the gene.^72^ Individual mutagenesis reactions were performed for each amino acid position. For each reaction, the forward primer contained a degenerate NNK sequence. Mutagenesis with this primer and a corresponding reverse primer introduced each of 20 possible amino acids and a stop codon. The PCR products for each zone were pooled. The pools were PCR-purified (QIAGEN), phosphorylated with T4 PNK (NEB), and self-ligated using T4 ligase (NEB). The pooled product was PCR-purified again and electroporated into ElectroMAX DH10B cells (ThermoFisher) using a Gene Pulser Electroporator (Bio-Rad; 2.0 kV, 25 µF, 200 Ω). Dilutions of the electroporated cells were plated on Luria-Broth agar plates supplemented with ampicillin to determine transformation efficiency. The library was purified using a MaxiPrep kit (Qiagen) and subcloned into a promoterless AttB pIRES:mCherry-BlastR plasmid^45,73^ using restriction digestion with appropriate restriction enzymes (Table S9) and T4 ligase (NEB). The library was then electroporated into ElectroMax DH10B cells and purified as above.

We next tagged the three variant zone plasmid libraries with random 18mer barcodes. The barcodes were generated by annealing a pair of primers specific to each zone. The forward primer sequence included spacer-BsiWI-RE2-N18-RE1-XbaI-spacer, where RE1 and RE2 denote the left and right restriction enzyme cut sites flanking the zone, BsiWI and XbaI represent specific restriction enzyme recognition sites, N18 denotes a sequence of 18 nucleotides (N’s), and spacer refers to a 4-nucleotide spacer sequence. The reverse primers were the reverse complement of the 3’ end of the forward primers. Lists of all primer sequences used in this study are presented in Table S10 and Table S11. These primers were annealed by incubating at 95°C for 3 minutes and cooling to 4°C by 1° per 10s. The annealed primers were extended to fully double-stranded DNA using Klenow polymerase (NEB). The double stranded barcode was purified by phenol-chloroform extraction, digested with BsiWI and XbaI restriction enzymes (ThermoFisher), and re-purified by phenol-chloroform extraction. The resulting DNA was ligated into the *KCNQ1*-myc plasmid library using T4 ligase (NEB) and electroporated into ElectroMax DH10B cells.

### Subassembly to determine the barcode-mutation relationship

Next, we performed “subassembly” by generating Illumina sequencing libraries to establish the relationship between barcodes and *KCNQ1* mutations. Given the considerable distance between the barcode and the mutation sites in zones 2 and 3, these zones were digested with restriction enzymes and self-ligated to bring the barcode closer to the mutant region. A series of PCR reactions using Illumina-compatible primers was then performed to amplify products containing the barcode on one end and the mutant region on the other (Table S12). These libraries were deep sequenced using the Illumina NovaSeq platform.

The following computational pipeline for subassembly analysis was implemented. First, we extracted barcodes and trimmed primer sequences from the raw fastq files using custom Python scripts. Barcode segments were identified using a constant 5-6 base prefix and suffix sequence that preceded or succeeded the barcode. Barcode frequencies were plotted in R, producing a bimodal distribution that distinguished high-frequency barcodes from low-frequency barcodes or barcodes with sequencing errors. We selected a minimum barcode frequency threshold based on the trough between the two modes and filtered out barcodes below that cutoff (Figure S18). Next, for cases where two barcodes were within two single-nucleotide mutations of each other, the lower frequency barcode was removed using a Python script. This removed low-frequency barcodes that were sequencing errors of genuine high-frequency barcodes. Reads corresponding to each retained barcode were sorted, barcodes were trimmed, and the reads were aligned to the WT *KCNQ1* plasmid sequence using *bwa.*^74^ Consensus sequences for each barcode were calculated using *samtools mpileup*^75^, and variant calls were transformed into a table format using R. Barcodes with single codon mutations causing missense, nonsense, or synonymous variants were retained for further analysis. Barcodes with fully WT sequence, harboring multiple mutations, or having low coverage with ambiguous genotypes were excluded from the dataset.^75^ A single barcode coincidentally present in libraries from two separate zones was removed. The mutation coverage of resulting amino acid changes at each position across all three zones is depicted in Figure S2.

### Cell lines used in this study

Experiments were conducted using either LP cells or LP-KCNQ1-HA cells (LP cells expressing a stably integrated HA-tagged copy of WT *KCNQ1*), which were used for heterozygous experiments. LP cells feature a doxycycline-inducible promoter upstream of an AttP integration landing pad site, a Blue Fluorescent Protein (BFP) sequence, and an iCasp caspase sequence.^29^ The AttB:mCherry:BlastR plasmid can integrate into the AttP landing pad. Integration of the AttB:mCherry:BlastR plasmid into the AttP landing pad induces a switch in gene expression, producing mCherry and Blasticidin resistance genes in place of BFP and iCasp. This dual-purpose switch employs mCherry as a marker of integration and Blasticidin resistance as a negative selection marker for successfully integrated cells. Unless otherwise mentioned, all cells were maintained at 37°C with 5% CO_2_ in HEK media, consisting of DMEM supplemented with 10% FBS (Corning), 1% non-essential amino acids (Corning), and 1% penicillin/streptomycin (Corning).

To create the LP-KCNQ1-HA cell line for heterozygous experiments, we cloned an HA-tagged copy of WT *KCNQ1* into a Sleeping Beauty transposon system, specifically pSBbi-GN (a gift from Eric Kowarz, Addgene #60517), using SfiI restriction digestion (NEB) to create pSBbi-GN::*KCNQ1*. This plasmid features constitutively active bidirectional promoters responsible for the expression of both *KCNQ1* and *EGFP*. This plasmid was randomly integrated into a non-landing pad site in LP cells. On day 0, the LP cell line was transfected with pSBbi-GN::*KCNQ1* and a plasmid expressing the Sleeping Beauty transposase, pCMV(CAT)T7-SB100 (a gift from Zsuzsanna Izsvak, Addgene #34879),^76^ using FuGENE6 (Promega) in a 1:3 ratio with DMEM (Thermo). On day 6, the cells were exposed to HEK media supplemented with 250µg/mL doxycycline (Sigma) to induce expression of the BFP/iCasp from the landing pad. The cells were sorted with a BD FACSAria IIIU (BD Biosciences), using a 100µm nozzle for cells expressing high levels of BFP and high levels of GFP, and plated onto amine-coated plates (Corning). Single-cell colonies grown in HEK media supplemented with 250µg/mL doxycycline for 48 hours were screened on a BD LSRFortessa SORP (BD Biosciences) to identify a colony with high rates of both landing pad (BFP) and *KCNQ1* (GFP) expression. The resulting landing pad cell line expressing WT *KCNQ1* is denoted as LP-KCNQ1-HA.

### Integration of the library into landing pad cells

HEK293 cells (LP or LP-KCNQ1-HA) were cultured in 10-cm dishes in HEK media until reaching approximately 70% confluence. Each dish was transfected with 15-20 µg of the *KCNQ1* variant library and 5 µg of the BxbI recombinase plasmid (a gift of Pawel Pelczar, Addgene #51271^77^) using either Lipofectamine 2000 (Invitrogen) or Lipofectamine 3000 (Invitrogen). Cells were washed with HEK media 5-6 hours post-transfection. The following day, cells were passaged using Accutase (Sigma) into three 10-cm dishes and fed with 10 mL dox media (HEK media supplemented with 250µg/mL doxycycline [Sigma]) to induce expression of the landing pad. Alternatively, for some replicate experiments, cells were grown in larger T-175 flasks, and each flask was transfected with 60 µg of the *KCNQ1* variant library and 5 µg of the Bxb1 recombinase plasmid. We refer to this method of selecting integrated cells as the “Dox-Blast-AP selection".

Upon successful integration of a single AttB-containing plasmid, the landing pad expresses only one *KCNQ1-*myc variant, mCherry, and BlastR. After 24 hours, cells were passaged in place using 1 mL Accutase, quenched with 9 mL selection media (HEK media supplemented with 250 µg/mL doxycycline (Sigma), 100 µg/mL blasticidin (Sigma), and 10 nM AP1903 [MedChemExpress]). Addition of blasticidin and AP1903 selected for successfully integrated cells that expressed *KCNQ1-*myc library:pIRES:mCherry:BlastR, and against non-integrated cells that continued to express BFP/iCasp9 caspase. Analytical flow cytometry was performed 7 days post-transfection to verify integration efficiency, and cell sorting took place 8-9 days post-transfection.

### MAVE assays of KCNQ1 surface abundance

Separate experiments were conducted for variant-only and heterozygous studies to evaluate the differences in surface abundance of *KCNQ1* variants alone and when co-expressed with WT KCNQ1-HA. The surface abundance of the myc-tagged *KCNQ1* variants and HA-tagged WT *KCNQ1* was quantified using fluorescence-activated cell sorting (FACS) combined with deep sequencing. For replicates 1-3 of the variant-only abundance experiments, *KCNQ1* zones 1, 2, and 3 were independently transfected into separate batches of cells and separately sorted and sequenced. For replicates 4-5 and all other abundance and function experiments, *KCNQ1* zones 1, 2, and 3 were pooled and transfected into 1 batch of cells per replicate.

The variant-only abundance experiments began with integrating the *KCNQ1* variant library into the landing pad site of LP cells, following Dox-Blast-AP selection.^50^ Seven days post-transfection, integration efficiency was quantified using flow cytometry. Specifically, the cells were washed with 1 mL of dPBS, dissociated with 200 µL Accutase at 37**°**C for 3 minutes, and then quenched with 800 µL HEK selection media. After centrifugation at 300g for 5 minutes, the cells were resuspended in 500 µL of calcium- and magnesium-free dPBS, and the transfection efficiency was assessed by comparing the relative expression of mCherry and BFP. Cells achieving over 80% transfection efficiency were selected for FACS. These cells, grown in 10-cm plates, were washed with 5 mL of calcium- and magnesium-free dPBS, dissociated with 1 mL Accutase at 37**°**C for 3 minutes, and quenched with 2 mL of HEK media. Two 10-cm plates were pooled per biological replicate, amounting to approximately 20 million cells. After centrifugation at 300g for 5 minutes, the cells were washed with 5 mL of blocking solution (dPBS with 1% bovine serum albumin) and stained with a 1:500 dilution of Myc-Tag (9B11) Mouse mAb (Alexa Fluor 647 Conjugate, Cell Signaling #2233) in blocking solution. The cells were incubated with shaking for 30 minutes at room temperature in the dark. After three additional washes with blocking solution, the cells were resuspended in 3 mL of blocking solution. Sorting was performed using BD FACSAria IIIU (BD Biosciences) equipped with a 100 µm nozzle. Compensation was applied using cells expressing single fluorophores for mCherry or BFP, and myc-tagged beads stained with AF647 (Cell Signaling), and single cells were identified based on forward and side scatter. BFP-/mCherry+ single cells were sorted into four bins based on AF647 staining intensity, with approximately 25% of the cells in each bin. At least 800,000 cells per bin were sorted into amine-coated plates (Fisher Scientific) containing HEK media. Cells were grown for 2-7 days post-sort until they reached one confluent 10 cm plate/sample, whereupon cell pellets were frozen at -20°C in preparation for Illumina sequencing (see “Post-assay barcode sequencing and score calculation” below).

We next assessed the LP-KCNQ1-HA system as a platform for studying the dominant negative effects of *KCNQ1* variants in a heterozygous context. We integrated a panel of five known variants from the literature into LP and LP-KCNQ1-HA cells, and measured surface abundance of the variants using flow cytometry with anti-myc or anti-HA antibodies under variant-only or heterozygous conditions, respectively. A similar approach was used to label surface expression, except with reduced volumes, followed by analytical flow cytometry on a BD 5-laser Fortessa instrument. Variant-only cells were labeled with an anti-myc antibody and heterozygous cells were labeled with an anti-HA antibody, each conjugated to Alexa Fluor 647 (Cell Signaling #2233 and #3444, respectively).

The heterozygous surface abundance MAVE assay was then adapted from the variant-only assay, with key modifications. The *KCNQ1* variant library was integrated into LP-KCNQ1-HA cells and transfection efficiency and FACS analysis were performed as described above for the variant-only abundance experiments, with the modification of staining cells with an HA-Tag (6E2) Mouse mAb (Alexa Fluor 647 Conjugate, Cell Signaling #3444), which specifically labels the WT HA-tagged allele. This approach allowed for the detection of dominant negative effects caused by mutant alleles on the surface expression of the WT *KCNQ1*.

### Development of a KCNQ1 function assay

LP cells were transfected with a 2:1 ratio of plasmids: 6.66 µg of WT *KCNQ1* and 3.33 µg of an empty vector. Both constructs express an mCherry fluorophore, with the WT *KCNQ1* plasmid exhibiting higher fluorescence intensity than the empty plasmid, enabling differentiation of cells carrying each plasmid. To facilitate integration into the landing pad, 1.8 µg of the BxbI recombinase plasmid (a gift from Pawel Pelczar, Addgene #5127114) was co-transfected using Lipofectamine 3000 (Invitrogen). Following passages in dox and selection media, as previously described, cells were treated with various drug conditions for 28 days. These conditions included a no-drug control, double drug media (selection media supplemented with 5uM each of the channel activators ML277 and R-L3)^78,79^, and triple-drug media (selection media supplemented with 5uM ML277, 5uM R-L3, and 10 nM ouabain). ML277 and ouabain were obtained from Tocris, and R-L3 (full name: L-364,373) was obtained from MedChemExpress. The media was changed three times per week. At seven-day intervals, starting from day 0, the ratio of plasmid abundance was quantified using analytical flow cytometry. The competitive index of WT *KCNQ1* at time point y was determined by comparing the changes in the abundance of cells expressing WT *KCNQ1* to cells expressing an empty plasmid. This comparison was quantified using analytical flow cytometry in two independent replicates. The formula used was 100*(A_WT, y_ / A_WT, day 0_) / (A_empty, y_ / A_empty, day 0_), where A_x, y_ represents the abundance of sample x at timepoint y. The competitive index for each condition/time point was averaged across the two replicate samples, and the mean and standard error of the mean were plotted.

### MAVE assays of KCNQ1 channel function

We incorporated the *KCNQ1* variant library into HEK293 cells, LP or LP-KCNQ1-HA, and used dox and selection media to select for successfully integrated cells, as previously described. Variant-only and heterozygous experiments were conducted separately to assess differences in the function of *KCNQ1*-myc variants alone and when co-expressed with an HA-tagged copy of WT *KCNQ1*. Seven days post-transfection, integration efficiency was assessed via flow cytometry. Cells with greater than 70% transfection efficiency were then cultured in HEK media supplemented with triple-drug media: 5 µM ML277, 5 µM R-L3, and 10 nM ouabain for 28 days, with media changes three times per week. Cell pellets were collected at seven-day intervals, including a control sample at day 0 (immediately prior to ML277 and R-L3 treatment). The collection process involved aspirating the HEK media, washing the cells with 1X PBS, resuspending cells in 1 mL of Accutase, and quenching with 1 mL of HEK media. The cell suspension was centrifuged at 300g for 5 minutes, after which the supernatant was removed and the cell pellets were stored at -20**°**C in preparation for Illumina sequencing.

### Post-assay barcode sequencing and score calculation

Genomic DNA was extracted from each abundance or function MAVE sample using a Quick-DNA midiprep kit (Zymo). Barcodes were amplified from at least 2µg of genomic DNA using Q5 polymerase with 25 amplification cycles, using 1 primer that bound the plasmid and 1 primer that bound the landing pad genomic site, to ensure amplification only of successfully integrated plasmids. Each primer pair also had specific barcode sequences on them for de-multiplexing following Illumina sequencing. Bands were excised from a gel and purified with a Qiagen gel extraction kit. Deep sequencing was performed using the Illumina NovaSeq using paired end 150nt reads, targeting at least 25 million reads per sample (Table S1). Barcode counts were quantified in each sample using a custom Python script and normalized by total read count for each sequenced sample (reads per million in that sample).

For the surface abundance assays, scores were calculated following a previously described approach^50^. Read counts for each variant were pooled across their corresponding barcodes in the subassembly. Raw surface abundance scores were then calculated as a weighted average of read frequencies from the samples from the four bins:

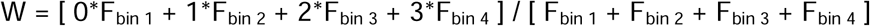

where F_bin_ is the fraction of the variant in the given bin compared to the other 3 bins.

For each replicate, scores were removed for any variant with a mean read frequency across the four bins below 25 per million. Next, for the abundance assay, raw scores were linearly transformed so that the median synonymous variant had a value of 1 and the median nonsense variant (residues 1-610) had a value of 0. For the heterozygous abundance assay, nonsense variants did not have reduced scores, so raw scores were linearly transformed so that the median synonymous variant had a value of 1 and the lowest possible score (100% abundance in bin 1) had a value of 0. Finally, transformed scores were averaged across replicates and renormalized with another linear transformation. For the variant-only abundance assay, scores were first normalized by zone and then merged across zones, because some experimental replicates were performed one zone at a time. For the heterozygous abundance assay, scores were normalized once across the gene. We categorized variants with score estimates within the 95% confidence interval of synonymous variant distribution as normal (higher than the 2.5th percentile or higher than the 97.5th percentile). Variants with scores above the normal range were considered gain-of-abundance. Variants with scores below the lower cutoff but above 0.25 were categorized as partial loss-of-abundance, and variants with scores below 0.25 were categorized as loss-of-abundance.

For the function assays, raw scores for each variant were calculated using a log-transformed ratio of timepoints:

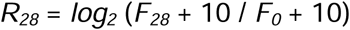

Where *R_28_* is the raw score and *F_x_* is the abundance (reads per million) at Day x.

Next, variants with an initial pre-drug abundance of less than 15 reads per million in a given replicate were removed from that replicate. Raw scores were linearly transformed as above so that the median synonymous variant had a value of 1 and the median nonsense variant (residues 1-610 for the trafficking assays and 1-238 and 308-610 for the function assays) had a value of 0. Noisy variants with a high standard error of the mean across replicates were removed (see Table S2). Finally, transformed scores were averaged across replicates, and renormalized as above so the median nonsense and synonymous variants had scores of 0 and 1. For the two function assays, scores were normalized once across the entire gene, because these experiments were performed by pooling all zones together in a single cell pool. We categorized variants with score estimates within the 95% confidence interval of synonymous variant distribution as normal (higher than the 2.5th percentile or higher than the 97.5th percentile). Variants with scores above the normal range were considered gain-of-function. Variants with scores below the lower cutoff but above 0.25 were categorized as partial loss-of-function, and variants with scores below 0.25 were categorized as loss-of-function.

We next considered that variants involved in binding ML277 and/or R-L3 might have scores in our function assays that are heavily influenced by altered drug binding instead of solely reflecting channel function. Five previous studies have reported *KCNQ1* residues critical for ML277 and/or R-L3 binding and channel activation.^80–84^ From these papers, we curated residues reported to influence drug-responsiveness. We applied two criteria: the residue must be reported in at least two independent studies, and the average effect size score must be a greater than 75% decrease in drug responsiveness compared to WT. Based on these criteria, we determined six key residues, W248, L251, V255, Y267, F335, and F339. In an initial analysis, most missense variants at these six positions also had low function scores, with 96 out of 112 variants (86%) having reduced function scores (below 0.58). We therefore conservatively excluded all missense mutations at these residues from the final MAVE function datasets. Since ML277 and R-L3 were not used in the abundance assays, we did not exclude these residues in the abundance datasets.

### Identification of dominant negative variants

Although the heterozygous function map provides the most direct measurement of dominant negative (DN) variant activity, the moderate level of noise in this assay prompted us to integrate data from all four MAVE datasets to identify DN variants. Variants showing reduced function in all four assays were classified as “severe DN variants”, whereas those with reduced function in three of the four assays were classified as “strong DN variants.” The cutoffs used for the 4 assays were 0.25, 0.25, 0.25, and 0, in the abundance, function, heterozygous abundance, and heterozygous function assays, respectively.

### Data Visualization

Heatmaps were generated in R using the *heatmap.2* function. Violin plots and scatterplots were generated using *ggplot2*. Beeswarm plots were plotted in R using the *ggbeeswarm* package.

### Measurement of KCNQ1 variant surface abundance in non-multiplexed assays

To measure individual MAVE abundance scores in a non-multiplexed format, we individually cloned 26 myc-tagged *KCNQ1* single variant plasmids. “Zone” plasmids containing single mutant zones were generated by Quikchange mutagenesis (Agilent) or synthesized by Genscript. The mutant zones were then subcloned into the promoterless AttB pIRES:mCherry-BlastR plasmid^45,73^ using restriction digestion with appropriate restriction enzymes (Table S9) and T4 ligase (NEB). LP cells were transfected independently, each with 0.6 µg of one of 26 myc-tagged *KCNQ1* variants and 0.1 µg of the BxbI recombinase plasmid (a gift from Pawel Pelczar, Addgene #5127114) using Lipofectamine 3000 (Invitrogen). Each *KCNQ1* variant plasmid was independently transfected and quantified twice to ensure reproducibility. Integrated cells were selected for using Dox-Blast-AP selection. Staining and surface abundance quantification using flow cytometry was performed as described above, on a BD 5-laser Fortessa instrument. Surface abundance scores were calculated by determining the median AF647 fluorescence of mCherry+ cells for each sample, subtracting AF647 background signal from unstained cells, and then normalizing to WT-expressing cells. These non-multiplexed data were combined with curated data from 3 papers^17,30,31^, which were then compared with MAVE scores, for a total of 158 variant comparisons. When the same variant was studied in multiple papers, the surface abundance was averaged across those studies.

### Measurement of KCNQ1 variant channel function by patch clamping

We conducted a PubMed search for “KCNQ1” on 2/11/25 and manually inspected all the resulting paper titles and abstracts. We identified candidate papers that included patch clamp data and curated these studies. Specifically, we curated data on variant-only current density, voltage of activation, and heterozygous current density for 259 variants across 47 papers. The resulting dataset is available in File S2. These studies utilized various heterologous expression systems, including Chinese Hamster Ovary cells (n=221), HEK293 (n=36 measurements), Xenopus oocytes (n=33), mouse cells (n=4), COS7 cells (n=2), HL1 cells (n=1), and guinea pig ventricular myocytes (n=1). When the same variant was studied in multiple papers, the functional properties were averaged across those studies.

In addition to the literature review, we performed patch clamp measurements of 20 *KCNQ1* variants using the SyncroPatch 384 PE automated patch clamp device. The cDNA of WT untagged *KCNQ1* (NM_000218.3) and *KCNE1* (NM_001127669.4) with an IRES cassette (KCNQ1-IRES-KCNE1, synthesised by Genscript Inc., Piscataway, USA) was subcloned into pcDNA5/FRT-hyg-fs.^85^ Twenty *KCNQ1* variants, made using site-directed mutagenesis, were subcloned into this WT template by Genscript. Stable Flp-In HEK293 cell lines co-expressing *KCNQ1* and KCNE1 were generated as described^86^ with minor modifications. Briefly, 60 ng WT or variant KCNQ1-IRES-KCNE1 plasmid was co-transfected with 60 ng pcDNA5/FRT-puro-fs, and 1.8 µg Flp recombinase expressing plasmid pOG44 (Thermo Fisher Scientific, #V600520) using 4.6 µL of Lipofectamine 3000 (Thermo Fisher Scientific, #L3000015) and 4.6 µL of P3000. Plasmids were transfected into Flp-In parental cells (Thermo Fisher, #R78007) seeded as 0.8 × 10L cells per well in 6-well plates, maintained in DMEM (Thermo Fisher Scientific, #11965092) supplemented with 10% FBS and 10 µg/mL of Blasticidin (InvivoGen, #ant-bl-1). Stable cell lines were selected using 100 µg/mL of Hygromycin (Thermo Fisher Scientific #10687010) and 0.25 µg/mL of Puromycin (Thermo Fisher Scientific #A1113803). Cells from T75 flasks were harvested using 1 mL of Accutase (Thermo Fisher Scientific #A1110501) and resuspended in divalent-free external recording solution (in mM: 140 Na, 5 KCl, 1 MgCl2, 10 HEPES, 5 glucose, pH 7.4 with NaOH). Automated patch clamp assays were performed using the Syncropatch 384 PE (Nanion Technologies, Munich, Germany) in the whole-cell configuration using the L-type plates (Nanion Technologies, NPC-384T L). For each 384-well plate, 96 wells contained *KCNQ1* WT positive control, 64 wells each for four *KCNQ1* variants, and 32 wells for non-transfected cells as negative control. External solution consisted of (in mM) 140 Na, 5 KCl, 2 CaCl2, 1 MgCl2, 10 HEPES, 5 glucose, pH 7.4 with NaOH. Internal solution contains (in mM) 60 KF, 50 KCl, 10 NaCl, 10 HEPES, 10 EGTA, and 2 Mg-ATP (pH 7.2 adjusted by KOH). The liquid junction potential (+8 mV) was corrected in all recordings. Voltage-dependent activation was recorded using 2-s voltage steps from -50 mV to + 60 mV in 10 mV increments, with tail currents recorded at -30 mV. Data were analysed and visualised using Nanion DataControl 384 1.9.7.2, Excel 365 (Microsoft), and Prism 10 (GraphPad). Only cells that passed the following QC criteria were included in data analysis: seal resistance > 500 MΩ, capacitance: 2-50 pF, and series resistance: 1-15 MΩ. Tail current density (pA/pF) was calculated from the maximum tail current measured at -30 mV after depolarizing at +60 mV for 2s. Tail current density measurements were square root-transformed to achieve a Gaussian distribution.^87^ Data for variants were then normalised to the mean of the square-root transformed WT data from the same plate, and mean baseline current from control cells not expressing *KCNQ1* was subtracted. The steady-state activation data analysis included only cells with peak currents greater than 300 pA at + 60 mV. The voltage at half activation (V1/2) was obtained by fitting the Boltzmann function to the tail current recorded at -30 mV using Prism 10 with the bottom value constrained between 0 and 0.05. Only V1/2 activation values that satisfied the following criteria were included: goodness of curve fit (R^2^ > 0.99), slope values (between 5 and 25), and the product of current amplitude and series resistance (< 20 mV).

For the example trace showing *I*_Ks_ current by manual patch clamping, Chinese Hamster Ovary cells were transiently cotransfected with WT *KCNQ1* and *KCNE1*-expressing plasmids and patch clamped, as previously described.^50^

### Principal Component Analysis and merged assay scores

Scores from the four *KCNQ1* MAVE datasets were filtered to retain only variants with scores from all four datasets (“complete” variants). Principal Component Analysis (PCA) and *k*-means clustering were performed on complete variants using the StandardScaler, PCA, and KMeans modules from scikit-learn v1.4.0. Clustering was tested for two to seven clusters; based on visual inspection of projections on the first two principal components, a six-cluster solution was selected as it best captured known variant classes (dominant negative, haploinsufficient, gain-of-function, partial loss-of-function, and two normal classes).

Classifiers were trained on complete missense variants with established clinical classifications (P/LP or B/LB; “labeled” variants; see Variant classification methods). Models were implemented using LogisticRegression, RandomForestClassifier, and GaussianNB from scikit-learn v1.4.0, with performance evaluated via Leave-One-Out Cross-Validation (LeaveOneOut module). Following model validation, a final model trained on all labeled missense variants was used to assign positive-class probabilities to unlabeled (i.e., unclassified) complete missense variants.

### Immunostaining and Confocal imaging

Immunostaining and confocal imaging were performed as described,^88^ except for different antibodies (anti-myc Alexa Fluor 647 and anti-myc Alexa Fluor 488; Cell Signaling).

### SpliceAI predictions of splice disruption

Our cDNA-based experiments do not assess the impact of variants on splicing. To model the potential splicing effects of *KCNQ1* variants, we obtained pre-calculated SpliceAI scores for all possible exonic variants of the *KCNQ1* genomic region from the SpliceAI dataset hosted on Illumina BaseSpace. Aggregate SpliceAI predictions were calculated using the formula P(aberrant splicing) = 1 × (1 − AG) × (1 − AL) × (1 − DG) × (1 − DL), where AG represents the probability of acceptor gain, AL the probability of acceptor loss, DG the probability of donor gain, and DL the probability of donor loss.^69,89^ These predictions were converted from hg19 to hg38 coordinates using the R implementation of the *liftover* tool in the *rtracklayer* package. We filtered the predictions to include only single nucleotide variants (SNVs) that impact coding sequences, and for each possible coding variant, we calculated the maximum aggregate SpliceAI score.

### ACMG assay calibration

To calibrate the MAVE assays we first generated sets of benign/likely benign and pathogenic/likely pathogenic control variants. We obtained variant classifications for all *KCNQ1* protein-coding variants in ClinVar (accessed on 9/18/25). Variants were considered in terms of their protein consequence. Variants with multiple unique ClinVar DNA changes resulting in identical effects on the protein sequence were combined, keeping the more definitive classification (e.g. P/LP and VUS was assigned a P/LP classification). 218 missense variants had a pathogenic/likely pathogenic ClinVar classification; these were considered the primary control “P/LP” set. Only 13 missense variants had a benign/likely benign ClinVar classification. To supplement this set, we implemented *KCNQ1*-specific Variant Curation Expert Panel (VCEP) variant frequency v.1.0.0 cutoffs (available at https://cspec.genome.network/cspec/ui/svi/doc/GN112). The VCEP guidelines recommend the use of BA1 (stand-alone allele frequency criterion) at a cutoff of MAF>0.004 in gnomAD and BS1 (strong allele frequency criterion) for variants at a cutoff of MAF>0.0004 in gnomAD. From gnomAD v.4.1.0, we obtained the maximum filtering allele frequency across five ancestry groups (African/African-American, East Asian, European non-Finnish, Latino/Admixed-American, or South Asian). We identified 5 missense variants that met BA1 (MAF>0.004) and 10 missense variants that met BS1 (0.004>MAF>0.0004). Lastly, we included a third set of variants as benign controls: variants with 0.0004>MAF>0.000134 and no reported LQT cases in a recent curation of 2,440 LQT patients with *KCNQ1* variants from the literature.^90^ The cutoff of 0.000134 was chosen because it is double the allele frequency of the most common pathogenic/likely pathogenic missense *KCNQ1* variant in ClinVar (K362R/c.1085A>G; gnomAD maximum filtering allele frequency of 6.7×10^-5^). In total, 29 missense B/LB variants were obtained through this approach, of which MAVE scores were available for 28 variants. We note that using relatively low allele frequency cutoffs for B/LB *KCNQ1* variants is supported by the Clingen-approved computational framework for allele frequency cutoffs^91^ and an empirical study of the allele frequency distribution of variants found in Long QT syndrome cases.^4^ We refer to the set of Clinvar variants, supplemented by these semi-rare variants as additional benign controls, as “Clinvar+". In addition to the primary analysis using the Clinvar+ missense variants for assay calibration, we also performed additional assay calibrations. Overall we used 4 benign/pathogenic “truth sets": 1) Clinvar+ missense variants (primary analysis), 2) Clinvar+ all variants (missense, nonsense, and synonymous), 3) Clinvar missense variants, and 4) Clinvar all variants.

We used three methods to calibrate the assays. First, receiver operating characteristic (ROC) curves were calculated and visualized using the *proc* package in R. Second, we used the binary “OddsPath” approach set forth by the ClinGen Sequence Variant Interpretation working group,^26^ as extended by Fayer et al.^92^ Third, we applied a continuous kernel density log-likelihood ratio (LLR) approach.^93^ Outlier values falling outside the range of (-0.3, 1.5) were excluded. Using the *maveLLR* R package, we utilized the *buildLLR.kernel* and *drawDensityLLR* functions with a bandwidth parameter set to 0.2. We adjusted *drawDensityLLR* to enable the inclusion of a benign-moderate category with symmetrical evidence strengths to the pathogenic-moderate category. The primary ROC and OddsPath analyses were conducted using the ClinVar+ missense truthset, while the continuous LLR analysis was performed on the ClinVar+ all-variant truthset to ensure a denser set of control variants.

### JLNS literature curation

Variants previously reported in association with Jervell and Lange-Nielsen syndrome (JLNS) were identified through a PubMed search using the search term “Jervell KCNQ1” on May 1, 2025 (File S3). These papers were manually curated, identifying 53 papers describing 52 unique variants found in at least one individual with JLNS (homozygous or compound heterozygous). Of these 52 variants, 42 had an available MAVE function score, which were plotted as a violin plot.

### SpliceAI

Precomputed SpliceAI scores for all coding variants on chromosome 11 (hg38) were obtained from Illumina Basespace.^43^ Using *tabix*^94^, the scores were filtered to isolate variants within the genomic region of *KCNQ1* (chr11:2,441,176-2,851,765). The filtered data was then processed using R to identify the corresponding cDNA and protein changes for each genomic coordinate variant.

### Visualization of protein structures

Secondary structure schematics were plotted using a custom R script, using boundaries obtained from the structure PDB ID: 6UZZ.^14^ The HD helix was manually added to complete the schematic, as it was not present in this structure. MAVE scores were mapped onto cryoEM structures of K_V_7.1 in PyMOL. PDB ID 6V01^14^ was used for the PIP_2_-bound structure, and PDB ID 6UZZ^14^ was used for all other K_V_7.1 structures with MAVE score maps. PDB ID 6UZZ was also used in Figure 1C to illustrate the tetrameric and monomeric structures of K_V_7.1. The HD helix is not present in cryo-EM structures, so two crystal structures of the HD helix were used (PBD ID 3BJ4 or 3HFE).^36,52^ The three-dimensional structure of *KCNQ1* nonsense variant S253X was predicted using ColabFold v1.5.5,^95^ which incorporates AlphaFold2^96^ and MMseqs2^97^ for protein structure prediction. The pipeline, executed with default settings, generated sequence alignments using MMseqs2 and templates from the PDB100 database. Visualization was performed using PyMOL.

### Case-control risk ratio calculations for missense KCNQ1 variants

Following a previously published approach,^98^ we calculated the risk ratio (RR) associated with *KCNQ1* missense variants using published LQTS case counts from an International LQTS Genetics Consortium (n=1,847 LQTS cases)^4^ and putative controls from gnomAD v4.1.^99^ Risk ratios (RRs) were calculated by the formula (a/[a+c])/(b/[b+d]), where a = disease cases with a variant, b = controls/reference population with a variant, c = disease cases without a variant, d = controls/reference population without a variant.^100^ In addition to calculating RRs for *KCNQ1* MAVE data, we compared the performance of the MAVE data to computational predictions for single-amino acid substitutions. AlphaMissense predictions^101^ for *KCNQ1* missense variants (ENST00000155840.11) were downloaded from https://alphamissense.hegelab.org/. CPT-1 pre-computed variant effect predictions were downloaded from https://doi.org/10.5281/zenodo.8140323.102 VARITY scores were retrieved from http://varity.varianteffect.org/ by searching for “KCNQ1” and downloading the associated files.^103^ REVEL scores were retrieved by downloading results from https://sites.google.com/site/revelgenomics/ and identifying lines associated with “ENST00000155840."^104^

### Biobank studies—overview

We integrated three datasets for the biobank analyses: All of Us, BioVU, and UK Biobank. Based on data availability, BioVU and UK Biobank data were used for the QTc analysis and BioVU and All of Us data were used for the LQTS code analysis.

### BioVU association studies

This study used data from BioVU, Vanderbilt University Medical Center’s (VUMC) biorepository linking DNA samples to de-identified electronic health records.^105^ Phenotypic data available from the deidentified electronic health records include diagnostic and procedure codes as well as electrocardiographic data. We extracted *KCNQ1* genotypes from a cohort of 249,927 participants in BioVU with available whole-genome sequencing. Sequencing, variant calling, and quality control was performed as previously described.^106^ The genotypes were extracted in Genome Reference Consortium Human Build 38 (GRCh38) coordinates through Hail Matrix files available from BioVU’s Terra cloud analysis environment. Principal components and ancestry data were obtained using predefined datasets created by the AGD flagship working group. Long QT syndrome (LQTS) was defined as presence of ICD-9 code 426.82 and/or ICD-10 code I45.81 in the health record. Electrocardiogram data including automated QTc interval measurements were extracted when available. Electrocardiograms with missing QTc values, QTc values outside the physiologic range (150-1000 ms), QRS duration <20 ms or >1000 ms, or PR interval <40 ms or >1000 ms were excluded from analysis. ECGs from participants with pacemakers were also excluded. Prolonged QTc was defined using sex-specific thresholds: any QTc >450ms in men and >470ms in women. We evaluated associations between MAVE function scores and three outcomes: QTc interval, prolonged QTc prevalence, and LQTS diagnosis. The function scores were modeled both as continuous variables and as function categories (LOF, partial LOF, normal, GOF) in separate models. Ordinary least squares (OLS) regression was used to model non-linear relationships between function scores or categories and QTc interval. We then added robust variance estimation clustering to account for repeated measurements within participants. Generalized linear models with logistic regression were used to predict prolonged QTc and LQTS diagnosis by function scores or categories. All models adjusted for age, sex, and the first five principal components of genetic ancestry. For continuous predictors, we used restricted cubic splines with 3 degrees of freedom, with knot placement at default locations determined by the distribution of the predictor. Statistical analyses were performed in RStudio (version 4.4.2) on the Terra cloud platform.

### All of Us association analyses

This study was performed in alignment with the ethical principles outlined in the All of Us Policy on the Ethical Conduct of Research. In the All of Us Research Program, informed consent for all participants is conducted in person or through an eConsent platform that includes primary consent, Health Insurance Portability and Accountability Act authorization for research EHRs and consent for return of genomic results. The protocol was reviewed by the Institutional Review Board (IRB) of the All of Us Research Program. The All of Us Institutional Review Board follows the regulations and guidance of the National Institutes of Health Office for Human Research Protections for all studies. Analysis of the All of Us cohort was conducted with whole-genome sequencing data from the All of Us Controlled Tier Dataset v8 (414,349 participants).

Post-sequencing variants and sample QC were performed by the All of Us Data and Research Center. Data were extracted and processed as previously described.^107^ Briefly, coding variants were extracted from Hail MatrixTables (version 0.2.130, Hail Team) and filtered for Phred quality score > 20, individual missingness < 10%, minimum read coverage depth of 7, and presence in at least one participant passed the allele balance threshold of 0.20. Carriers of variants with minor allele frequency <0.1% were included in this analysis. Where participants had >1 scored *KCNQ1* variant, only the lowest-scoring variant was retained. Long QT Syndrome diagnosis was ascertained based on ICD-9 and ICD-10 codes in electronic health records available in All of Us. For each participant, a minimum of two LQTS codes was required for an LQTS diagnosis; participants with only one LQTS code were excluded from the analysis. To assess the association between function scores and LQTS penetrance, the proportion of subjects with an LQTS code was assessed according to bins of functional categories (LOF, partial LOF, normal, GOF). These bins each contained at least 20 participants, meeting the All of Us requirement prohibiting the publication of data representing fewer than 20 participants.

### UK Biobank association analyses

We assessed the association between *KCNQ1* MAVE scores and corrected QT interval (QTc) in the UK Biobank. We accessed exome sequencing data from 49,960 individuals and identified rare *KCNQ1* coding variants with allele frequency <1×10LL. For each carrier, we computed a function score by multiplying the *in silico* function impact score of the specific mutation they carried by the dosage of the effect allele (0, 1, or 2). Among this cohort, 13,924 individuals had at least one available QTc measurement, including 137 mutation carriers. For individuals with multiple QTc measurements, we selected the highest QTc value to use in downstream analysis. To assess the association between mutation score and QTc, we fit a linear regression model of QTc on binary mutation carrier status and an interaction term between carrier status and mutation score to estimate the effect of mutation score specifically among carriers. The model was adjusted for age, sex, the first five genetic principal components, and QT-prolonging drug exposure (amiodarone, amitriptyline, amitriptyline hydrochloride + perphenazine 10mg/2mg tablet, azithromycin, chloroquine, ciprofloxacin, citalopram, clarithromycin, erythromycin, erythromycin product, erythromycin + zinc acetate 40mg/12mg/ml topical solution, escitalopram, haloperidol, hydroxychloroquine, levofloxacin, ondansetron, procainamide, quetiapine, risperidone, sotalol, sotalol hydrochloride + hydrochlorothiazide 80mg/12.5mg tablet, topical erythromycin).

## RESOURCE AVAILABILITY

### Lead contact

Andrew Glazer.

## Materials availability

All unique/stable reagents generated in this study are available from the corresponding author with a completed materials transfer agreement.

## Data and code availability

The MAVE datasets are available in the supplemental files and will be deposited in MaveDB and ClinVar upon paper publication. Analysis code is available at https://github.com/GlazerLab/KCNQ1_MAVE. Any additional information required to reanalyze the data reported in this paper is available from the corresponding author upon request.

## Data Availability

All data produced in the present study are available in the manuscript or upon reasonable request to the authors.

## Acknowledgements

We thank Ashli Chew, Zerubabell Daniel, and Lynn Hall for assistance with experiments, Kayla Carmichael and Jeff Calhoun for assistance with computational analyses, Doug Fowler and Kenneth Matreyek for providing the HEK293 landing pad cells, and Al George, Chuck Sanders, Henrike Heyne, Francesca Rissom, and the CardioVar consortium for helpful discussions. This project was supported by NIH grants R01 HL164675, R00 HG010904, R35 GM150465, R01 HL160863, K12 CA090625, K08 HG014001; American Heart Association fellowships 25PRE1360565, 23CDA1042141; and a Medical Research Future Fund: Genomics Health Futures Mission grant MRF2016760.

## Author contributions

Conceptualization: MLH, DMR, and AMG. Formal analysis: MLH, EO, MCL, JES, QS, TA, RC, AM, DJB, RK, JWO, JIV, CAN, DMR, and AMG. Funding acquisition: BMK, CAM, FPR, VNP, ERAA, DMR, and AMG. Investigation: MLH, QS, RED, TY, MRF, MEC, DJB, JLA, MAB, RK, ES, CAN, and AMG. Supervision: GD, BK, CAM, BCK, FPR, VNP, EAA, JIV, CAN, DMR, and AMG. Writing—original draft: MLH, EO, MCL, DMR, and AMG. Writing—review and editing: all authors.

## Declaration of Interests

CAM: Atman Health (founder and shareholder), Tanaist (founder and shareholder), TMA Precision Health (scientific advisor and shareholder), Everyone Medicines (scientific advisor), LifeMD (non-executive director and shareholder). VNP: consultant or scientific advisory board member for Lexeo Therapeutics, Solid Biosciences, and Constantiam Biosciences. EA: Founder: Personalis, Deepcell, Svexa, Saturnus Bio, Swift Bio; Founder Advisor: Candela, Parameter Health; Advisor: Pacific Biosciences; Non-executive director: AstraZeneca, Dexcom; Publicly traded stock: Personalis, Pacific Biosciences, AstraZeneca; Collaborative support in kind: Illumina, Pacific Biosciences, Oxford Nanopore, Cache, Cellsonics. FPR: advisor and shareholder with Constantiam Biosciences, shareholder in SeqWell. No other authors declare any interests.

## Declaration of generative AI in the writing process

During the preparation of this work the author(s) used ChatGPT, Gemini, and Grammarly for computer programming assistance or to improve the readability and language of the manuscript. After using these tools, the authors reviewed and edited the content as needed and take full responsibility for the content of the published article.

## Notes

### Author Declarations

The protocol was reviewed by the Institutional Review Board (IRB) of the All of Us Research Program. The Institutional Review Boards of the All of Us Research Program, UK Biobank, and Vanderbilt University Medical Center gave ethical approval for this work.

