## Supplemental Figures and Tables for "Mapping the Functional Landscape of *KCNQ1* to Define Ion Channel Mechanisms and Arrhythmia Risk"

Figure S1: Quantification of selection efficiency of function assay.

Figure S2: Mutation coverage by zone.

Figure S3: Examination of outlier synonymous variants.

Figure S4: Charge sensitivity

Figure S5: Coiled-coil structure and mutational sensitivity

Figure S6: Windowed truncated functionality of *KCNQ1* nonsense variants in some contexts.

Figure S7: Representative traces of individual *KCNQ1* variants.

Figure S8: Activation curves of individual *KCNQ1* variants.

Figure S9: Structural mapping of scores and their relation to pore proximity

Figure S10: Pilot measurement of dominant negative effects.

Figure S11: Experimental details of heterozygous MAVE experiments

Figure S12: Detailed analysis of principal component analysis and clustering datasets

Figure S13: Performance of classifiers trained on labeled missense variants using all four assays.

Figure S14: MAVE scores by clinical classification and variant type across four truth sets

Figure S15: Depletion of *KCNQ1* variants with abnormal scores from the population.

Figure S16: Comparison of MAVE scores with computational predictors

Figure S17: Additional risk ratios from case-control comparison.

Figure S18: Representative histogram of subassembly cutoff at lowest frequency score

**Supplemental Tables:**

Table S1: Sequencing depth and barcode quality metrics

Table S2: Variant coverage, composition, and replicate reproducibility

Table S3: Surface abundance levels for individual *KCNQ1* variants

Table S4: Calibration of MAVE assays with benign and pathogenic variants

Table S5: Variant counts in *k*-means clustering

Table S6: Examination of 6 VUS from LQTS patients

Table S7: Variants with elevated SpliceAI scores

Table S8: Case-control risk ratios

Table S9: KCNQ1 Plasmid Zone System

Table S10: Primers used in this study (non-Illumina)

Table S11: Illumina sequencing primers used in this study

Table S12: Summary of MAVE sequencing samples and primer pairs

**Supplemental Files:**

File S1: Comprehensive summary dataset

File S2: Patch clamp literature curation

File S3: JLNS literature curation

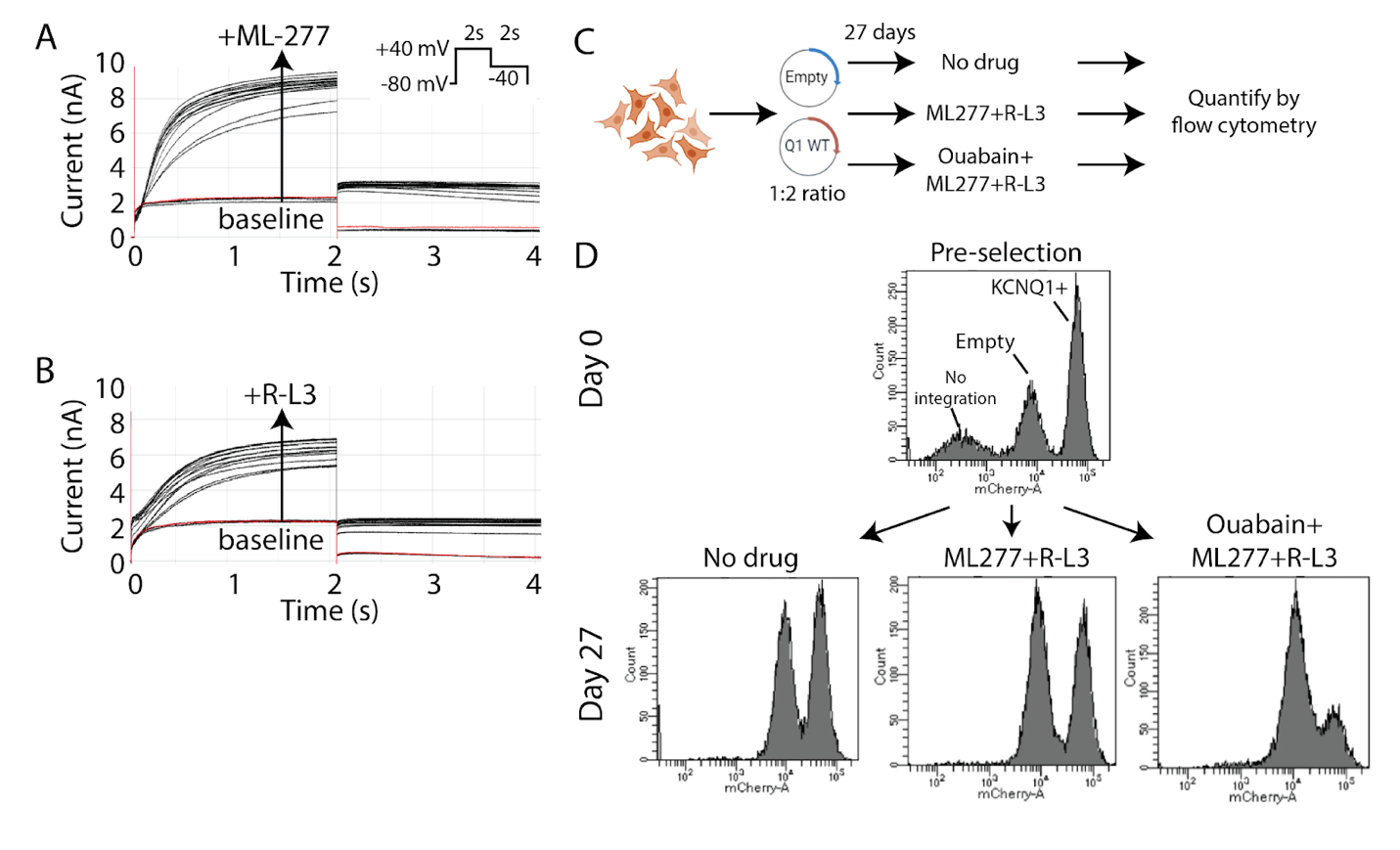

**Figure S1: Quantification of selection efficiency of function assay.** A) Representative electrophysiological traces from repetitive stimulation assays performed on HEK293 cells stably expressing *KCNQ1*, following acute exposure to ML-277 or B) R-L3. C) Schematic representation of the experimental design for developing the function assay. D) Representative histogram depicting the relative abundance of HEK293 cells expressing *KCNQ1* versus an empty vector. Cells were transfected with a 1:2 ratio of empty vector to *KCNQ1* WT. The analysis compared the cell populations before drug treatment (day 0) and after 27 days of treatment under different conditions: no drug, ML277 + R-L3, and ML277 + R-L3 + Ouabain. On Day 0, the number of cells expressing *KCNQ1* is higher than those expressing the empty vector. On Day 27, cells treated with either no drug or the combination of ML277 and R-L3 exhibit similar abundance levels. In contrast, cells treated with ML277, R-L3, and ouabain show a substantial reduction in the proportion of *KCNQ1*-expressing cells compared to the initial empty vector condition. These results are quantified and normalized in Figure 1D. The data consist of two biological replicates for each condition.

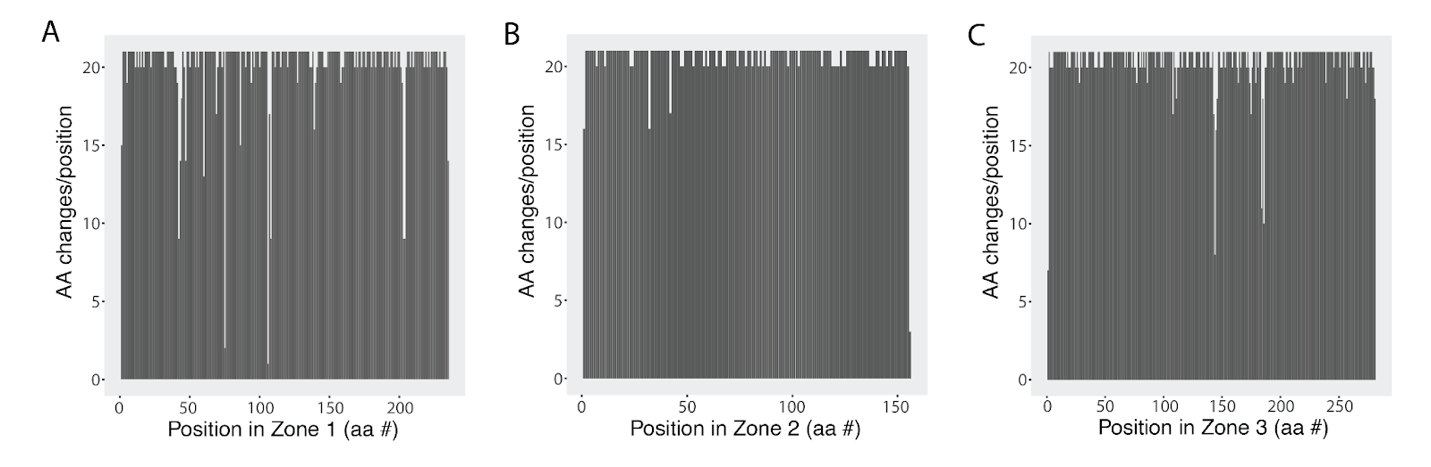

**Figure S2: Mutation coverage by zone.**

These bar graphs show the saturation of amino acid changes per position across all three zones of *KCNQ1*. Each bar corresponds to a single amino acid position, indicating the extent of mutation coverage across the specified region.

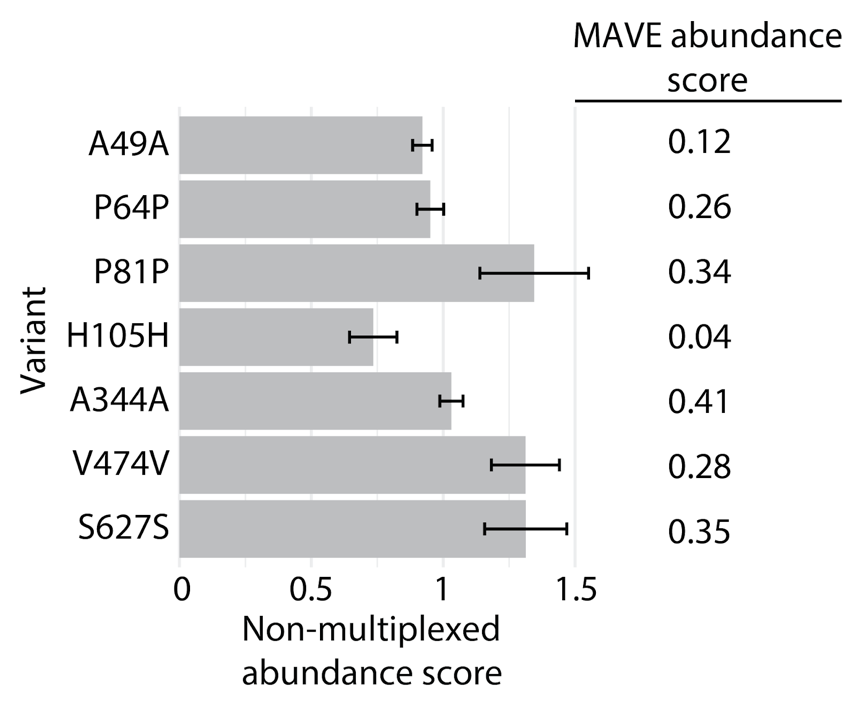

**Figure S3: Examination of outlier synonymous variants.** Among 541 synonymous variants with available MAVE scores, 21 (3%) had MAVE surface abundance scores below 0.5. Single variant validation measurements of 7 of the 21 variants were performed. All 7 variants had near-normal abundance (0.7-1.3), suggesting that most rare outlier synonymous MAVE scores reflect rare experimental noise in the MAVE assay. The data consist of two biological replicates for each variant.

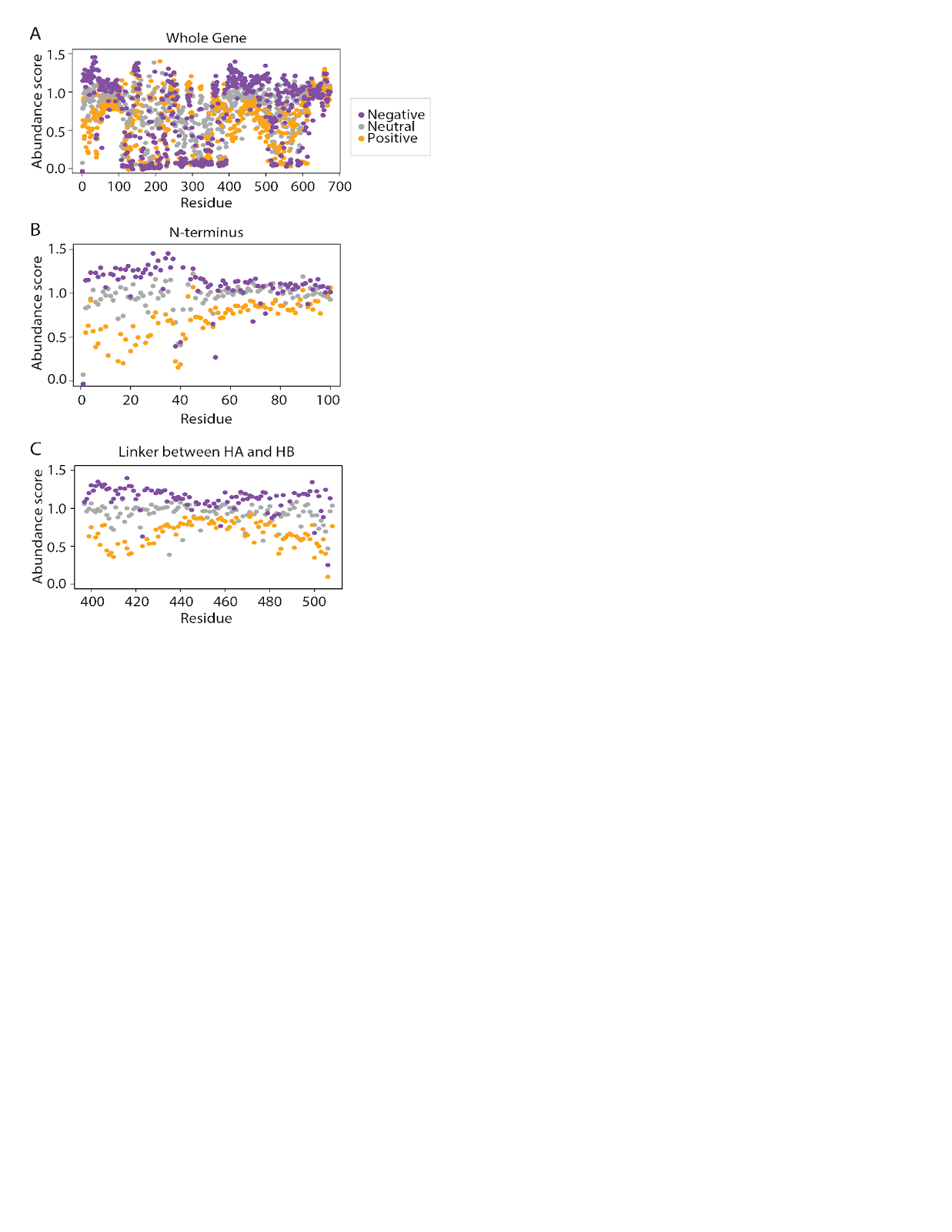

**Figure S4: Regional charge sensitivity.** Average abundance scores of variants categorized by their effect on residue charge: more negative, neutral, or more positive. A) whole protein; B-C) the first 40 aa of the N-terminus (B) and the linker between the HA and HB helices (C) show differential charge sensitivity.

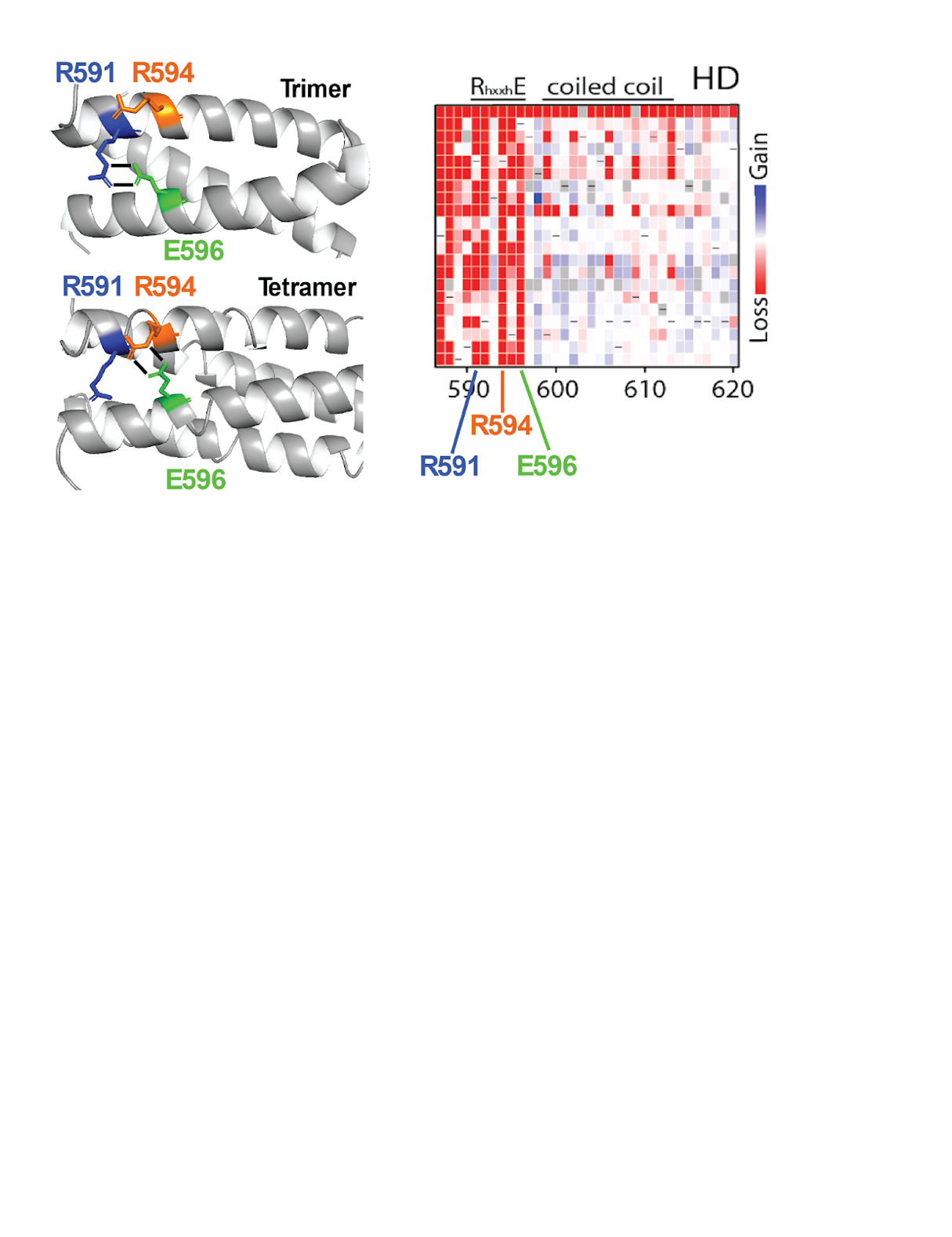

**Figure S5: Coiled-coil structure and mutational sensitivity.** Two structural models show key interactions that support oligomerization of the HD helix: the R591–E596 salt bridge bonds promote trimer formation, whereas the R594–E596 salt bridge bonds stabilize the tetrameric coiled-coil. The heatmap shows that residues R591, R594, and E596 are highly sensitive to mutation, with most substitutions leading to loss of function. Eight variants at R591, R594, and E596 are currently classified as pathogenic or likely pathogenic (P/LP) in ClinVar.

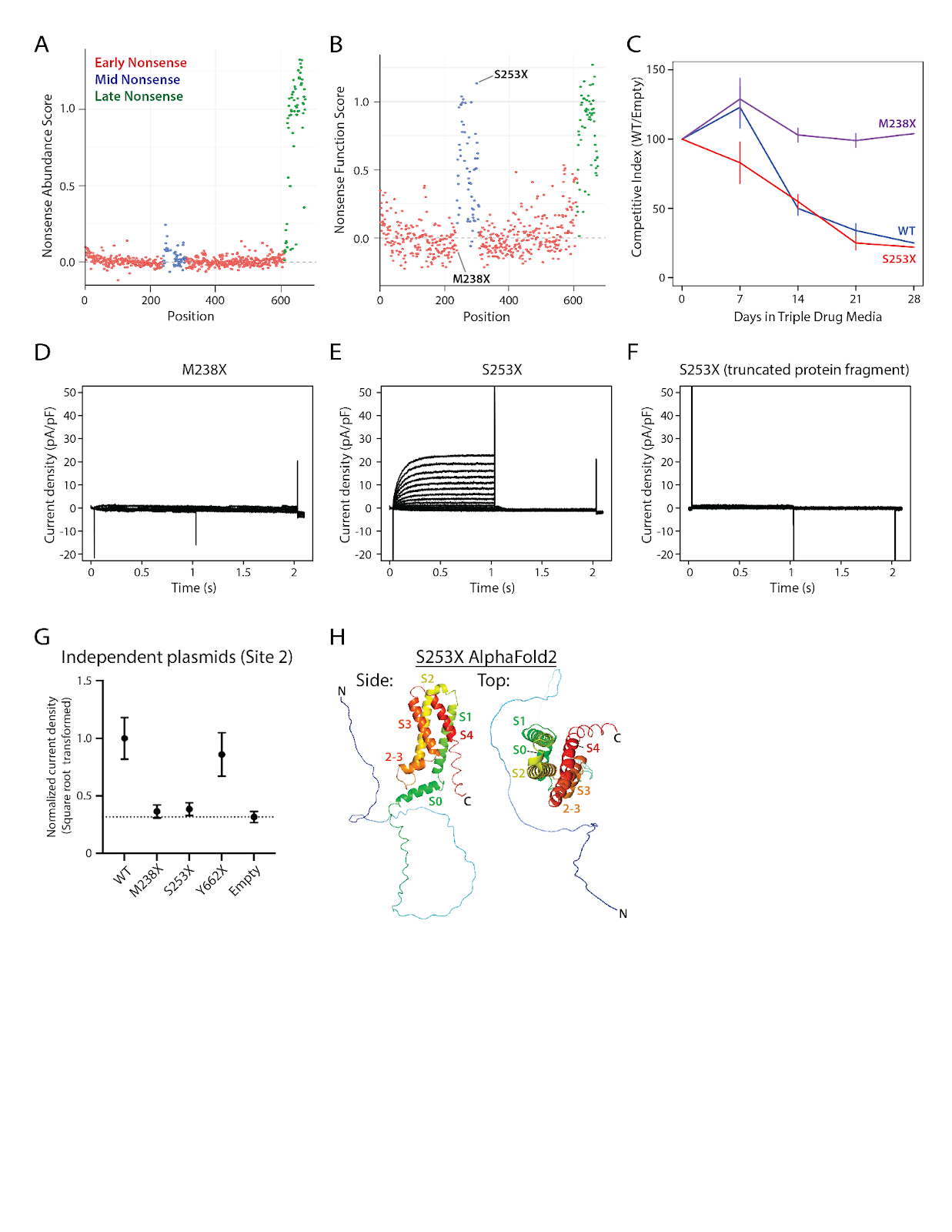

**Figure S6: Windowed Truncated Functionality of *KCNQ1* nonsense variants in some contexts.**

A-B) Dot plots of MAVE scores show (A) abundance and (B) function of *KCNQ1* nonsense variants along the protein. Early nonsense variants (residues 1-610) exhibit reduced abundance, while late nonsense variants (>610) show normal or increased levels. Function scores are predominantly reduced for early nonsense variants, except for residues 239-307, while late nonsense variants generally retain or enhance function. C) Competitive growth assays in triple-drug media over 28 days reveal that the early nonsense variant M238X confers resistance (loss-of-function), while WT and the mid-nonsense variant S253X remain sensitive, aligning with MAVE scores. D-F) Patch clamp measurements of HEK293 cells expressing M238X, S253X, or S253X (truncated protein fragment containing only residues 1-252). The S253X nonsense variant surprisingly had K_V_7.1-like current, but the truncated fragment did not. G) Quantification of current density of cells expressing *KCNQ1* variants at Site 2 (different site and plasmid system than panels D-F. At this site, S253X did not have K_V_7.1-like current over background. H) AlphaFold2-predicted structure of K_V_7.1 S253X. Left: side view, Right: top view. The voltage-sensing domain (VSD) remains intact, while the pore and C-terminal cytoplasmic domains are absent. This truncation is predicted to prevent normal potassium conductance. Overall, these experiments are consistent with a model of cryptic readthrough of the stop codon in some plasmid systems, producing K_V_7.1-like current.

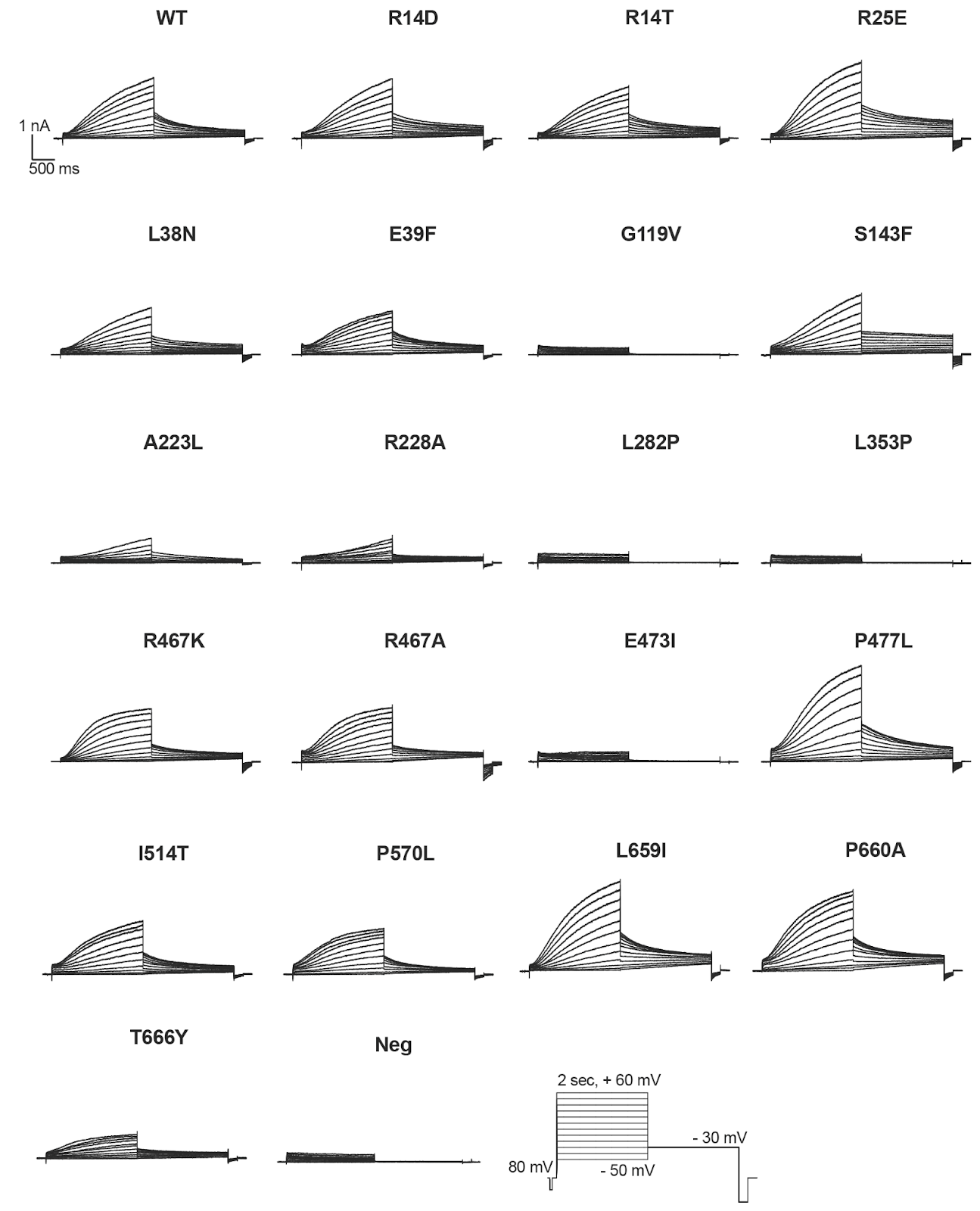

**Figure S7: Representative traces of individual *KCNQ1* variants.**

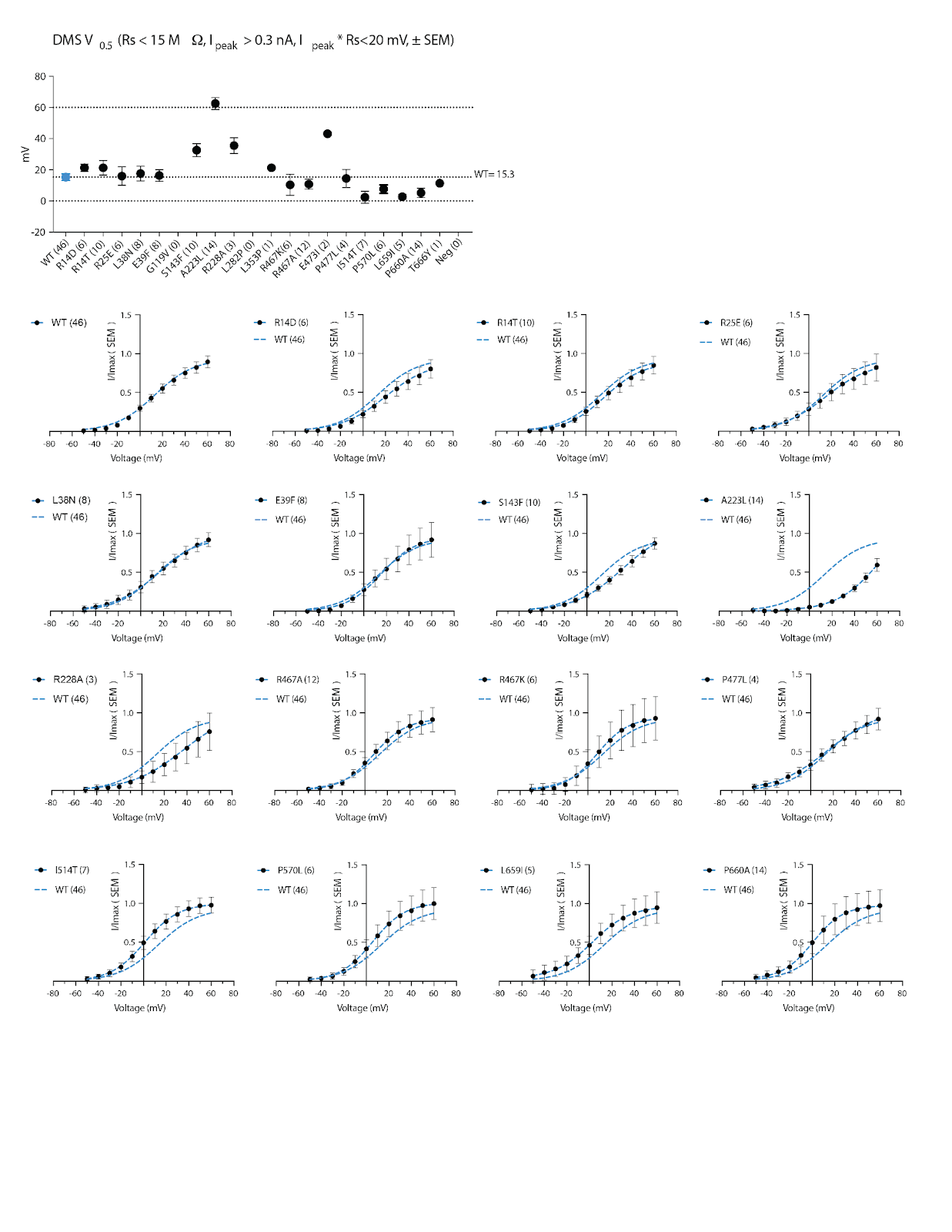

**Figure S8: Activation curves of individual *KCNQ1* variants.**

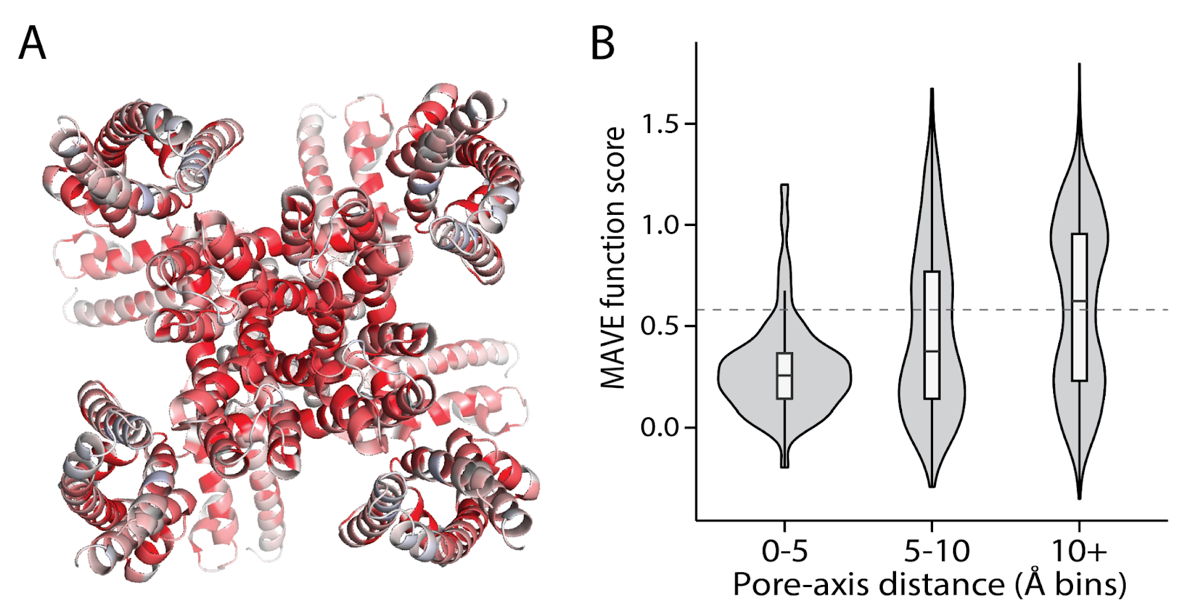

**Figure S9: Structural mapping of function scores and their relation to pore proximity.**

Left: Heatmap showing function scores on the tetrameric protein structure, with blue indicating high function and red indicating loss of function, concentrated near the pore. Right: Violin plot showing MAVE function scores relative to distance from the pore. Regions closer to the pore consistently exhibit lower function scores, underscoring the trend of function loss associated with proximity to the pore.

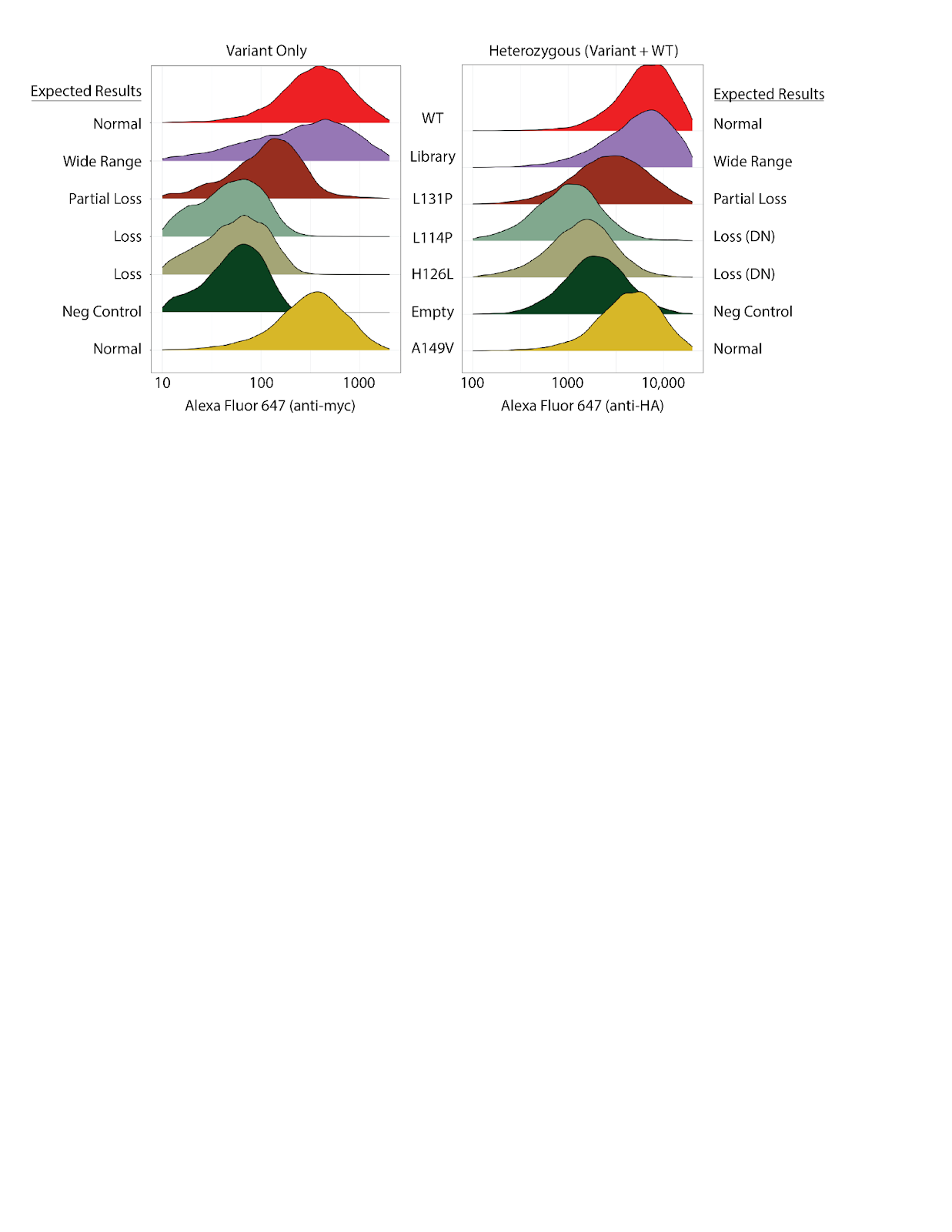

**Figure S10: Pilot measurement of dominant negative effects.** To validate the cell system for measuring dominant negative effects of *KCNQ1* variants, LP or LP-*KCNQ1*-HA cells (which stably express WT *KCNQ1*-HA) were integrated with myc-tagged *KCNQ1* variants, including controls and known loss-of-abundance variants. Surface expression was tracked using an anti-myc antibody (for variant only experiments) or an anti-HA antibody (for heterozygous experiments). We measured 4 previously studied mutants with a range of dominant negative properties.^17^ Flow cytometry measurements confirmed that L114P and H126L reduced WT *KCNQ1* surface abundance, consistent with their expected dominant negative behavior. As anticipated, A149V showed normal trafficking without dominant negative effects, while L131P exhibited a slight impact on WT *KCNQ1*, contrary to predictions of neutrality. These results support the use of this system in MAVE experiments to assess all *KCNQ1* variants’ impacts in a heterozygous context.

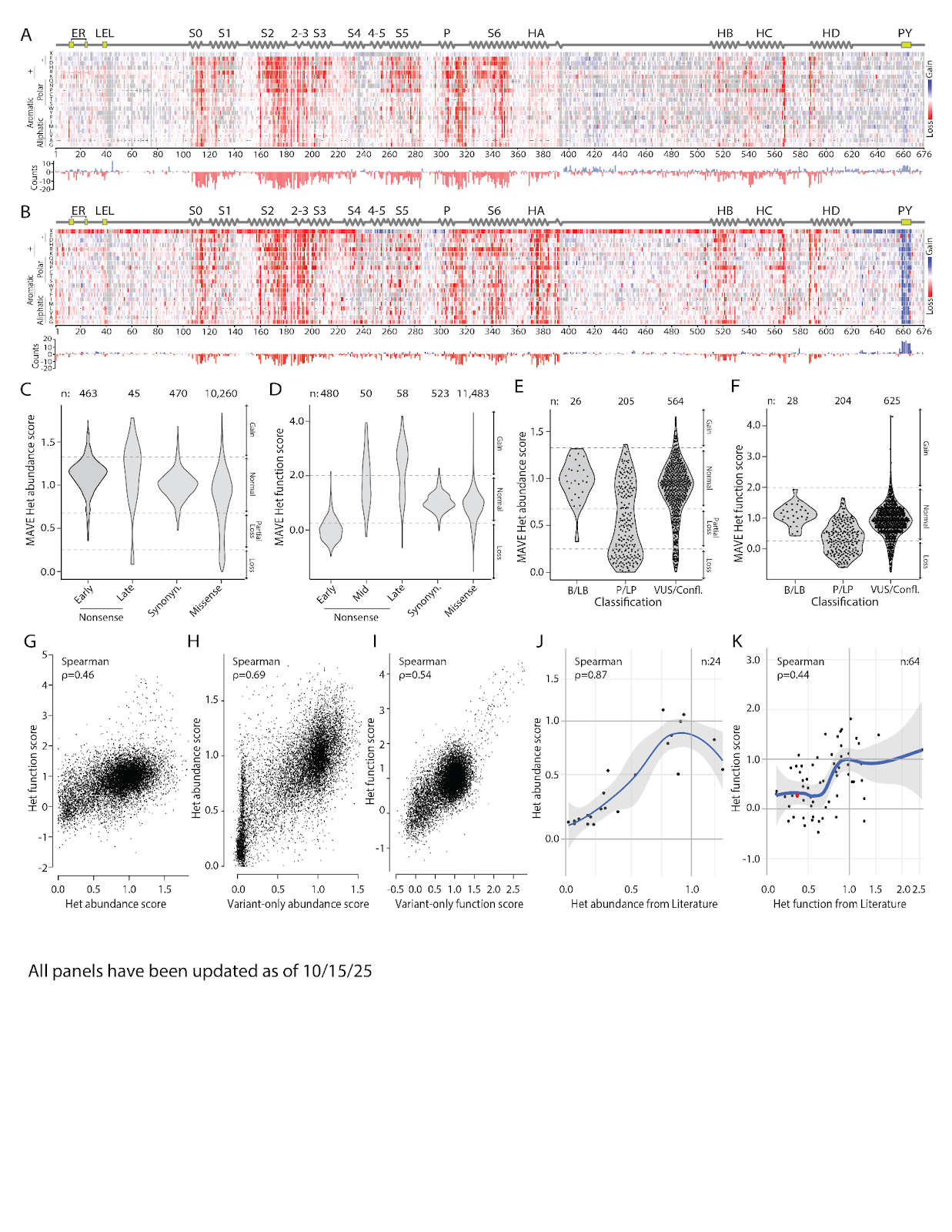

**Figure S11: Experimental details of heterozygous MAVE experiments.** LP cells expressing WT *KCNQ1*-HA were transfected with a barcoded library of *KCNQ1*-myc variants. Cell surface abundance and function assays were performed as described previously. A) Heatmap showing the cell surface abundance of WT *KCNQ1*-HA when co-expressed with *KCNQ1*-myc variants, to measure dominant negative effects on WT *KCNQ1*-HA, and the relative abundance of variants showing increased (blue) or decreased (red) surface abundance at specific amino acid positions. B) Heatmap illustrating survival of cells co-expressing WT *KCNQ1*-HA and variant *KCNQ1*-myc following 28 days of triple-drug treatment, and the relative abundance of variants showing increased (blue) or decreased (red) function at specific amino acid positions. For A and B, the color gradient indicates surface abundance or function, ranging from red (low) to white (normal) to blue (high). Black dashes indicate WT residues at each position. C) Distribution of heterozygous surface abundance scores for early nonsense (residues 1–610), late nonsense (>610), synonymous, and missense variants. D) Distribution of function scores for early nonsense (residues 1–238, 308-610), mid nonsense (239-307), late nonsense (>610), synonymous, and missense variants. E–F) Violin plots comparing heterozygous (het) MAVE scores for E) abundance and F) function in variants classified as Benign/Likely Benign (B/LB), Pathogenic/Likely Pathogenic (P/LP), or Variants of Uncertain Significance (VUS). G–I) Dot plot comparisons of heterozygous function and abundance scores: G) het function versus het abundance (ρ = 0.46), H) het abundance versus variant-only (var-only) abundance (ρ = 0.69), and I) het function versus hom function of missense variants (ρ = 0.54). J–K) Comparisons of MAVE abundance scores with literature-reported data: J) het abundance scores (ρ = 0.87) and K) het peak current scores (ρ = 0.44). Red dot highlights G314S, a known *KCNQ1* variant with dominant negative effects on WT *KCNQ1* function.

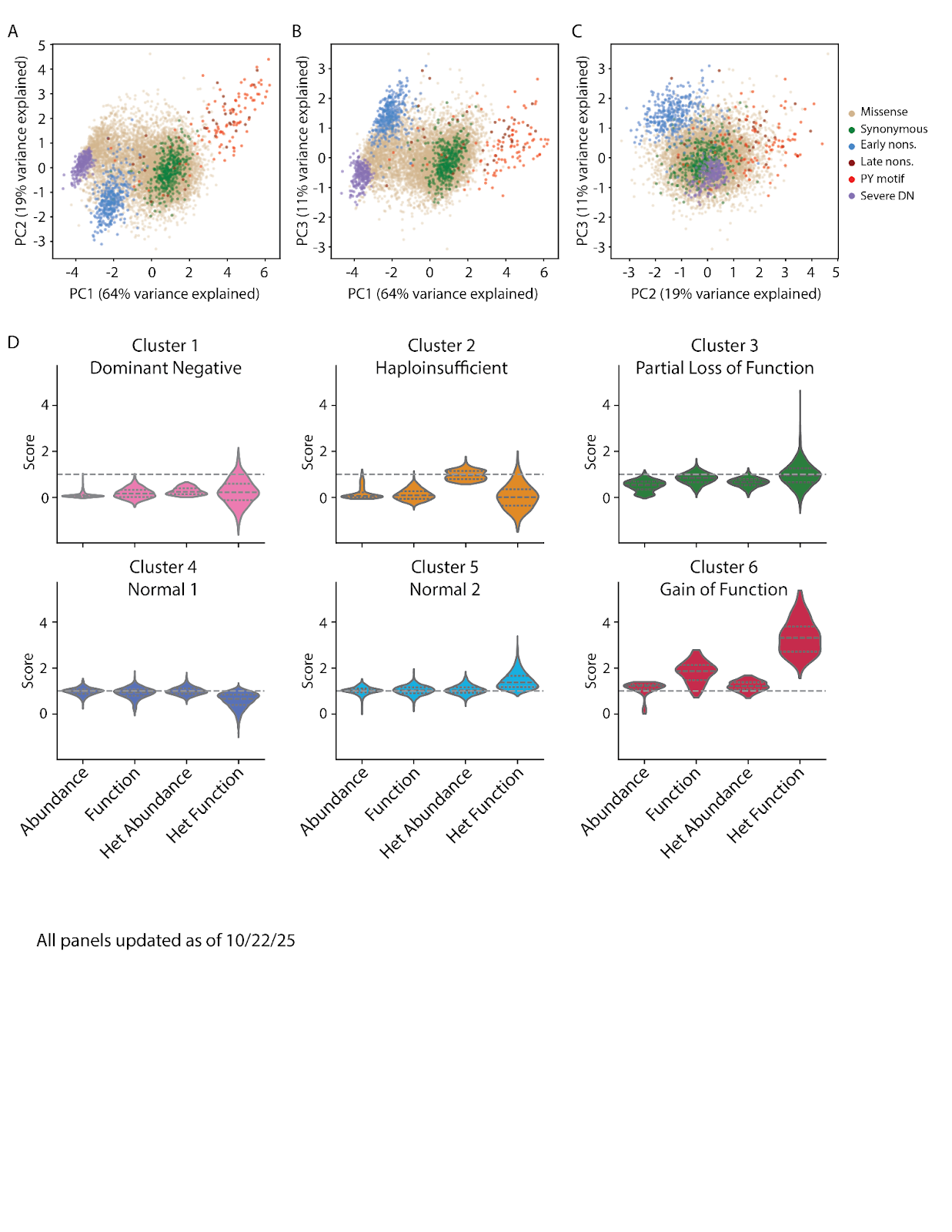

**Figure S12: Detailed analysis of principal component analysis and clustering datasets.**

A-C) Principal Component Analysis of the 4 MAVE scores. The plots show PC1 vs PC2, PC1 vs PC3, and PC2 vs PC3. Legend: color coding by variant class. D) *k*-means clustering (K=6) of the 4 MAVE scores. The plots show the distribution of raw scores for each cluster. Plots of these variants color-coded by cluster are shown in Figure 5C.

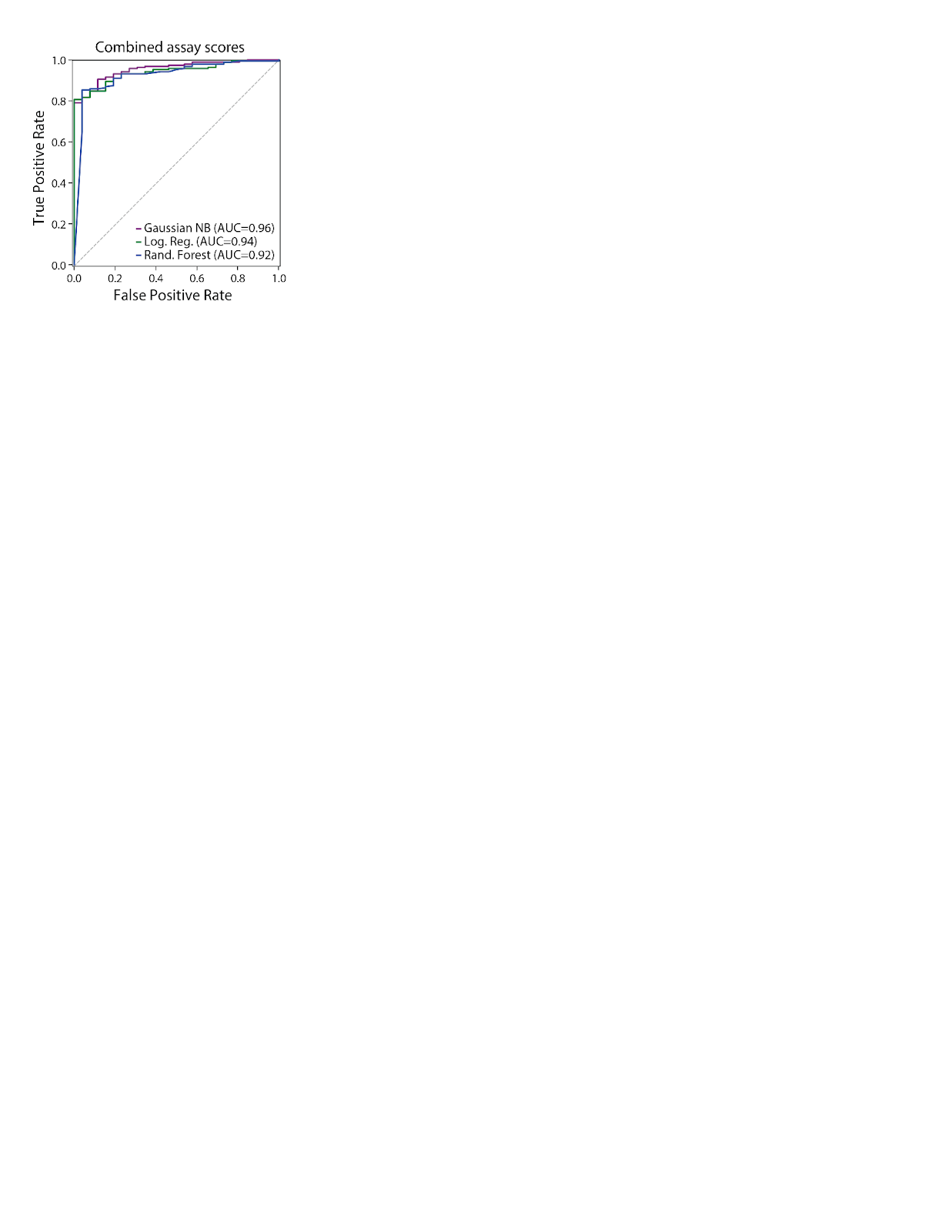

**Figure S13: Performance of classifiers trained on labeled missense variants using all four assays.** ROC curves showing performance of 3 methods that combine all 4 assays together (Gaussian Naive Bayes [NB], Logistic regression, and Random forest).

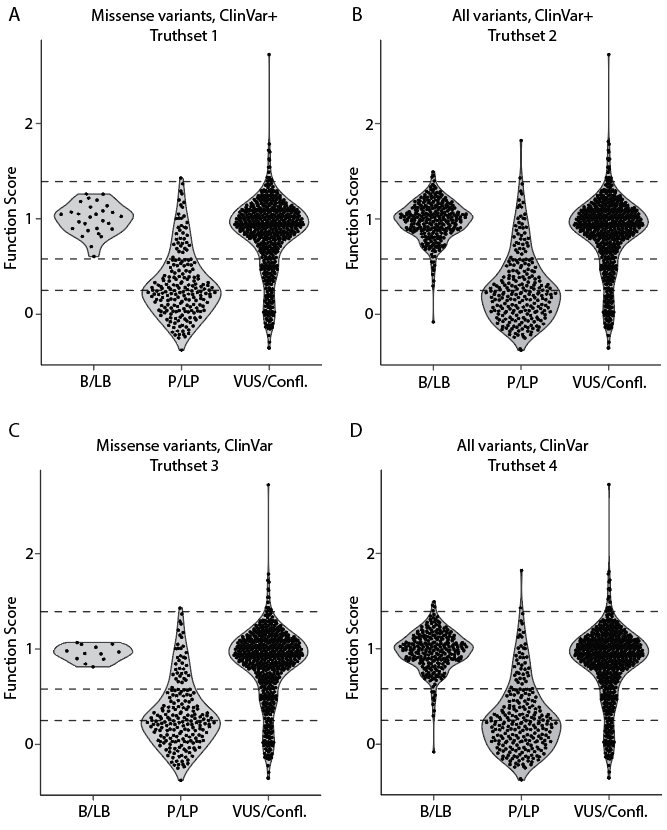

**Figure S14: MAVE function scores by clinical classification and variant type across four truth sets.** MAVE function scores are grouped by variant classification (B/LB, P/LP, VUS/Conflicting). A) Truth Set 1: ClinVar+, missense variants only. This panel reproduces Panel 3J and represents the primary analysis of the study. B) Truth Set 2: ClinVar+, including missense, nonsense, and synonymous variants. C) Truth Set 3: ClinVar only, missense variants only. D) Truth Set 4: ClinVar only, including missense, nonsense, and synonymous variants. ClinVar+ includes ClinVar-annotated pathogenic/likely pathogenic (P/LP) and benign/likely benign (B/LB) variants, supplemented with high allele frequency variants as additional benign controls (see Methods for details).

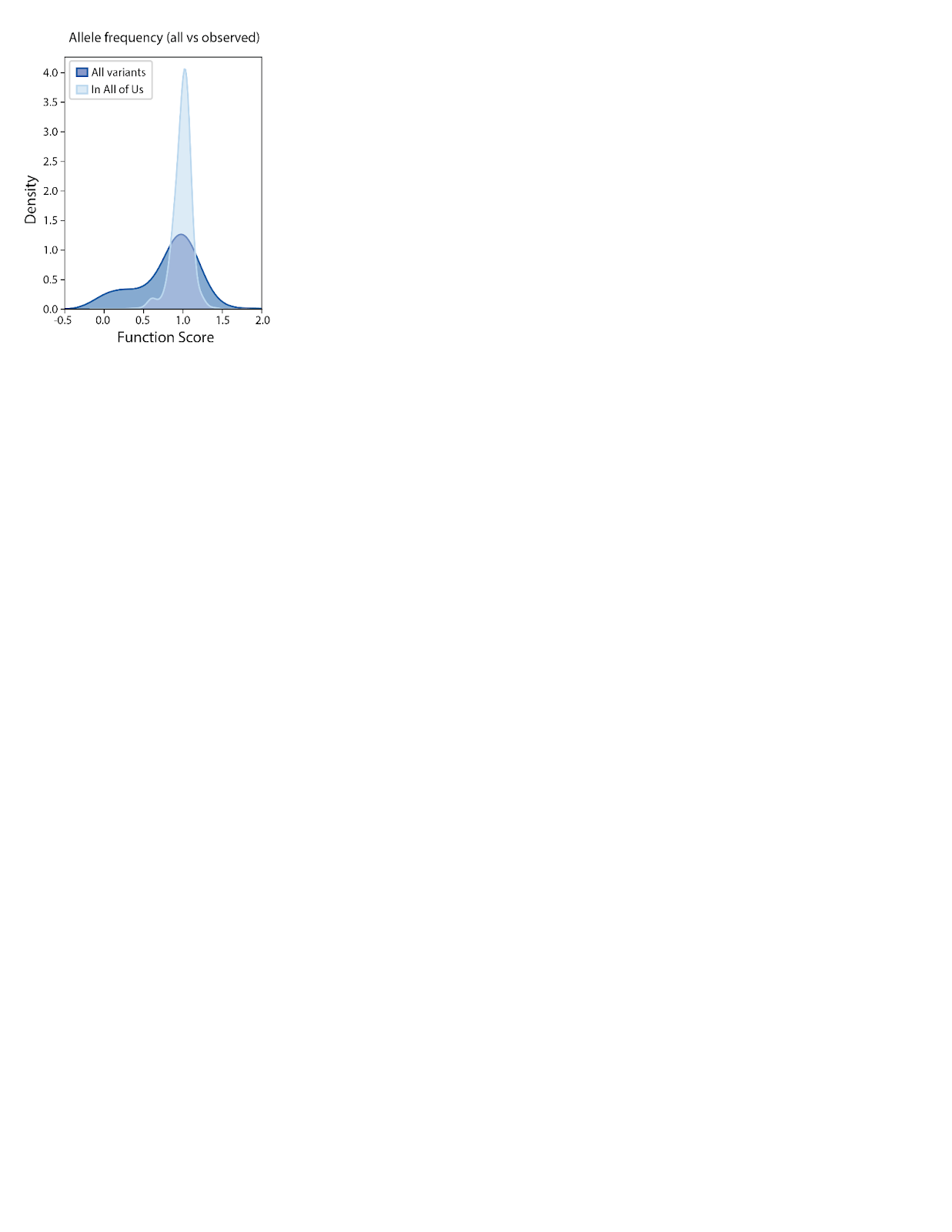

**Figure S15: Depletion of *KCNQ1* variants with abnormal scores from the population.** Distribution of scores for all assayed variants (dark blue) versus those found in the All of Us cohort (light blue).

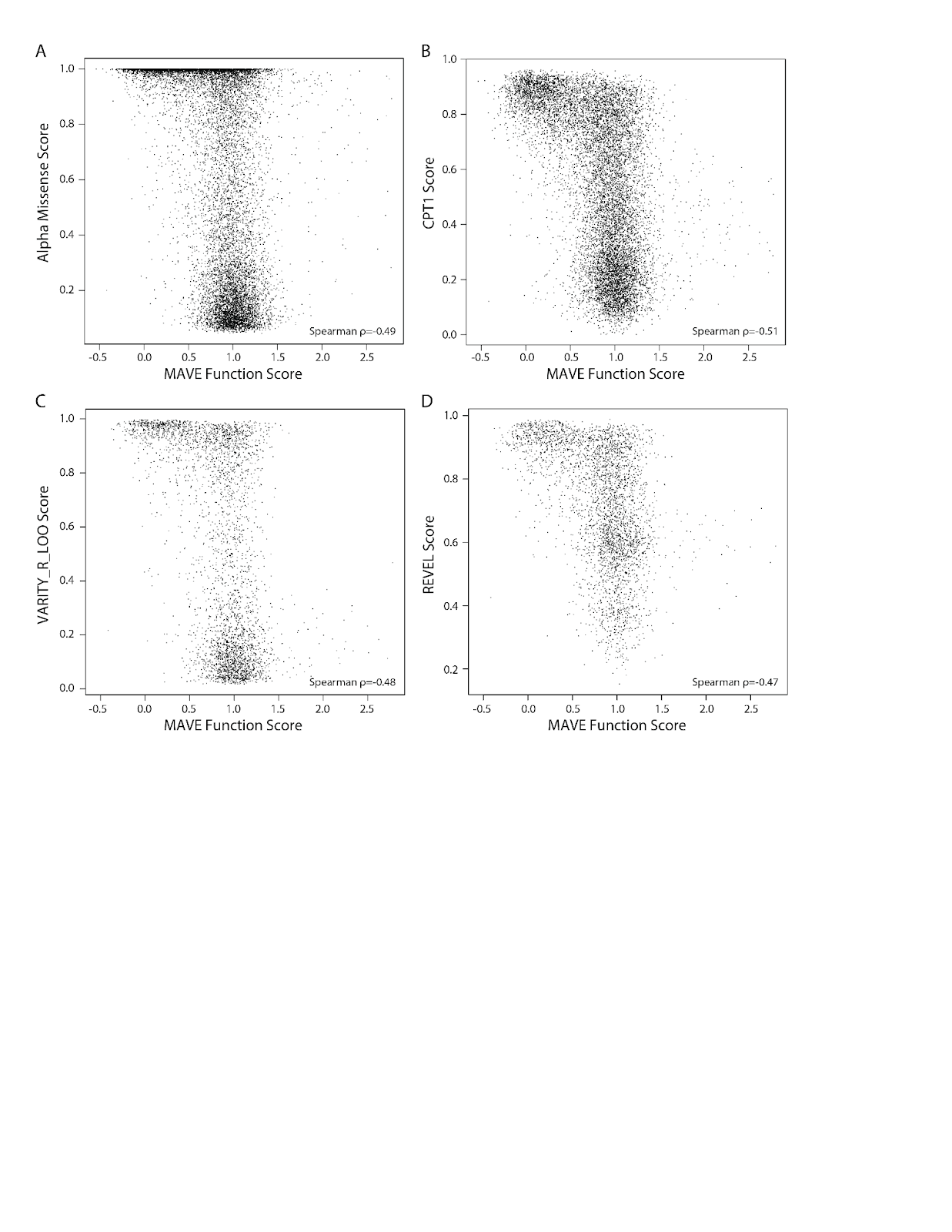

**Figure S16: Comparison of MAVE scores with computational predictors.** Scatter plots show the relationship between MAVE function scores and four computational predictors: (A) alpha missense score, (B) CPT1 score, (C) VARITY score, and (D) REVEL score. Each point represents a variant, with its MAVE score on the x-axis and the predictor score on the y-axis.

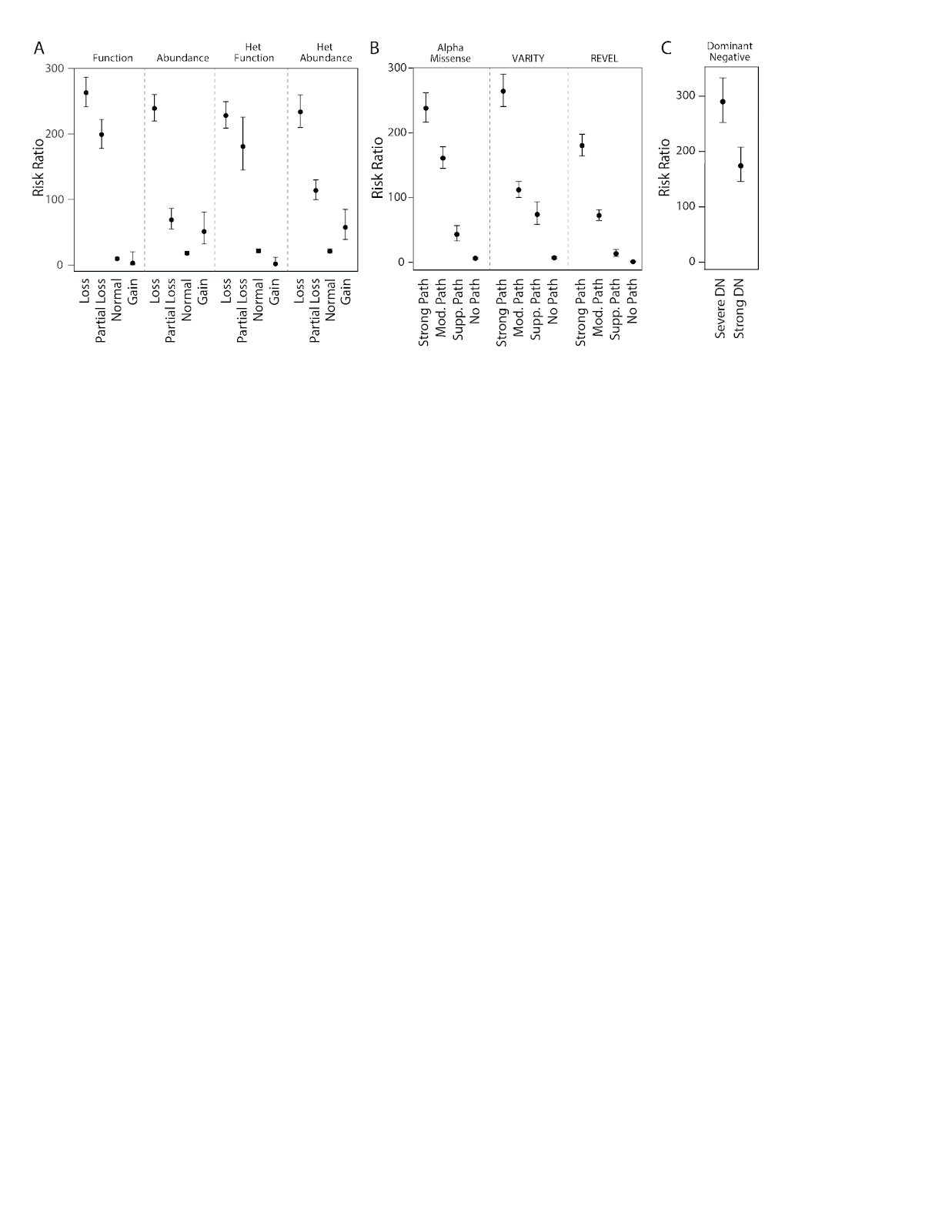

**Figure S17: Additional risk ratios from case-control comparison.** Risk ratios (±95% CI) for variant classes in an LQTS cohort compared to gnomAD controls are presented. Numbers above data points indicate unique variant counts. Risk ratios are derived from A) MAVE assays, B) computational predictors, and C) severe/strong dominant negative variants.

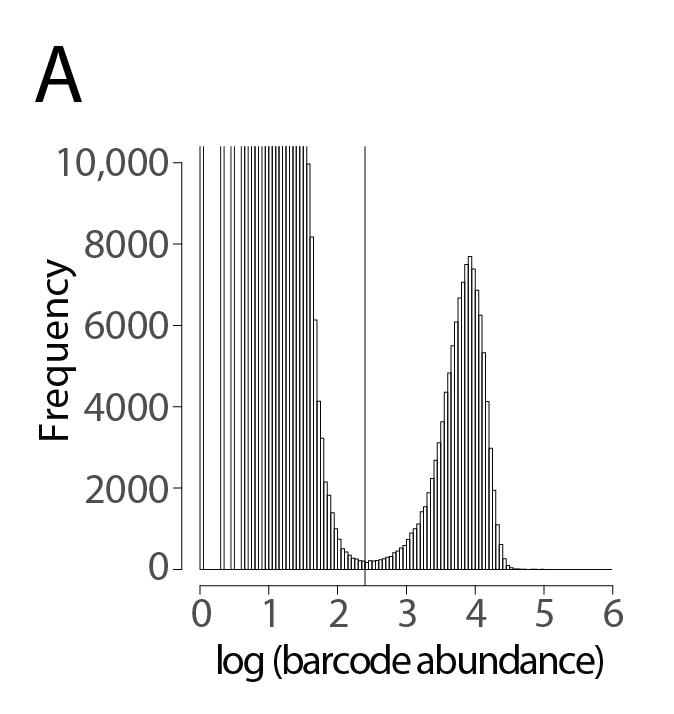

**Figure S18: Representative histogram of subassembly cutoff at lowest frequency score.**

Barcode frequencies were plotted on a log scale and a cutoff was chosen at the low point of the trough between the two modes.

**Table S1: Sequencing depth and barcode quality metrics**

| **Experiment** | **# Replicates** | **Library IDs** | **Total Reads** | **Mean reads/sample** | **Total reads w/good barcodes** | **Mean reads w/good barcodes** |
| --- | --- | --- | --- | --- | --- | --- |
| Abundance | 5 | 8952, 9004, 9418, 9612, 10029 | 1594.8M | 66.5M | 838.7M | 41.9M |
| Function | 6 | 11823, 12246 | 1049.1M | 56.7M | 1055.0M | 35.2M |
| Het Abundance | 3 | 10211 | 455.6M | 38.0M | 314.1M | 26.2M |
| Het Function | 15 | 12331, 12332, 13772, 13773 | 1859.3M | 40.4M | 1388.6M | 30.2M |

**Table S2: Variant coverage, composition, and replicate reproducibility**

| **Experiment** | **Cutoffs** | **Total Variants** | **Early/mid/late nonsense** | **Synonymous**  **Variants** | **Missense**  **Variants** | **Replicate correlations** | **% of possible variants** |
| --- | --- | --- | --- | --- | --- | --- | --- |
| Abundance | ≥ 100/rep,  ≥ 2 reps,  SEM < 0.3 | 13,252 | 510/49/63 | 541 | 12,089 | 0.90  (0.89-0.92) | 93% |
| Function | 15/replicate,  ≥ 2 reps,  SEM < 0.3 | 12,112 | 441/50/58 | 494 | 11,069 | 0.50  (0.46-0.54) | 85% |
| Heterozygous Abundance | ≥ 100/rep  ≥ 2 reps,  SEM < 0.3 | 11,248 | 428/35/45 | 470 | 10,270 | 0.56  (0.56-0.57) | 79% |
| Heterozygous  Function | 15/rep,  ≥ 2 reps,  SEM < 0.5 | 12,693 | 480/50/58 | 523 | 11,582 | 0.20  (0.13-0.29) | 89% |
| All assays | N/A | 13,392 | 510/61/64 | 544 | 12,213 | N/A | 94% |

**Table S3: Surface abundance levels for individual *KCNQ1* variants.** Each value represents the mean surface expression, determined from 2 experimental replicates in validation flow cytometry experiments.

| **Variant** | **MAVE Abundance Score** | **Individual Abundance Score (mean)** | **Individual Abundance Score (SE)** | **Category** | **Variant Type** |
| --- | --- | --- | --- | --- | --- |
| R14D | 1.44 | 133.8 | 26.4 | ER retention | missense |
| R14T | 1.33 | 141.1 | 37.1 | ER retention | missense |
| R25E | 1.4 | 225.1 | 24.5 | ER retention | missense |
| L38N | 0.15 | 29.3 | 9.5 | LEL motif | missense |
| E39F | NA | -0.8 | 0.8 | LEL motif | missense |
| G119V | 0.12 | 16.4 | 2.8 | VUS | missense |
| S143F | 0.38 | 55.8 | 7.2 | VUS | missense |
| A223L | 1.51 | 108.8 | 26.4 | S4 helix | missense |
| R228A | 1.52 | 142 | 46 | S4 helix | missense |
| Q260K | NA | 54.6 | 4.7 | Partial LOF | missense |
| L282P | 0.1 | -2.5 | 0.5 | VUS | missense |
| A352T | 0.57 | 32.6 | 0.3 | Partial LOF | missense |
| L353P | 0.07 | -0.5 | 1 | VUS | missense |
| S409R | 0.41 | 40.6 | 1.3 | Partial LOF | missense |
| K467A | 1.24 | 127.2 | 23.9 | Cytosolic | missense |
| K467R | 1.19 | 257.4 | 3.8 | Cytosolic | missense |
| E473I | 0.09 | 1.2 | 0.6 | Cytosolic | missense |
| P477L | 0.32 | 24.1 | 7.2 | Cytosolic | missense |
| E487K | 0.32 | 49.5 | 0.1 | Benign, low MAVE abundance | missense |
| L506V | 0.82 | 68.8 | 1.6 | Partial LOF | missense |
| I514T | 0.24 | 19.5 | 5.3 | VUS | missense |
| N551Y | 0.64 | 54.2 | 0.5 | Partial LOF | missense |
| P570L | 0.32 | 39.7 | 8.9 | VUS | missense |
| L659I | NA | 205.2 | 63.6 | PY motif | missense |
| Q664W | NA | 296.6 | 64.8 | PY motif | missense |
| T666Y | 0.03 | -0.3 | 0.1 | PY motif | missense |
| A49A | 0.16 | 92.1 | 3.7 | Discordant synonymous | synonymous |
| P64P | 0.19 | 95.1 | 5.1 | Discordant synonymous | synonymous |
| P81P | 0.38 | 134.5 | 20.6 | Discordant synonymous | synonymous |
| H105H | 0.09 | 73.5 | 9 | Discordant synonymous | synonymous |
| A344A | 0.41 | 103.1 | 4.4 | Discordant synonymous | synonymous |
| V474V | 0.32 | 131.2 | 12.9 | Discordant synonymous | synonymous |
| S627S | 0.39 | 131.3 | 15.6 | Discordant synonymous | synonymous |

**Table S4: Calibration of MAVE assays with benign and pathogenic variants**

Odds of pathogenicity (OddsPath) calculations for MAVE assays, using the approach of Brnich et al.[31892348] Four truth sets were used: 1 = ClinVar+, missense only (primary analysis); 2 = ClinVar+, missense+nonsense+synonymous; 3 = ClinVar only, missense only; 4 = ClinVar only, missense+nonsense+synonymous. ClinVar+ indicates ClinVar-annotated P/LP and B/LB variants, supplemented with high allele frequency variants in the benign group (see methods). The bolded row in line 1 is the primary calibration featured in this study (function assay scores calibrated with Truth Set 1). This primary calibration is also displayed in Figure 5F.

| **MAVE**  **assay** | **Truth**  **Set** | **Benign +**  **Normal**  **function** | **Benign + Abnormal function** | **Pathogenic + Normal function** | **Pathogenic + Abnormal function** | **OddsPath**  **(Path)** | **Evidence strength (PS3)** | **OddsPath (Benign)** | **Evidence strength (BS3)** |
| --- | --- | --- | --- | --- | --- | --- | --- | --- | --- |
| Function | 1 | **28** | **0** | **36** | **168** | **23.06** | **strong** | **0.1765** | **moderate** |
|  | 2 | 285 | 13 | 41 | 208 | 19.15 | strong | 0.1722 | moderate |
|  | 3 | 12 | 0 | 36 | 168 | 9.88 | moderate | 0.1765 | moderate |
|  | 4 | 264 | 13 | 41 | 208 | 17.8 | moderate | 0.1728 | moderate |
| Abundance | 1 | 26 | 2 | 59 | 158 | 10.19 | moderate | 0.2928 | supporting |
|  | 2 | 302 | 16 | 60 | 209 | 15.44 | moderate | 0.2349 | supporting |
|  | 3 | 11 | 1 | 59 | 158 | 8.74 | moderate | 0.2966 | supporting |
|  | 4 | 282 | 15 | 60 | 209 | 15.38 | moderate | 0.2349 | supporting |
| Het Function | 1 | 27 | 1 | 97 | 107 | 14.69 | moderate | 0.4931 | indeterminate |
|  | 2 | 290 | 19 | 111 | 143 | 9.16 | moderate | 0.4656 | supporting |
|  | 3 | 12 | 0 | 97 | 107 | 6.29 | moderate | 0.4755 | supporting |
|  | 4 | 270 | 18 | 111 | 143 | 9.01 | moderate | 0.4661 | supporting |
| Heterozygous Abundance | 1 | 25 | 1 | 76 | 129 | 16.36 | moderate | 0.3856 | supporting |
|  | 2 | 266 | 14 | 116 | 133 | 10.68 | moderate | 0.4904 | indeterminate |
|  | 3 | 11 | 0 | 76 | 129 | 6.92 | moderate | 0.3707 | supporting |
|  | 4 | 248 | 13 | 116 | 133 | 10.72 | moderate | 0.4903 | indeterminate |

**Table S5: Variant counts in K-means clusters**

| **Category** | **Cluster 1**  **DN** | **Cluster 2**  **Haploinsufficient** | **Cluster 3**  **Partial LOF** | **Cluster 4**  **Normal 1** | **Cluster 5**  **Normal 2** | **Cluster 6**  **Gain of function** |
| --- | --- | --- | --- | --- | --- | --- |
| Missense | 1402 | 460 | 1412 | 3291 | 2761 | 97 |
| Synonymous | 1 | 5 | 5 | 255 | 172 | 1 |
| Early Nonsense | 8 | 367 | 7 | 0 | 0 | 0 |
| Late Nonsense | 0 | 3 | 4 | 1 | 6 | 26 |
| PY motif | 2 | 1 | 1 | 2 | 5 | 88 |
| Severe DN | 290 | 0 | 0 | 0 | 0 | 0 |
| Strong DN | 426 | 294 | 1 | 0 | 0 | 0 |
| **P/LP 1 (main)** | **109** | **26** | **14** | **35** | **7** | **0** |
| **B/LB 1 (main)** | **0** | **0** | **2** | **11** | **13** | **0** |
| P/LP 2 | 110 | 61 | 15 | 36 | 7 | 0 |
| B/LB 2 | 0 | 2 | 6 | 146 | 112 | 0 |
| P/LP 3 | 109 | 26 | 14 | 35 | 7 | 0 |
| B/LB 3 | 0 | 0 | 1 | 4 | 6 | 0 |
| P/LP 4 | 110 | 61 | 15 | 36 | 7 | 0 |
| B/LB 4 | 0 | 2 | 5 | 137 | 103 | 0 |

**Table S6: Examination of 6 VUS from LQTS patients**

| **Variant** | **LQT patients**  **(Walsh 2021)** | **Function**  **score** | **Function**  **category** | **Peak Current**  **(norm.)** | **ΔV1/2**  **Act** | **LLR** | **LLR evidence**  **strength** |
| --- | --- | --- | --- | --- | --- | --- | --- |
| G119V | 1 | -0.13 | Loss | 0.18 | NA | 1.96 | Pathogenic strong |
| L353P | 2 | -0.06 | Loss | 0.16 | NA | 1.94 | Pathogenic strong |
| I514T | 6 | 0.37 | Partial Loss | 0.66 | -12.9 | 0.84 | Pathogenic moderate |
| L282P | 1 | 0.39 | Partial Loss | 0.21 | NA | 0.78 | Pathogenic moderate |
| P570L | 4 | 0.68 | Normal | 0.46 | -7.7 | -0.22 | No evidence |
| S143F | 1 | 1.17 | Normal | 0.79 | 17.3 | -0.94 | Benign moderate |

Patient counts are from Walsh et al, 2021 (PMID 32893267). Function score and category are from the variant-only MAVE scores. Peak current (normalized to wildtype) and ΔV1/2 Act are from automated patch clamping in this study. LLR=Log likelihood ratio.

**Table S7: Variants with elevated SpliceAI scores**

| **Mutation** | **Type** | **Acceptor Loss** | **Donor Loss** | **Acceptor Gain** | **Donor Gain** | **Merged** |
| --- | --- | --- | --- | --- | --- | --- |
| P61T | missense | 0 | 0 | 0.29 | 0 | 0.29 |
| P84Q | missense | 0 | 0 | 0.21 | 0 | 0.21 |
| S86S | synonymous | 0 | 0 | 0.21 | 0 | 0.21 |
| C122W | missense | 0 | 0 | 0.3 | 0.02 | 0.31 |
| I138I | synonymous | 0.19 | 0.01 | 0 | 0 | 0.2 |
| M159L | missense | 0 | 0 | 0.22 | 0.01 | 0.23 |
| E160K | missense | 0.68 | 0 | 0 | 0 | 0.68 |
| E160Q | missense | 0.87 | 0.01 | 0 | 0 | 0.87 |
| S182S | synonymous | 0 | 0 | 0.97 | 0 | 0.97 |
| K183R | missense | 0 | 0 | 0.36 | 0 | 0.36 |
| V185V | synonymous | 0 | 0 | 0.27 | 0.01 | 0.28 |
| R190G | missense | 0 | 0 | 0.34 | 0.11 | 0.41 |
| R190W | missense | 0 | 0 | 0.24 | 0.02 | 0.26 |
| L191L | synonymous | 0 | 0 | 0.23 | 0.01 | 0.24 |
| L191V | missense | 0 | 0 | 0.32 | 0.05 | 0.35 |
| R192G | missense | 0 | 0 | 0.28 | 0.03 | 0.3 |
| R195P | missense | 0 | 0 | 0.21 | 0.01 | 0.22 |
| R195R | synonymous | 0 | 0 | 0.32 | 0.14 | 0.42 |
| V221V | synonymous | 0 | 0 | 0.36 | 0 | 0.36 |
| F222C | missense | 0 | 0 | 0.42 | 0 | 0.42 |
| T224R | missense | 0 | 0 | 0.23 | 0 | 0.23 |
| R228S | missense | 0.43 | 0.01 | 0 | 0 | 0.44 |
| R228T | missense | 0 | 0 | 0.09 | 0.15 | 0.23 |
| G229G | synonymous | 0.26 | 0 | 0 | 0 | 0.26 |
| I230I | synonymous | 0.26 | 0 | 0 | 0 | 0.26 |
| R231C | missense | 0.29 | 0 | 0 | 0 | 0.29 |
| R231L | missense | 0.39 | 0 | 0 | 0 | 0.39 |
| R231P | missense | 0.21 | 0 | 0 | 0 | 0.21 |
| R231R | synonymous | 0.23 | 0 | 0 | 0 | 0.23 |
| R231S | missense | 0.26 | 0 | 0 | 0 | 0.26 |
| L233L | synonymous | 0.35 | 0 | 0 | 0 | 0.35 |
| I235V | missense | 0.26 | 0 | 0 | 0 | 0.26 |
| G245V | missense | 0 | 0 | 0.94 | 0.1 | 0.95 |
| Q260H | missense | 0 | 0 | 0.3 | 0.01 | 0.31 |
| E261K | missense | 0.76 | 0.01 | 0 | 0 | 0.76 |
| E261Q | missense | 0.9 | 0.01 | 0 | 0 | 0.9 |
| L262M | missense | 0.23 | 0 | 0 | 0 | 0.23 |
| L262Q | missense | 0.67 | 0 | 0 | 0 | 0.67 |
| I263V | missense | 0.32 | 0 | 0 | 0 | 0.32 |
| I268I | synonymous | 0.37 | 0.01 | 0 | 0 | 0.38 |
| L271L | synonymous | 0.87 | 0 | 0 | 0 | 0.87 |
| L271Q | missense | 0.9 | 0 | 0 | 0 | 0.9 |
| F279L | missense | 0.42 | 0 | 0 | 0 | 0.42 |
| L282L | synonymous | 0.79 | 0 | 0 | 0 | 0.79 |
| L282Q | missense | 0.84 | 0 | 0 | 0 | 0.84 |
| A283A | synonymous | 0.34 | 0 | 0 | 0 | 0.34 |
| N289S | missense | 0 | 0 | 0.23 | 0 | 0.23 |
| F296V | missense | 0 | 0 | 0.3 | 0 | 0.3 |
| G297G | synonymous | 0 | 0 | 0.21 | 0 | 0.21 |
| S298R | missense | 0 | 0 | 1 | 0.96 | 1 |
| D301G | missense | 0 | 0 | 1 | 0.78 | 1 |
| W304R | missense | 0 | 0 | 0.64 | 0 | 0.64 |
| G306V | missense | 0 | 0 | 0.43 | 0 | 0.43 |
| V307V | synonymous | 0 | 0 | 0.25 | 0.17 | 0.38 |
| T322R | missense | 0 | 0 | 0.31 | 0 | 0.31 |
| G325G | synonymous | 0 | 0 | 0.88 | 0 | 0.88 |
| A344A | synonymous | 0 | 0 | 0.55 | 0.55 | 0.8 |
| G345R | missense | 0.04 | 0.33 | 0 | 0 | 0.36 |
| G345V | missense | 0.2 | 0 | 0 | 0 | 0.2 |
| G345W | missense | 0.05 | 0.33 | 0 | 0 | 0.36 |
| L347H | missense | 0.32 | 0 | 0 | 0 | 0.32 |
| G348D | missense | 0.42 | 0.02 | 0 | 0 | 0.43 |
| G348G | synonymous | 0.04 | 0.25 | 0 | 0 | 0.28 |
| G348S | missense | 0.25 | 0.16 | 0 | 0 | 0.37 |
| A352S | missense | 0.16 | 0.07 | 0 | 0 | 0.22 |
| L353L | synonymous | 0.14 | 0.29 | 0 | 0 | 0.39 |
| K354Q | missense | 0.77 | 0.51 | 0 | 0 | 0.89 |
| F364F | synonymous | 0 | 0 | 0 | 0.2 | 0.2 |
| F364L | missense | 0 | 0 | 0 | 0.22 | 0.22 |
| Q376H | missense | 0 | 0 | 0 | 0.92 | 0.92 |
| Q376L | missense | 0 | 0 | 0 | 0.72 | 0.72 |
| Q376P | missense | 0 | 0 | 0 | 0.62 | 0.62 |
| Q376R | missense | 0 | 0 | 0 | 0.67 | 0.67 |
| T377T | synonymous | 0.84 | 0 | 0 | 0 | 0.84 |
| K414R | missense | 0 | 0 | 0.45 | 0.01 | 0.46 |
| S415Y | missense | 0 | 0 | 0.31 | 0 | 0.31 |
| V417V | synonymous | 0 | 0 | 0.3 | 0.05 | 0.34 |
| K419E | missense | 0.34 | 0 | 0 | 0 | 0.34 |
| G460G | synonymous | 0 | 0 | 0.79 | 0.02 | 0.79 |
| S463R | missense | 0 | 0 | 0.9 | 0.08 | 0.91 |
| V465I | missense | 0 | 0 | 0 | 0.51 | 0.51 |
| V465L | missense | 0 | 0 | 0 | 0.51 | 0.51 |
| V465V | synonymous | 0.35 | 0 | 0 | 0 | 0.35 |
| R466G | missense | 0.76 | 0 | 0 | 0 | 0.76 |
| G493G | synonymous | 0 | 0 | 0.95 | 0.29 | 0.96 |
| V516V | synonymous | 0 | 0 | 0.21 | 0 | 0.21 |
| Q531L | missense | 0.2 | 0 | 0 | 0 | 0.2 |
| Q531P | missense | 0.29 | 0 | 0 | 0 | 0.29 |
| Q531Q | synonymous | 0.95 | 0 | 0 | 0 | 0.95 |
| A532A | synonymous | 0.25 | 0 | 0 | 0 | 0.25 |
| A532E | missense | 0.23 | 0 | 0 | 0 | 0.23 |
| L552L | synonymous | 0 | 0 | 0.22 | 0 | 0.22 |
| M553K | missense | 0 | 0 | 0.4 | 0.06 | 0.44 |
| V554V | synonymous | 0 | 0 | 0.4 | 0.07 | 0.44 |
| R555R | synonymous | 0 | 0 | 0.42 | 0.47 | 0.69 |
| R555S | missense | 0 | 0 | 0.41 | 0.32 | 0.6 |
| I556I | synonymous | 0 | 0 | 0.21 | 0.01 | 0.22 |
| K557N | missense | 0 | 0 | 0.2 | 0.01 | 0.21 |
| K557R | missense | 0 | 0 | 0.25 | 0.01 | 0.26 |
| E558G | missense | 0 | 0 | 0.36 | 0.03 | 0.38 |
| E558V | missense | 0 | 0 | 0.6 | 0.02 | 0.61 |
| L559L | synonymous | 0 | 0 | 0.2 | 0 | 0.2 |
| L559V | missense | 0 | 0 | 0.38 | 0.01 | 0.39 |
| R561G | missense | 0 | 0 | 0.41 | 0.1 | 0.47 |
| R561S | missense | 0 | 0 | 0.94 | 0.03 | 0.94 |
| R562G | missense | 0 | 0 | 0.42 | 0.48 | 0.7 |
| R562K | missense | 0 | 0 | 0.55 | 0.53 | 0.79 |
| R562M | missense | 0 | 0 | 0.49 | 0.56 | 0.78 |
| R562R | synonymous | 0 | 0 | 0.38 | 0.21 | 0.51 |
| R562T | missense | 0 | 0 | 0.59 | 0.83 | 0.93 |
| R562W | missense | 0 | 0 | 0.82 | 0.38 | 0.89 |
| L563Q | missense | 0.34 | 0.01 | 0 | 0 | 0.35 |
| G568G | synonymous | 0 | 0.19 | 0.78 | 0 | 0.82 |
| E578K | missense | 0 | 0 | 0 | 0.61 | 0.61 |
| E578Q | missense | 0 | 0 | 0.01 | 0.73 | 0.73 |
| K579E | missense | 0.35 | 0 | 0 | 0 | 0.35 |
| K579Q | missense | 0.52 | 0 | 0 | 0 | 0.52 |
| K598N | missense | 0 | 0 | 0.17 | 0.88 | 0.9 |
| L665V | missense | 0 | 0 | 0.2 | 0 | 0.2 |

**Table S8: Case-control risk ratios**

| **Category** | **Range** | **Description** | **RR** | **95% CI Lower** | **95% CI Upper** | **A** | **B** | **C** | **D** | **Variant #** |
| --- | --- | --- | --- | --- | --- | --- | --- | --- | --- | --- |
| Function scores | <0.25 | Loss | 263 | 241 | 286 | 274 | 3420 | 218 | 1610845 | 1422 |
|  | 0.25-0.58 | Partial Loss | 199 | 178 | 222 | 194 | 3500 | 255 | 1607469 | 1297 |
|  | 0.58-1.391 | Normal | 9.7 | 8.1 | 11.7 | 116 | 3578 | 5016 | 1535934 | 8495 |
|  | >1.391 | Gain | 2.9 | 0.4 | 20.2 | 1 | 3693 | 147 | 1560104 | 349 |
| AlphaMissense | <0.792 | No Path | 6.3 | 5 | 7.9 | 72 | 3622 | 4786 | 1525024 | 6915 |
|  | 0.792-0.906 | Supp Path | 43.3 | 33.2 | 56.7 | 49 | 3645 | 450 | 1605049 | 874 |
|  | 0.906-0.99 | Mod Path | 161 | 145 | 179 | 262 | 3432 | 501 | 1604964 | 2073 |
|  | >0.99 | Strong Path | 238 | 216 | 262 | 235 | 3459 | 225 | 1607343 | 6915 |
| Function and AlphaMissense | <0.58 | Concordant Path | 273 | 254 | 294 | 394 | 3300 | 310 | 1608981 | 2287 |
|  | 0.58-1.391 | Discordant | 73.3 | 59.4 | 90.5 | 74 | 3620 | 374 | 1602227 | 2019 |
|  | 0.58-1.391 | Concordant Benign | 1.8 | 1.1 | 3.1 | 14 | 3680 | 3109 | 1502288 | 2268 |
| K-means clusters | 1 | DN | 272 | 250 | 295 | 299 | 3395 | 224 | 1609763 | 1423 |
|  | 2 | Haploinsufficient | 172 | 139 | 213 | 53 | 3641 | 84 | 1612975 | 861 |
|  | 3 | Partial Loss | 60.7 | 49 | 75.1 | 74 | 3620 | 453 | 1560226 | 1517 |
|  | 4 | Normal 1 | 20 | 16.6 | 24.1 | 110 | 3584 | 2261 | 1542328 | 2929 |
|  | 5 | Normal 2 | 7.3 | 5.3 | 10.2 | 35 | 3659 | 1967 | 1528262 | 3143 |
| Abundance scores | <0.25 | Loss | 239 | 220 | 260 | 321 | 3373 | 319 | 1603566 | 2329 |
|  | 0.25-0.61 | Partial Loss | 68.9 | 55 | 86.4 | 65 | 3629 | 345 | 1574166 | 1351 |
|  | 0.61-1.29 | Normal | 18 | 15.7 | 20.6 | 212 | 3482 | 5012 | 1536792 | 8645 |
|  | >1.29 | Gain | 51.2 | 32.3 | 81 | 16 | 3678 | 111 | 1490152 | 305 |
| Het Function scores | <0.25 | Loss | 228 | 209 | 249 | 299 | 3395 | 324 | 1609692 | 1426 |
|  | 0.25-0.363 | Partial Loss | 181 | 145 | 226 | 47 | 3647 | 66 | 1582273 | 359 |
|  | 0.363-1.717 | Normal | 21.5 | 19 | 24.3 | 254 | 3440 | 5040 | 1535656 | 9655 |
|  | >1.717 | Gain | 1.7 | 0.2 | 11.9 | 1 | 3693 | 251 | 1565037 | 566 |
| Het Abundance scores | <0.25 | Loss | 234 | 210 | 260 | 184 | 3510 | 178 | 1608746 | 921 |
|  | 0.25-0.676 | Partial Loss | 114 | 99.8 | 130 | 177 | 3517 | 521 | 1574456 | 1872 |
|  | 0.676-1.325 | Normal | 21.2 | 18.6 | 24.2 | 218 | 3476 | 4339 | 1537287 | 7401 |
|  | >1.325 | Gain | 57.5 | 39 | 84.8 | 22 | 3672 | 136 | 1512242 | 546 |
| Dominant negative | 4/4 Loss | Severe | 290 | 252 | 333 | 74 | 3620 | 40 | 1613150 | 290 |
|  | 3/4 Loss | Strong | 174 | 146 | 208 | 77 | 3617 | 120 | 1607129 | 548 |
| VARITY_R_LOO | <0.675 | No Path | 7 | 5.6 | 8.8 | 79 | 3615 | 4665 | 1520664 | 2286 |
|  | 0.675-0.842 | Supp Path | 73.9 | 58.4 | 93.5 | 59 | 3635 | 295 | 1609008 | 356 |
|  | 0.842-0.965 | Mod Path | 112 | 100 | 125 | 263 | 3431 | 840 | 1606452 | 872 |
|  | >0.965 | Strong Path | 264 | 241 | 290 | 214 | 3480 | 162 | 1612133 | 463 |
| REVEL | <0.644 | No Path | 1.1 | 0.6 | 2.2 | 9 | 3685 | 3212 | 1489002 | 1517 |
|  | 0.644-0.773 | Supp Path | 13.5 | 9.2 | 19.9 | 25 | 3669 | 783 | 1600247 | 701 |
|  | 0.773-0.932 | Mod Path | 72.4 | 64.5 | 81.3 | 264 | 3430 | 1444 | 1603972 | 1249 |
|  | >0.932 | Strong Path | 180 | 164 | 198 | 317 | 3377 | 523 | 1609165 | 510 |

Risk ratios (RR) and 95% confidence intervals (CI) comparing the frequency of variants in LQT cases versus gnomAD controls, stratified by category. These values correspond to the odds ratio plots in Figure 6E and Figure S17. Columns A, B, C, and D refer to case-control counts used to calculate risk ratios (see methods). Variant # indicates the total number of missense variants in the category.

**Table S9: *KCNQ1* plasmid zone system**

| **Zone** | ***KCNQ1* cDNA coordinates** | **Upstream Restriction Enzyme** | **Downstream Restriction Enzyme** |
| --- | --- | --- | --- |
| 1 | 1-704 | ClaI | Bsu36I |
| 2 | 712-1177 | Bsu36I | BglII |
| 3 | 1184-2031 | BglII | AgeI |

**Table S10: QuikChange primers used in this study**

| ag2044 | ccaacagcttcgccAaggacctggacctg | E487K Quikchange |
| --- | --- | --- |
| ag2057 | gcgggcggcgcTctctacgcgcc | A49A Quikchange |
| ag2058 | ccgcgcccccGgcgtccccgg | P64P Quikchange |
| ag2059 | gaccttggcccTcgtccgccggt | P81P Quikchange |
| ag2060 | gcgcgcacccaTgtccagggccg | H105H Quikchange |
| ag2061 | gcccaacactgctggaagtTagcatgcccc | V474V Quikchange |
| ag2062 | cccccggcagTggcggccccc | S627S Quikchange |
| ag2063 | ctgctgtcacccagGcccaaacccaagaa | S409R Quikchange |
| ag2065 | gggattcttggctcggggtttAccctgaaggtg | A352T Quikchange |
| ag2066 | gcagggccacctcTacctcatggtgcg | N551Y Quikchange |
| ag2067 | cccacatctcacagGtgcgggaacaccat | L506V Quikchange |
| ag2069 | ggtcttcatccaccgcAaggagctgataaccac | Q260K Quikchange |
| ag2251 | TTTGCGCTCCCAGCAGGGATTCTTGGCTC | A344A Quikchange |
| ag2252 | TGCGCTCCCAGCCGGGATTCTTGGC | A344A Quikchange |
| ag2253 | TTTGCGCTCCCAGCTGGGATTCTTGGCTC | A344A Quikchange |

**Table S11: Illumina sequencing primers used in this study**

| **Primer ID** | **Sequence** |
| --- | --- |
| ag1591 | AATGATACGGCGACCACCGAGATCTACACATGAGATCATACACTCTTTCCCTACACGACGCTCTTCCGATCTTCTTCGCCCTTAGACACCAT |
| ag1592 | AATGATACGGCGACCACCGAGATCTACACGCAGAGCTGCACACTCTTTCCCTACACGACGCTCTTCCGATCTTCTTCGCCCTTAGACACCAT |
| ag1593 | AATGATACGGCGACCACCGAGATCTACACTGTCGCTGGTACACTCTTTCCCTACACGACGCTCTTCCGATCTTCTTCGCCCTTAGACACCAT |
| ag1594 | AATGATACGGCGACCACCGAGATCTACACCACTATCAACACACTCTTTCCCTACACGACGCTCTTCCGATCTTCTTCGCCCTTAGACACCAT |
| ag1595 | AATGATACGGCGACCACCGAGATCTACACCTCTGCAGCGACACTCTTTCCCTACACGACGCTCTTCCGATCTTCTTCGCCCTTAGACACCAT |
| ag1596 | AATGATACGGCGACCACCGAGATCTACACTCTCATGATAACACTCTTTCCCTACACGACGCTCTTCCGATCTTCTTCGCCCTTAGACACCAT |
| ag1597 | AATGATACGGCGACCACCGAGATCTACACTATCTTGTAGACACTCTTTCCCTACACGACGCTCTTCCGATCTTCTTCGCCCTTAGACACCAT |
| ag1598 | AATGATACGGCGACCACCGAGATCTACACCGCTCCACGAACACTCTTTCCCTACACGACGCTCTTCCGATCTTCTTCGCCCTTAGACACCAT |
| ag1599 | AATGATACGGCGACCACCGAGATCTACACATTGCCGAGTACACTCTTTCCCTACACGACGCTCTTCCGATCTTCTTCGCCCTTAGACACCAT |
| ag1600 | AATGATACGGCGACCACCGAGATCTACACGCCATTAGACACACTCTTTCCCTACACGACGCTCTTCCGATCTTCTTCGCCCTTAGACACCAT |
| ag1601 | AATGATACGGCGACCACCGAGATCTACACAGCACATCCTACACTCTTTCCCTACACGACGCTCTTCCGATCTTCTTCGCCCTTAGACACCAT |
| ag1602 | AATGATACGGCGACCACCGAGATCTACACGATGTGCTTCACACTCTTTCCCTACACGACGCTCTTCCGATCTTCTTCGCCCTTAGACACCAT |
| ag1603 | CAAGCAGAAGACGGCATACGAGATGCCGACAAGAGTGACTGGAGTTCAGACGTGTGCTCTTCCGATCTcggcAattccggaCGTACG |
| ag1604 | CAAGCAGAAGACGGCATACGAGATATTAGTGGAGGTGACTGGAGTTCAGACGTGTGCTCTTCCGATCTcggcAattccggaCGTACG |
| ag1605 | CAAGCAGAAGACGGCATACGAGATCTGGCTTGCCGTGACTGGAGTTCAGACGTGTGCTCTTCCGATCTcggcAattccggaCGTACG |
| ag1606 | CAAGCAGAAGACGGCATACGAGATTCAATCCATTGTGACTGGAGTTCAGACGTGTGCTCTTCCGATCTcggcAattccggaCGTACG |
| ag1609 | CAAGCAGAAGACGGCATACGAGATCTGTTGGTCCGTGACTGGAGTTCAGACGTGTGCTCTTCCGATCTcggcAattccggaCGTACG |
| ag1610 | CAAGCAGAAGACGGCATACGAGATTCACCAACTTGTGACTGGAGTTCAGACGTGTGCTCTTCCGATCTcggcAattccggaCGTACG |
| ag1611 | CAAGCAGAAGACGGCATACGAGATGGTGCTATATGTGACTGGAGTTCAGACGTGTGCTCTTCCGATCTcggcAattccggaCGTACG |
| ag1612 | CAAGCAGAAGACGGCATACGAGATAACATCGCGCGTGACTGGAGTTCAGACGTGTGCTCTTCCGATCTcggcAattccggaCGTACG |
| ag1613 | CAAGCAGAAGACGGCATACGAGATGGTGGACGTGGTGACTGGAGTTCAGACGTGTGCTCTTCCGATCTcggcAattccggaCGTACG |
| ag1614 | CAAGCAGAAGACGGCATACGAGATAACAAGTACAGTGACTGGAGTTCAGACGTGTGCTCTTCCGATCTcggcAattccggaCGTACG |
| ag1615 | CAAGCAGAAGACGGCATACGAGATACCGTTACAAGTGACTGGAGTTCAGACGTGTGCTCTTCCGATCTcggcAattccggaCGTACG |
| ag1616 | CAAGCAGAAGACGGCATACGAGATGTTACCGTGGGTGACTGGAGTTCAGACGTGTGCTCTTCCGATCTcggcAattccggaCGTACG |
| ag1966 | CAAGCAGAAGACGGCATACGAGATTCTCTAGATTGTGACTGGAGTTCAGACGTGTGCTCTTCCGATCTATCGTCcggcAattccggaCGTACG |
| ag1967 | CAAGCAGAAGACGGCATACGAGATGGTAACGCAGGTGACTGGAGTTCAGACGTGTGCTCTTCCGATCTGCTGATcggcAattccggaCGTACG |
| ag1968 | CAAGCAGAAGACGGCATACGAGATAACGGTATGAGTGACTGGAGTTCAGACGTGTGCTCTTCCGATCTCCTAGCcggcAattccggaCGTACG |
| ag1969 | CAAGCAGAAGACGGCATACGAGATTCGACTTAAGGTGACTGGAGTTCAGACGTGTGCTCTTCCGATCTTACGCAcggcAattccggaCGTACG |
| ag1970 | CAAGCAGAAGACGGCATACGAGATCTAGTCCGGAGTGACTGGAGTTCAGACGTGTGCTCTTCCGATCTAGTCTGcggcAattccggaCGTACG |
| ag1971 | CAAGCAGAAGACGGCATACGAGATGAGCAAGGCCGTGACTGGAGTTCAGACGTGTGCTCTTCCGATCTCATCGTcggcAattccggaCGTACG |
| ag1972 | CAAGCAGAAGACGGCATACGAGATAGATGGAATTGTGACTGGAGTTCAGACGTGTGCTCTTCCGATCTTGCTGAcggcAattccggaCGTACG |
| ag1973 | CAAGCAGAAGACGGCATACGAGATGGCGAATTCTGTGACTGGAGTTCAGACGTGTGCTCTTCCGATCTGCCAGTcggcAattccggaCGTACG |
| ag1974 | CAAGCAGAAGACGGCATACGAGATAATAGGCCTCGTGACTGGAGTTCAGACGTGTGCTCTTCCGATCTCTACGTcggcAattccggaCGTACG |
| ag1975 | CAAGCAGAAGACGGCATACGAGATTGGTGCCTGGGTGACTGGAGTTCAGACGTGTGCTCTTCCGATCTTCGAGCcggcAattccggaCGTACG |
| ag1976 | CAAGCAGAAGACGGCATACGAGATCAACATTCAAGTGACTGGAGTTCAGACGTGTGCTCTTCCGATCTCGTACTcggcAattccggaCGTACG |
| ag1977 | CAAGCAGAAGACGGCATACGAGATTAACTTAGCGGTGACTGGAGTTCAGACGTGTGCTCTTCCGATCTACAGTCcggcAattccggaCGTACG |
| ag1978 | AATGATACGGCGACCACCGAGATCTACACAGGTCAGATAACACTCTTTCCCTACACGACGCTCTTCCGATCTATCGTCtgtgcatgttctccttaatcagc |
| ag1979 | AATGATACGGCGACCACCGAGATCTACACAAGGCCACGGACACTCTTTCCCTACACGACGCTCTTCCGATCTGCTGATtgtgcatgttctccttaatcagc |
| ag1980 | AATGATACGGCGACCACCGAGATCTACACGGAATTGTAAACACTCTTTCCCTACACGACGCTCTTCCGATCTCCTAGCtgtgcatgttctccttaatcagc |
| ag1981 | AATGATACGGCGACCACCGAGATCTACACCCTGACCACTACACTCTTTCCCTACACGACGCTCTTCCGATCTTACGCAtgtgcatgttctccttaatcagc |
| ag1982 | AATGATACGGCGACCACCGAGATCTACACTTCAGTTGTCACACTCTTTCCCTACACGACGCTCTTCCGATCTAGTCTGtgtgcatgttctccttaatcagc |
| ag1983 | AATGATACGGCGACCACCGAGATCTACACCGCATTCCGTACACTCTTTCCCTACACGACGCTCTTCCGATCTCATCGTtgtgcatgttctccttaatcagc |
| ag1984 | AATGATACGGCGACCACCGAGATCTACACTATGCCTTACACACTCTTTCCCTACACGACGCTCTTCCGATCTTGCTGAtgtgcatgttctccttaatcagc |
| ag1985 | AATGATACGGCGACCACCGAGATCTACACGTATTGACGTACACTCTTTCCCTACACGACGCTCTTCCGATCTGCCAGTtgtgcatgttctccttaatcagc |
| ag1986 | AATGATACGGCGACCACCGAGATCTACACACGCCAGTACACACTCTTTCCCTACACGACGCTCTTCCGATCTCTACGTtgtgcatgttctccttaatcagc |
| ag1987 | AATGATACGGCGACCACCGAGATCTACACAACCATAGAAACACTCTTTCCCTACACGACGCTCTTCCGATCTTCGAGCtgtgcatgttctccttaatcagc |
| ag1988 | AATGATACGGCGACCACCGAGATCTACACGGTTGCGAGGACACTCTTTCCCTACACGACGCTCTTCCGATCTCGTACTtgtgcatgttctccttaatcagc |
| ag1989 | AATGATACGGCGACCACCGAGATCTACACTTACGCACCTACACTCTTTCCCTACACGACGCTCTTCCGATCTACAGTCtgtgcatgttctccttaatcagc |
| ag2037 | CAAGCAGAAGACGGCATACGAGATCCACCTGTGTGTGACTGGAGTTCAGACGTGTGCTCTTCCGATCTGTAGCTcggcAattccggaCGTACG |
| ag2038 | CAAGCAGAAGACGGCATACGAGATTGCCTGGTGGGTGACTGGAGTTCAGACGTGTGCTCTTCCGATCTTGCATGcggcAattccggaCGTACG |
| ag2039 | CAAGCAGAAGACGGCATACGAGATCATTCAACAAGTGACTGGAGTTCAGACGTGTGCTCTTCCGATCTCGACTAcggcAattccggaCGTACG |
| ag2040 | AATGATACGGCGACCACCGAGATCTACACTTGGAATTCCACACTCTTTCCCTACACGACGCTCTTCCGATCTGTAGCTtgtgcatgttctccttaatcagc |
| ag2041 | AATGATACGGCGACCACCGAGATCTACACTAATGTGTCTACACTCTTTCCCTACACGACGCTCTTCCGATCTTGCATGtgtgcatgttctccttaatcagc |
| ag2042 | AATGATACGGCGACCACCGAGATCTACACCGGCACACTCACACTCTTTCCCTACACGACGCTCTTCCGATCTCGACTAtgtgcatgttctccttaatcagc |

**Table S12: Summary of MAVE sequencing samples and primer pairs**

| **Experiment** | **Sample ID** | **Description** | **Primer Pairs** | **i7 sequence** | **i5 sequence** |
| --- | --- | --- | --- | --- | --- |
| Abundance | 8952- AG-1 | KCNQ1-myc variant library (zone 1), Replicate 1 Bin 1 | ag1591+ag1609 | GGACCAACAG | ATGATCTCAT |
| Abundance | 8952- AG-2 | KCNQ1-myc variant library (zone 1), Replicate 1 Bin 2 | ag1592+ag1610 | AAGTTGGTGA | GCAGCTCTGC |
| Abundance | 8952- AG-3 | KCNQ1-myc variant library (zone 1), Replicate 1 Bin 3 | ag1593+ag1611 | ATATAGCACC | ACCAGCGACA |
| Abundance | 8952- AG-4 | KCNQ1-myc variant library (zone 1), Replicate 1 Bin 4 | ag1594+ag1612 | GCGCGATGTT | GTTGATAGTG |
| Abundance | 8952- AG-5 | KCNQ1-myc variant library (zone 1), Replicate 2 Bin 1 | ag1595+ag1613 | CACGTCCACC | CGCTGCAGAG |
| Abundance | 8952- AG-6 | KCNQ1-myc variant library (zone 1), Replicate 2 Bin 2 | ag1596+ag1614 | TGTACTTGTT | TATCATGAGA |
| Abundance | 8952- AG-7 | KCNQ1-myc variant library (zone 1), Replicate 2 Bin 3 | ag1597+ag1615 | TTGTAACGGT | CTACAAGATA |
| Abundance | 8952- AG-8 | KCNQ1-myc variant library (zone 1), Replicate 2 Bin 4 | ag1598+ag1616 | CCACGGTAAC | TCGTGGAGCG |
| Abundance | 8952- AG-9 | KCNQ1-myc variant library (zone 1), Replicate 3 Bin 1 | ag1599+ag1603 | TCTTGTCGGC | ACTCGGCAAT |
| Abundance | 8952- AG-10 | KCNQ1-myc variant library (zone 1), Replicate 3 Bin 2 | ag1600+ag1604 | CTCCACTAAT | GTCTAATGGC |
| Abundance | 8952-AG-11 | KCNQ1-myc variant library (zone 1), Replicate 3 Bin 3 | ag1601+ag1605 | GGCAAGCCAG | AGGATGTGCT |
| Abundance | 8952-AG-12 | KCNQ1-myc variant library (zone 1), Replicate 3 Bin 4 | ag1602+ag1606 | AATGGATTGA | GAAGCACATC |
| Abundance | 9004-AG-1 | KCNQ1-myc variant library (zone 2), Replicate 1 Bin 1 | ag1591+ag1609 | GGACCAACAG | ATGATCTCAT |
| Abundance | 9004-AG-2 | KCNQ1-myc variant library (zone 2), Replicate 1 Bin 2 | ag1592+ag1610 | AAGTTGGTGA | GCAGCTCTGC |
| Abundance | 9004-AG-3 | KCNQ1-myc variant library (zone 2), Replicate 1 Bin 3 | ag1593+ag1611 | ATATAGCACC | ACCAGCGACA |
| Abundance | 9004-AG-4 | KCNQ1-myc variant library (zone 2), Replicate 1 Bin 4 | ag1594+ag1612 | GCGCGATGTT | GTTGATAGTG |
| Abundance | 9004-AG-5 | KCNQ1-myc variant library (zone 2), Replicate 2 Bin 1 | ag1595+ag1613 | CACGTCCACC | CGCTGCAGAG |
| Abundance | 9004-AG-6 | KCNQ1-myc variant library (zone 2), Replicate 2 Bin 2 | ag1596+ag1614 | TGTACTTGTT | TATCATGAGA |
| Abundance | 9004-AG-7 | KCNQ1-myc variant library (zone 2), Replicate 2 Bin 3 | ag1597+ag1615 | TTGTAACGGT | CTACAAGATA |
| Abundance | 9004-AG-8 | KCNQ1-myc variant library (zone 2), Replicate 2 Bin 4 | ag1598+ag1616 | CCACGGTAAC | TCGTGGAGCG |
| Abundance | 9814-AG-1 | KCNQ1-myc variant library (zone 2), Replicate 3 Bin 1 | ag1591+ag1609 | GGACCAACAG | ATGATCTCAT |
| Abundance | 9814-AG-2 | KCNQ1-myc variant library (zone 2), Replicate 3 Bin 2 | ag1592+ag1610 | AAGTTGGTGA | GCAGCTCTGC |
| Abundance | 9814-AG-3 | KCNQ1-myc variant library (zone 2), Replicate 3 Bin 3 | ag1593+ag1611 | ATATAGCACC | ACCAGCGACA |
| Abundance | 9814-AG-4 | KCNQ1-myc variant library (zone 2), Replicate 3 Bin 4 | ag1594+ag1612 | GCGCGATGTT | GTTGATAGTG |
| Abundance | 9612-AG-1 | KCNQ1-myc variant library (zone 3), Replicate 1 Bin 1 | ag1591+ag1609 | GGACCAACAG | ATGAGATCAT |
| Abundance | 9612-AG-2 | KCNQ1-myc variant library (zone 3), Replicate 1 Bin 2 | ag1592+ag1610 | AAGTTGGTGA | GCAGAGCTGC |
| Abundance | 9612-AG-3 | KCNQ1-myc variant library (zone 3), Replicate 1 Bin 3 | ag1593+ag1611 | ATATAGCACC | TGTCGCTGGT |
| Abundance | 9612-AG-4 | KCNQ1-myc variant library (zone 3), Replicate 1 Bin 4 | ag1594+ag1612 | GCGCGATGTT | CACTATCAAC |
| Abundance | 9612-AG-5 | KCNQ1-myc variant library (zone 3), Replicate 2 Bin 1 | ag1595+ag1613 | CACGTCCACC | CTCTGCAGCG |
| Abundance | 9612-AG-6 | KCNQ1-myc variant library (zone 3), Replicate 2 Bin 2 | ag1596+ag1614 | TGTACTTGTT | TCTCATGATA |
| Abundance | 9612-AG-7 | KCNQ1-myc variant library (zone 3), Replicate 2 Bin 3 | ag1597+ag1615 | TTGTAACGGT | TATCTTGTAG |
| Abundance | 9612-AG-8 | KCNQ1-myc variant library (zone 3), Replicate 2 Bin 4 | ag1598+ag1616 | CCACGGTAAC | CGCTCCACGA |
| Abundance | 9612-AG-9 | KCNQ1-myc variant library (zone 3), Replicate 3 Bin 1 | ag1599+ag1603 | TCTTGTCGGC | ATTGCCGAGT |
| Abundance | 9612-AG-10 | KCNQ1-myc variant library (zone 3), Replicate 3 Bin 2 | ag1600+ag1604 | CTCCACTAAT | GCCATTAGAC |
| Abundance | 9612-AG-11 | KCNQ1-myc variant library (zone 3), Replicate 3 Bin 3 | ag1601+ag1605 | GGCAAGCCAG | AGCACATCCT |
| Abundance | 9612-AG-12 | KCNQ1-myc variant library (zone 3), Replicate 3 Bin 4 | ag1602+ag1606 | AATGGATTGA | GATGTGCTTC |
| Abundance | 10029-MH-1 | KCNQ1-myc variant library (zones 1-3), Replicate 1 Bin 1 | ag1591+ag1609 | GGACCAACAG | ATGATCTCAT |
| Abundance | 10029-MH-2 | KCNQ1-myc variant library (zones 1-3), Replicate 1 Bin 2 | ag1592+ag1610 | AAGTTGGTGA | GCAGCTCTGC |
| Abundance | 10029-MH-3 | KCNQ1-myc variant library (zones 1-3), Replicate 1 Bin 3 | ag1593+ag1611 | ATATAGCACC | ACCAGCGACA |
| Abundance | 10029-MH-4 | KCNQ1-myc variant library (zones 1-3), Replicate 1 Bin 4 | ag1594+ag1612 | GCGCGATGTT | GTTGATAGTG |
| Abundance | 10029-MH-5 | KCNQ1-myc variant library (zones 1-3), Replicate 2 Bin 1 | ag1595+ag1613 | CACGTCCACC | CGCTGCAGAG |
| Abundance | 10029-MH-6 | KCNQ1-myc variant library (zones 1-3), Replicate 2 Bin 2 | ag1596+ag1614 | TGTACTTGTT | TATCATGAGA |
| Abundance | 10029-MH-7 | KCNQ1-myc variant library (zones 1-3), Replicate 2 Bin 3 | ag1597+ag1615 | TTGTAACGGT | CTACAAGATA |
| Abundance | 10029-MH-8 | KCNQ1-myc variant library (zones 1-3), Replicate 2 Bin 4 | ag1598+ag1616 | CCACGGTAAC | TCGTGGAGCG |
| Heterozygous Abundance | 10211-MH-1 | KCNQ1-myc variant library (zones 1-3) + KCNQ1-HA, Replicate 1 Bin 1 | ag1591+ag1609 | GGACCAACAG | ATGAGATCAT |
| Heterozygous Abundance | 10211-MH-2 | KCNQ1-myc variant library (zones 1-3) + KCNQ1-HA, Replicate 1 Bin 2 | ag1592+ag1610 | AAGTTGGTGA | GCAGAGCTGC |
| Heterozygous Abundance | 10211-MH-3 | KCNQ1-myc variant library (zones 1-3) + KCNQ1-HA, Replicate 1 Bin 3 | ag1593+ag1611 | ATATAGCACC | TGTCGCTGGT |
| Heterozygous Abundance | 10211-MH-4 | KCNQ1-myc variant library (zones 1-3) + KCNQ1-HA, Replicate 1 Bin 4 | ag1594+ag1612 | GCGCGATGTT | CACTATCAAC |
| Heterozygous Abundance | 10211-MH-5 | KCNQ1-myc variant library (zones 1-3) + KCNQ1-HA, Replicate 2 Bin 1 | ag1595+ag1613 | CACGTCCACC | CTCTGCAGCG |
| Heterozygous Abundance | 10211-MH-6 | KCNQ1-myc variant library (zones 1-3) + KCNQ1-HA, Replicate 2 Bin 2 | ag1596+ag1614 | TGTACTTGTT | TCTCATGATA |
| Heterozygous Abundance | 10211-MH-7 | KCNQ1-myc variant library (zones 1-3) + KCNQ1-HA, Replicate 2 Bin 3 | ag1597+ag1615 | TTGTAACGGT | TATCTTGTAG |
| Heterozygous Abundance | 10211-MH-8 | KCNQ1-myc variant library (zones 1-3) + KCNQ1-HA, Replicate 2 Bin 4 | ag1598+ag1616 | CCACGGTAAC | CGCTCCACGA |
| Heterozygous Abundance | 10211-MH-9 | KCNQ1-myc variant library (zones 1-3) + KCNQ1-HA, Replicate 3 Bin 1 | ag1599+ag1603 | TCTTGTCGGC | ATTGCCGAGT |
| Heterozygous Abundance | 10211-MH-10 | KCNQ1-myc variant library (zones 1-3) + KCNQ1-HA, Replicate 3 Bin 2 | ag1600+ag1604 | CTCCACTAAT | GCCATTAGAC |
| Heterozygous Abundance | 10211-MH-11 | KCNQ1-myc variant library (zones 1-3) + KCNQ1-HA, Replicate 3 Bin 3 | ag1601+ag1605 | GGCAAGCCAG | AGCACATCCT |
| Heterozygous Abundance | 10211-MH-12 | KCNQ1-myc variant library (zones 1-3) + KCNQ1-HA, Replicate 3 Bin 4 | ag1602+ag1606 | AATGGATTGA | GATGTGCTTC |
| Functional | 11823-MH-1 | KCNQ1-myc variant library (zones 1-3), Replicate 1 Day 0 | ag1966+ag1978 | AATCTAGAGA | AGGTCAGATA |
| Functional | 11823-MH-2 | KCNQ1-myc variant library (zones 1-3), Replicate 2 Day 0 | ag1967+ag1979 | CTGCGTTACC | AAGGCCACGG |
| Functional | 11823-MH-3 | KCNQ1-myc variant library (zones 1-3), Replicate 3 Day 0 | ag1968+ag1980 | TCATACCGTT | GGAATTGTAA |
| Functional | 11823-MH-4 | KCNQ1-myc variant library (zones 1-3), Replicate 1 Day 7 | ag1969+ag1981 | CTTAAGTCGA | CCTGACCACT |
| Functional | 11823-MH-5 | KCNQ1-myc variant library (zones 1-3), Replicate 2 Day 7 | ag1970+ag1982 | TCCGGACTAG | TTCAGTTGTC |
| Functional | 11823-MH-6 | KCNQ1-myc variant library (zones 1-3), Replicate 3 Day 7 | ag1971+ag1983 | GGCCTTGCTC | CGCATTCCGT |
| Functional | 11823-MH-7 | KCNQ1-myc variant library (zones 1-3), Replicate 1 Day 14 | ag1972+ag1984 | AATTCCATCT | TATGCCTTAC |
| Functional | 11823-MH-8 | KCNQ1-myc variant library (zones 1-3), Replicate 2 Day 14 | ag1973+ag1985 | AGAATTCGCC | GTATTGACGT |
| Functional | 11823-MH-9 | KCNQ1-myc variant library (zones 1-3), Replicate 3 Day 14 | ag1974+ag1986 | GAGGCCTATT | ACGCCAGTAC |
| Functional | 11823-MH-10 | KCNQ1-myc variant library (zones 1-3), Replicate 1 Day 21 | ag1975+ag1987 | CCAGGCACCA | AACCATAGAA |
| Functional | 11823-MH-11 | KCNQ1-myc variant library (zones 1-3), Replicate 2 Day 21 | ag1976+ag1988 | TTGAATGTTG | GGTTGCGAGG |
| Functional | 11823-MH-12 | KCNQ1-myc variant library (zones 1-3), Replicate 3 Day 21 | ag1977+ag1989 | CGCTAAGTTA | TTACGCACCT |
| Functional | 11823-MH-13 | KCNQ1-myc variant library (zones 1-3), Replicate 1 Day 28 | ag2037+ag2040 | ACACAGGTGG | TTGGAATTCC |
| Functional | 11823-MH-14 | KCNQ1-myc variant library (zones 1-3), Replicate 2 Day 28 | ag2038+ag2041 | CCACCAGGCA | TAATGTGTCT |
| Functional | 11823-MH-15 | KCNQ1-myc variant library (zones 1-3), Replicate 3 Day 28 | ag2039+ag2042 | TTGTTGAATG | CGGCACACTC |
| Functional | 12246-MH-1 | KCNQ1-myc variant library (zones 1-3), Replicate 4 Day 0 | ag1966+ag1978 | AATCTAGAGA | AGGTCAGATA |
| Functional | 12246-MH-2 | KCNQ1-myc variant library (zones 1-3), Replicate 5 Day 0 | ag1967+ag1979 | CTGCGTTACC | AAGGCCACGG |
| Functional | 12246-MH-3 | KCNQ1-myc variant library (zones 1-3), Replicate 6 Day 0 | ag1968+ag1980 | TCATACCGTT | GGAATTGTAA |
| Functional | 12246-MH-4 | KCNQ1-myc variant library (zones 1-3), Replicate 4 Day 7 | ag1969+ag1981 | CTTAAGTCGA | CCTGACCACT |
| Functional | 12246-MH-5 | KCNQ1-myc variant library (zones 1-3), Replicate 5 Day 7 | ag1970+ag1982 | TCCGGACTAG | TTCAGTTGTC |
| Functional | 12246-MH-6 | KCNQ1-myc variant library (zones 1-3), Replicate 6 Day 7 | ag1971+ag1983 | GGCCTTGCTC | CGCATTCCGT |
| Functional | 12246-MH-7 | KCNQ1-myc variant library (zones 1-3), Replicate 4 Day 14 | ag1972+ag1984 | AATTCCATCT | TATGCCTTAC |
| Functional | 12246-MH-8 | KCNQ1-myc variant library (zones 1-3), Replicate 5 Day 14 | ag1973+ag1985 | AGAATTCGCC | GTATTGACGT |
| Functional | 12246-MH-9 | KCNQ1-myc variant library (zones 1-3), Replicate 6 Day 14 | ag1974+ag1986 | GAGGCCTATT | ACGCCAGTAC |
| Functional | 12246-MH-10 | KCNQ1-myc variant library (zones 1-3), Replicate 4 Day 21 | ag1975+ag1987 | CCAGGCACCA | AACCATAGAA |
| Functional | 12246-MH-11 | KCNQ1-myc variant library (zones 1-3), Replicate 5 Day 21 | ag1976+ag1988 | TTGAATGTTG | GGTTGCGAGG |
| Functional | 12246-MH-12 | KCNQ1-myc variant library (zones 1-3), Replicate 6 Day 21 | ag1977+ag1989 | CGCTAAGTTA | TTACGCACCT |
| Functional | 12246-MH-13 | KCNQ1-myc variant library (zones 1-3), Replicate 4 Day 28 | ag2037+ag2040 | ACACAGGTGG | TTGGAATTCC |
| Functional | 12246-MH-14 | KCNQ1-myc variant library (zones 1-3), Replicate 5 Day 28 | ag2038+ag2041 | CCACCAGGCA | TAATGTGTCT |
| Functional | 12246-MH-15 | KCNQ1-myc variant library (zones 1-3), Replicate 6 Day 28 | ag2039+ag2042 | TTGTTGAATG | CGGCACACTC |
| Heterozygous Functional | 12331-MH-1 | KCNQ1-myc variant library (zones 1-3) + KCNQ1-HA, Replicate 1 Day 0 | ag1966+ag1978 | AATCTAGAGA | AGGTCAGATA |
| Heterozygous Functional | 12331-MH-2 | KCNQ1-myc variant library (zones 1-3) + KCNQ1-HA, Replicate 2 Day 0 | ag1967+ag1979 | CTGCGTTACC | AAGGCCACGG |
| Heterozygous Functional | 12331-MH-3 | KCNQ1-myc variant library (zones 1-3) + KCNQ1-HA, Replicate 3 Day 0 | ag1968+ag1980 | TCATACCGTT | GGAATTGTAA |
| Heterozygous Functional | 12331-MH-4 | KCNQ1-myc variant library (zones 1-3) + KCNQ1-HA, Replicate 4 Day 0 | ag1969+ag1981 | CTTAAGTCGA | CCTGACCACT |
| Heterozygous Functional | 12331-MH-5 | KCNQ1-myc variant library (zones 1-3) + KCNQ1-HA, Replicate 5 Day 0 | ag1970+ag1982 | TCCGGACTAG | TTCAGTTGTC |
| Heterozygous Functional | 12331-MH-6 | KCNQ1-myc variant library (zones 1-3) + KCNQ1-HA, Replicate 6 Day 0 | ag1971+ag1983 | GGCCTTGCTC | CGCATTCCGT |
| Heterozygous Functional | 12331-MH-7 | KCNQ1-myc variant library (zones 1-3) + KCNQ1-HA, Replicate 1 Day 7 | ag1972+ag1984 | AATTCCATCT | TATGCCTTAC |
| Heterozygous Functional | 12331-MH-8 | KCNQ1-myc variant library (zones 1-3) + KCNQ1-HA, Replicate 2 Day 7 | ag1973+ag1985 | AGAATTCGCC | GTATTGACGT |
| Heterozygous Functional | 12331-MH-9 | KCNQ1-myc variant library (zones 1-3) + KCNQ1-HA, Replicate 3 Day 7 | ag1974+ag1986 | GAGGCCTATT | ACGCCAGTAC |
| Heterozygous Functional | 12331-MH-10 | KCNQ1-myc variant library (zones 1-3) + KCNQ1-HA, Replicate 4 Day 7 | ag1975+ag1987 | CCAGGCACCA | AACCATAGAA |
| Heterozygous Functional | 12331-MH-11 | KCNQ1-myc variant library (zones 1-3) + KCNQ1-HA, Replicate 5 Day 7 | ag1976+ag1988 | TTGAATGTTG | GGTTGCGAGG |
| Heterozygous Functional | 12331-MH-12 | KCNQ1-myc variant library (zones 1-3) + KCNQ1-HA, Replicate 6 Day 7 | ag1977+ag1989 | CGCTAAGTTA | TTACGCACCT |
| Heterozygous Functional | 12331-MH-13 | KCNQ1-myc variant library (zones 1-3) + KCNQ1-HA, Replicate 1 Day 14 | ag2037+ag2040 | ACACAGGTGG | TTGGAATTCC |
| Heterozygous Functional | 12331-MH-14 | KCNQ1-myc variant library (zones 1-3) + KCNQ1-HA, Replicate 2 Day 14 | ag2038+ag2041 | CCACCAGGCA | TAATGTGTCT |
| Heterozygous Functional | 12331-MH-15 | KCNQ1-myc variant library (zones 1-3) + KCNQ1-HA, Replicate 3 Day 14 | ag2039+ag2042 | TTGTTGAATG | CGGCACACTC |
| Heterozygous Functional | 12332-MH-1 | KCNQ1-myc variant library (zones 1-3) + KCNQ1-HA, Replicate 4 Day 14 | ag1966+ag1978 | AATCTAGAGA | AGGTCAGATA |
| Heterozygous Functional | 12332-MH-2 | KCNQ1-myc variant library (zones 1-3) + KCNQ1-HA, Replicate 5 Day 14 | ag1967+ag1979 | CTGCGTTACC | AAGGCCACGG |
| Heterozygous Functional | 12332-MH-3 | KCNQ1-myc variant library (zones 1-3) + KCNQ1-HA, Replicate 6 Day 14 | ag1968+ag1980 | TCATACCGTT | GGAATTGTAA |
| Heterozygous Functional | 12332-MH-4 | KCNQ1-myc variant library (zones 1-3) + KCNQ1-HA, Replicate 1 Day 21 | ag1969+ag1981 | CTTAAGTCGA | CCTGACCACT |
| Heterozygous Functional | 12332-MH-5 | KCNQ1-myc variant library (zones 1-3) + KCNQ1-HA, Replicate 2 Day 21 | ag1970+ag1982 | TCCGGACTAG | TTCAGTTGTC |
| Heterozygous Functional | 12332-MH-6 | KCNQ1-myc variant library (zones 1-3) + KCNQ1-HA, Replicate 3 Day 21 | ag1971+ag1983 | GGCCTTGCTC | CGCATTCCGT |
| Heterozygous Functional | 12332-MH-7 | KCNQ1-myc variant library (zones 1-3) + KCNQ1-HA, Replicate 4 Day 21 | ag1972+ag1984 | AATTCCATCT | TATGCCTTAC |
| Heterozygous Functional | 12332-MH-8 | KCNQ1-myc variant library (zones 1-3) + KCNQ1-HA, Replicate 5 Day 21 | ag1973+ag1985 | AGAATTCGCC | GTATTGACGT |
| Heterozygous Functional | 12332-MH-9 | KCNQ1-myc variant library (zones 1-3) + KCNQ1-HA, Replicate 6 Day 21 | ag1974+ag1986 | GAGGCCTATT | ACGCCAGTAC |
| Heterozygous Functional | 12332-MH-10 | KCNQ1-myc variant library (zones 1-3) + KCNQ1-HA, Replicate 1 Day 28 | ag1975+ag1987 | CCAGGCACCA | AACCATAGAA |
| Heterozygous Functional | 12332-MH-11 | KCNQ1-myc variant library (zones 1-3) + KCNQ1-HA, Replicate 2 Day 28 | ag1976+ag1988 | TTGAATGTTG | GGTTGCGAGG |
| Heterozygous Functional | 12332-MH-12 | KCNQ1-myc variant library (zones 1-3) + KCNQ1-HA, Replicate 3 Day 28 | ag1977+ag1989 | CGCTAAGTTA | TTACGCACCT |
| Heterozygous Functional | 12332-MH-13 | KCNQ1-myc variant library (zones 1-3) + KCNQ1-HA, Replicate 4 Day 28 | ag2037+ag2040 | ACACAGGTGG | TTGGAATTCC |
| Heterozygous Functional | 12332-MH-14 | KCNQ1-myc variant library (zones 1-3) + KCNQ1-HA, Replicate 5 Day 28 | ag2038+ag2041 | CCACCAGGCA | TAATGTGTCT |
| Heterozygous Functional | 12332-MH-15 | KCNQ1-myc variant library (zones 1-3) + KCNQ1-HA, Replicate 6 Day 28 | ag2039+ag2042 | TTGTTGAATG | CGGCACACTC |
| Heterozygous Functional | 13772-MH-2 | KCNQ1-myc variant library (zones 1-3) + KCNQ1-HA, Replicate 7 Day 0 | ag1967+ag1979 | CTGCGTTACC | AAGGCCACGG |
| Heterozygous Functional | 13772-MH-3 | KCNQ1-myc variant library (zones 1-3) + KCNQ1-HA, Replicate 8 Day 0 | ag1968+ag1980 | TCATACCGTT | GGAATTGTAA |
| Heterozygous Functional | 13772-MH-4 | KCNQ1-myc variant library (zones 1-3) + KCNQ1-HA, Replicate 9 Day 0 | ag1969+ag1981 | CTTAAGTCGA | CCTGACCACT |
| Heterozygous Functional | 13772-MH-5 | KCNQ1-myc variant library (zones 1-3) + KCNQ1-HA, Replicate 10 Day 0 | ag1970+ag1982 | TCCGGACTAG | TTCAGTTGTC |
| Heterozygous Functional | 13772-MH-6 | KCNQ1-myc variant library (zones 1-3) + KCNQ1-HA, Replicate 11 Day 0 | ag1971+ag1983 | GGCCTTGCTC | CGCATTCCGT |
| Heterozygous Functional | 13772-MH-7 | KCNQ1-myc variant library (zones 1-3) + KCNQ1-HA, Replicate 12 Day 0 | ag1972+ag1984 | AATTCCATCT | TATGCCTTAC |
| Heterozygous Functional | 13772-MH-8 | KCNQ1-myc variant library (zones 1-3) + KCNQ1-HA, Replicate 13 Day 0 | ag1973+ag1985 | AGAATTCGCC | GTATTGACGT |
| Heterozygous Functional | 13772-MH-9 | KCNQ1-myc variant library (zones 1-3) + KCNQ1-HA, Replicate 14 Day 0 | ag1974+ag1986 | GAGGCCTATT | ACGCCAGTAC |
| Heterozygous Functional | 13773-MH-1 | KCNQ1-myc variant library (zones 1-3) + KCNQ1-HA, Replicate 7 Day 28 | ag1966+ag1978 | CTGCGTTACC | AAGGCCACGG |
| Heterozygous Functional | 13773-MH-2 | KCNQ1-myc variant library (zones 1-3) + KCNQ1-HA, Replicate 8 Day 28 | ag1967+ag1979 | TCATACCGTT | GGAATTGTAA |
| Heterozygous Functional | 13773-MH-3 | KCNQ1-myc variant library (zones 1-3) + KCNQ1-HA, Replicate 9 Day 28 | ag1968+ag1980 | CTTAAGTCGA | CCTGACCACT |
| Heterozygous Functional | 13773-MH-4 | KCNQ1-myc variant library (zones 1-3) + KCNQ1-HA, Replicate 10 Day 28 | ag1969+ag1981 | TCCGGACTAG | TTCAGTTGTC |
| Heterozygous Functional | 13773-MH-5 | KCNQ1-myc variant library (zones 1-3) + KCNQ1-HA, Replicate 11 Day 28 | ag1970+ag1982 | GGCCTTGCTC | CGCATTCCGT |
| Heterozygous Functional | 13773-MH-6 | KCNQ1-myc variant library (zones 1-3) + KCNQ1-HA, Replicate 12 Day 28 | ag1971+ag1983 | AATTCCATCT | TATGCCTTAC |
| Heterozygous Functional | 13773-MH-7 | KCNQ1-myc variant library (zones 1-3) + KCNQ1-HA, Replicate 13 Day 28 | ag1972+ag1984 | AGAATTCGCC | GTATTGACGT |
| Heterozygous Functional | 13773-MH-8 | KCNQ1-myc variant library (zones 1-3) + KCNQ1-HA, Replicate 14 Day 28 | ag1973+ag1985 | GAGGCCTATT | ACGCCAGTAC |
